# AI-based radiomics for pancreatic cysts: high diagnostic performance amid a persistent translational gap

**DOI:** 10.64898/2026.02.10.26345995

**Authors:** JD Lettner, T Evrenoglou, H Binder, S Fichtner-Feigl, C Neubauer, DA Ruess

**Affiliations:** Department of General and Visceral Surgery, Center for Surgery, Faculty of Medicine, Medical Center - University of Freiburg, Freiburg, Germany; Department of Interventional and Diagnostic Radiology. University Hospital Freiburg, Freiburg, Germany; Institute of Medical Biometry and Statistics, Faculty of Medicine and Medical Center—University of Freiburg, Germany

**Author notes:** Corresponding author: JD Lettner, Hugstetterstraße 55, 79106 Freiburg Germany, mail; Tel.: +49 0761/270 23650. Shared last authorship (C Neubauer, DA Ruess).

## Abstract

**Background:** AI-based radiomics has demonstrated promising diagnostic performance for pancreatic cystic neoplasms, yet clinical translation remains limited. Whether this reflects insufficient model performance or structural limitations of the evidence base remains unclear.

**Methods:** We performed a systematic review and diagnostic test accuracy meta-analysis of AI-based radiomics in pancreatic cyst (2015–2025), addressing two clinically relevant tasks (Q1: cyst type differentiation/Q2: malignancy or high-grade dysplasia prediction). Training and validation datasets were synthesized independently using hierarchical models. Study evaluation extended beyond diagnostic performance to a four-dimensional framework integrating RQS 2.0, METRICS, TRIPOD+AI and PROBAST+AI explicitly contrasting pooled diagnostic performance with reporting quality, methodological rigor, and risk of bias. The review was pre-registered (PROSPERO) and conducted according to PRISMA 2020.

**Results:** Twenty-nine studies were included (Q1: n = 15; Q2: n = 14), predominantly retrospective and single center. Training-based analyses showed high apparent diagnostic performance for Q1 (pooled sensitivity/specificity: 0.89 [95% CI, 0.85–0.92]/ 0.90 [0.85–0.93]), but there was substantial heterogeneity (τ² = 0.56/0.78; ρ = 0.38). Validation-based performance remained high (0.86 [0.82–0.89]/ 0.88 [0.81–0.93]), while heterogeneity persisted and prediction regions exceeded confidence regions. Training-based analyses demonstrated similarly high apparent performance (0.88 [0.79–0.95]/0.89 [0.81–0.94]) for Q2, with pronounced heterogeneity (τ² = 1.98/1.61; ρ = 0.63). Validation-based performance was slightly lower, yet still clinically comparable (0.82 [0.75–0.89]/0.86 [0.80–0.91]), and heterogeneity persisted (τ² = 0.71/0.43; ρ = 0.15). Across both tasks, high diagnostic accuracy occurred alongside incomplete reporting, limited validation and an elevated risk of bias.

**Conclusion:** AI-based radiomics for pancreatic cysts has reached a structural performance plateau. Further improvements in diagnostic accuracy alone are insufficient to achieve clinical translation and must be accompanied by a paradigm shift from performance-driven model development toward decision-anchored study designs, robust validation strategies, transparent reporting standard, and clinically integrated evaluation frameworks.

**Summary:** Although pancreatic cystic lesions are increasingly being detected, imaging-based decision-making remains limited, particularly regarding differentiating between cyst types and stratifying malignancy risk. In this PRISMA-compliant and PROSPERO-registered systematic review and meta-analysis of diagnostic tests, we evaluated the use of AI-based radiomics for these two tasks, as well as its contextualized performance. In addition, a four-dimensional framework was employed to conduct the evaluation, incorporating diagnostic accuracy, reporting quality, risk of bias, and radiomics maturity. Across studies published between 2015 and 2025, the pooled diagnostic performance was consistently high, with only modest declines observed from the training to the validation stage. Nevertheless, considerable heterogeneity between studies and limited transportability remained evident. Multidimensional evaluation indicated a systematic dissociation between reported performance and methodological robustness, characterized by incomplete reporting, restricted validation, and an elevated risk of bias. These limitations were consistent across both clinical questions and were not resolved by increasing model complexity. The findings of this meta-analysis suggest that the structural performance of AI-based radiomics for pancreatic cysts has plateaued. To progress towards clinical translation, it is necessary to employ study designs anchored in decision-making processes, robust multi-center validation, and transparent, reproducible evaluation frameworks. This is preferred to further optimization of model architecture alone.

## 1. Introduction

Notwithstanding the rapid advancements being made in the domain of artificial intelligence, the clinical implementation of this technology for the differentiation and treatment of pancreatic cysts appears to be a considerable distance away. For clinicians, two questions predominate in decision-making: what is the nature of the lesion and what is its potential severity? Although typically addressed separately in literature, these represent decisions that are inherently sequential and linked in clinical practice. It is therefore essential to understand their relationship at a meta-analytical level in order to obtain not only performance diagnostics but also a comprehensive picture of the field.

Pancreatic cystic lesions are detected more frequently than they are understood. Cross-sectional imaging reveals cysts incidentally in up to 13% of adults, with a prevalence of cyst occurrence of almost 60% in the 60–70 age group^1,2^. Two fundamental questions continue to challenge diagnostic pathways. Firstly, it is necessary to determine whether a lesion represents a benign serous cyst that can be managed with no intervention or routine observation, or a mucinous cystic lesion (MCN) or intraductal papillary mucinous neoplasm (IPMN) that requires structured surveillance or surgical intervention. Secondly, in lesions selected for surveillance, it is essential to ascertain the true risk of malignant transformation or high-grade dysplasia, and how this risk should guide the intensity and timing of follow-up or intervention^3,4^. Misclassification is costly in both directions, either exposing patients to unnecessary pancreatic surgery at high risk or delaying treatment for high-grade dysplasia or incipient carcinoma^3,5–8^. Among pancreatic cystic neoplasms, IPMN, MCN, serous (SCN) and solid pseudopapillary neoplasm (SPN) account for most entities, with malignant potentials ranging from <5% in SCN to up to 32% in main-duct IPMN^2,9^. New data show that IPMN patients are up to nine times more likely to develop pancreatic cancer than the general population^10^. Even though major guidelines have reduced uncertainty, they are predominantly reliant on retrospective evidence and exhibit limited control for tumor- and patient-specific characteristics. Furthermore, considerable heterogeneity is observed in terms of scope, recommendations for resection versus surveillance, and follow-up^4,11,12^. Even with advanced imaging and endoscopic evaluation, diagnostic uncertainty often remains, leading to surgical resection^2,11^. This clinical dilemma is exacerbated by the persistently poor five-year survival rate of 13% for pancreatic cancer^13^. Consequently, clinical decisions remain individualized and experience-based, perpetuating a diagnostic grey zone.

In this context, radiomics can be defined as an AI-based imaging approach that enables quantitative characterization of imaging biomarkers, not discernible by visual inspection^14,15^. The application of this technique to pancreatic cystic lesions has shown its potential to support the classification of cyst subtypes and the estimation of malignancy risk. Aiming to reduce subjectivity and inter-observer variability, particularly in borderline or indeterminate cases with reproducible, data-driven imaging signatures^16^. The number of publications on AI-based radiomics in pancreatic cysts has increased significantly since 2015, mainly due to cohorts from individual centers reporting promising results in both distinguishing cysts and predicting malignancy^16,17^. Despite growing interest in end-to-end deep learning, radiomics remains particularly relevant for cystic lesions of the pancreas due to its modular and partially interpretable architecture, which fulfills clinical requirements for transparency and methodological verifiability. However, many studies exhibit recurring limitations, including heterogeneous imaging protocols, limited sample sizes, extensive and insufficiently transparent feature selection, reliance on internal validation often without independent internal validation cohorts and infrequent external testing, collectively impeding transferability to clinical practice^18^.

Although recent reviews report good diagnostic accuracy, they rarely include a comprehensive quality assessment or consider the entire clinical pathway^16,17,19^. This results in a fragmented, largely descriptive picture that reflects the structure of the underlying evidence rather than the shortcomings of individual reviews. Consequently, the accelerated development of medical AI applications for pancreatic cysts requires an analysis that examines interrelated clinical tasks sequentially, evaluates diagnostic accuracy and transferability, and incorporates benchmarks for quality assessment. This may provide a more holistic picture and support the transfer of these applications into clinical practice.

To achieve this objective, a four-dimensional assessment framework was employed, integrating AI-based radiomics diagnostic performance, bias potential, reporting quality, and compliance with radiomics-specific methodological requirements. These factors were then directly linked to meta-analytical results^20–23^. The analysis addresses two clinically sequential questions: (Q1) the differentiation between serous and mucinous cystic neoplasms and (Q2) the prediction of malignancy in mucinous and IPMN lesions, thereby delineating the boundary between experimental potential and clinical translation.

## 2. Methods

### 2.1 Study design and eligibility

This systematic review and meta-analysis were conducted in accordance with the PRISMA 2020 statement. A predefined protocol specified the research questions, eligibility criteria, data-extraction items, and statistical analysis plan, and was prospectively registered in PROSPERO (CRD420251137552). Eligible studies developed or validated AI-based radiomics models using CT or MRI data. Both handcrafted radiomics combined with machine-learning classifiers and hybrid deep-learning architecture based on imaging-derived features were included. Studies were required to report the area under the receiver operating characteristic curve (AUC) together with corresponding sensitivity and specificity to allow quantitative synthesis. Studies reporting accuracy alone were excluded, as accuracy is threshold-dependent, influenced by disease prevalence, and therefore unsuitable for meta-analytic pooling and inconsistent with reporting and bias-assessment standards. Considering the anticipated heterogeneity in modelling pipelines, validation strategies, and reporting quality, the analysis was planned *a priori* to integrate pooled diagnostic performance with structured assessments of reporting quality, risk of bias, and radiomics standards using the Radiomics Quality Score 2.0 (RQS 2.0), the Transparent Reporting of a multivariable prediction model for Individual Prognosis Or Diagnosis plus Artificial Intelligence (TRIPOD+AI), the Prediction Model Risk of Bias Assessment Tool plus Artificial Intelligence (PROBAST+AI), and the methodological radiomics score (METRICS).

### 2.2 Literature search and selection

A systematic literature search was conducted in PubMed, Embase, IEEE Xplore and Google Scholar to identify relevant studies published between January 2015 and July 2025. The search strategy combined terms relating to radiomics, artificial intelligence, machine learning, deep learning, pancreatic disease and neoplasms, and was adapted to the syntax of each database. IEEE Xplore was included to capture early technical developments. Google Scholar was also searched to enhance the identification of interdisciplinary AI-based radiomics studies and to support the citation-based identification of relevant literature. The complete search strategies for all databases are provided in the supplementary information. Only peer-reviewed articles published in English and indexed in the Journal Citation Reports were considered, to ensure adherence to peer-review standards and enable data traceability. Conference abstracts, reviews and studies published in non-indexed journals were excluded to ensure data quality and reproducibility. All records were imported into a reference management system and de-duplicated according to predefined eligibility criteria. Two reviewers independently screened titles and abstracts, followed by a full-text assessment of potentially eligible studies. Discrepancies were resolved by consensus. The study selection process adhered to the PRISMA 2020 guidelines and is illustrated in Table 1.

**Table 1.** Characteristics of studies included in the meta-analysis. Fifteen studies addressed cyst type differentiation (Q1) and fourteen studies addressed malignancy or high-grade dysplasia prediction in mucinous and IPMN lesions (Q2). Expanded study characteristics are provided in the Supplementary Information. Not reported / NA indicates that values were not reported in the original study or could not be computed due to insufficient data.

| Study | Country | Design | N [groups] | Modality | Segmentation | Reference standards | Validation | Best Performance Training, AUC [CI] | Best Performance, Validation, AUC [CI] |
| --- | --- | --- | --- | --- | --- | --- | --- | --- | --- |
| **(Q1), n= 15** |  |  |  |  |  |  |  |  |  |
| Wei et al. (2019)<sup>30</sup> | China | Retrospective, monocentric | 260 [SCN=102, non-SCN=158] | CT | manual | Histopathology | Internal | 0.77 [0.76-0.77] | 0.84 [0.74-0.94] |
| Yang J et al. (2019)<sup>31</sup> | China | Retrospective, monocentric | 78 [SCN=53, non-SCN=25] | CT | manual | Histopathology | Internal | 0.77 [0.64-0.90] | 0.66 [0.35-0.97] |
| Xie H et al. (2020)<sup>32</sup> | China | Retrospective, monocentric | 57 [SCN=26, non-SCN=31] | CT | manual | Histopathology | Internal Cross | 0.99 [0.97-1.00] | Not reported |
| Chen HY et al. (2021)<sup>33</sup> | China | Retrospective, multicentric | 100 [SCN=57, non-SCN=43] | CT | manual | Histopathology | External | 0.93 [0.84-0.98] | 0.89 [0.72-0.97] |
| Chen Sh et al. (2021)<sup>34</sup> | China | Retrospective, monocentric | 89 [SCN=31, non-SCN=58] | CT | manual | Histopathology | Internal | 0.97 [0.910-1.00] | 0.87 [0.65–0.98] |
| Gao et al. (2021)<sup>35</sup> | China | Retrospective, monocentric | 170 [SCN=115, non-SCN=55] | CT | manual | Histopathology | Internal | 0.91 [0.84-0.97] | 0.90 [0.81-1.00] |
| Xie T et al. (2021)<sup>36</sup> | China | Retrospective, monocentric | 216 [SCN=113, non-SCN=103] | CT | semi-automated | Histopathology | Internal Cross | 0.78 [0.72-0.84] | Not reported |
| Awe et al. (2022)<sup>37</sup> | USA | Retrospective, monocentric | 99 [SCN=19, non-SCN=80] | CT | semi-automated | Histopathology | Internal Cross | Not reported | 0.73 [0.62-0.84] |
| Dong et al. (2022)<sup>38</sup> | China | Retrospective, monocentric | 129 [SCN=49, non-SCN=80] | CT | semi-automated | Histopathology | Internal | 0.87 [0.80-0.94] | 0.85 [0.72-0.98] |
| Yang R et al. (2022)<sup>39</sup> | China | Retrospective, monocentric | 110 [SCN=63, non-SCN=47] | CT | manual | Histopathology | None | 0.97 [0.93-1.00] | Not reported |
| Liang et al. (2022)<sup>40</sup> | China | Retrospective, monocentric | 193 [SCN=99, non-SCN=94] | CT | manual | Histopathology | Internal | 0.97 [0.95-1.00] | Not reported |
| Chu et al. (2022)<sup>41</sup> | USA | Retrospective, monocentric | 214 [SCN=60, non-SCN=97] | CT | manual | Histopathology | Internal Cross | 0.94 [0.90-0.98] | Not reported |
| Fang et al. (2023)<sup>42</sup> | China | Retrospective, monocentric | 304 [SCN=216, non-SCN=88] | MRI | manual | Histopathology | Internal | 0.93 [0.90-0.96] | 0.86 [0.75-0.96] |
| Ansari et al. (2024)<sup>43</sup> | USA | Retrospective, monocentric | 136 [SCN=49, non-SCN=87] | MRI | manual | EUS FNA Histology | External | 0.98 [0.96-1.00] | 0.90 [Not reported/NA] |
| Torra-Ferrer et al. (2025)<sup>44</sup> | Spain | Retrospective, multicentric | 261 [SCN=90, non-SCN=171] | CT | manual | Histopathology | External | 0.90 [0.85-0.95] | 0.93 [0.87-0.99] |
| **(Q2), n= 14** |  |  |  |  |  |  |  |  |  |
| Hanania et al. (2016)<sup>45</sup> | USA | Retrospective, monocentric | 53 [benign=19, malignant/HGD=34] | CT | manual | Histopathology | Internal Cross | 0.96 [0.92-0.99] | Not reported |
| Permuth et al. (2016)<sup>46</sup> | USA | Retrospective, monocentric | 38 [benign=20, malignant/HGD=18] | CT | semi-automated | Histopathology | Internal Cross | 0.87 [0.84-0.89] | Not reported |
| Chakraborty et al. (2018)<sup>47</sup> | USA | Retrospective, monocentric | 103 [benign=76, malignant/HGD=27] | CT | manual | Histopathology | Internal Cross | 0.81 [0.70-0.92] | Not reported |
| Polk et al. (2020)<sup>48</sup> | USA | Retrospective, monocentric | 51 [benign=22, malignant/HGD=29] | CT | semi-automated | Histopathology | Internal Cross | 0.93 [0.85-1.00] | Not reported |
| Tobaly et al. (2020)<sup>49</sup> | France | Retrospective, multicentric | 408 [benign=181, malignant/HGD=227] | CT | semi-automated | Histopathology | External | 0.83 [0.78-0.87] | 0.71 [0.66-0.84] |
| Harrington et al. (2020)<sup>50</sup> | USA | Retrospective, monocentric | 33 [benign=26, malignant/HGD=7] | CT | manual | Histopathology | Not reported | 0.98 [0.91-1.00] | Not reported |
| Li et al (2021)<sup>51</sup> | China | Retrospective, monocentric | 107 [benign=71, malignant/HGD=36] | CT | manual | Histopathology | Internal | 0.83 [0.73 – 0.93] | 0.92 [0.78 – 1.00] |
| Cui et al. (2021)<sup>52</sup> | China | Retrospective, multicentric | 202 [benign=152, malignant/HGD=50] | MRI | manual | Histopathology | External | 0.90 [0.83-0.95] | 0.88 [0.75-0.95] |
| Cheng Sh et al. (2022)<sup>53</sup> | China | Retrospective, monocentric | 60 [benign=37, malignant/HGD=23] | CT | manual | Histopathology | Internal Cross | 0.86 [0.77-0.96] | Not reported |
|  | China | Retrospective, monocentric | 60 [benign=37, malignant/HGD=23] | MRI | manual | Histopathology | Internal Cross | 0.94 [0.88-1.00] | Not reported |
| Wang et al. (2022)<sup>54</sup> | China | Retrospective, multicentric | 363 [benign=218, malignant/HGD=150] | CT | manual | Histopathology | External | 0.88 [0.78-0.98] | 0.77 [0.64-0.90] |
| Flammia et al. (2023)<sup>55</sup> | Italy | Retrospective, monocentric | 50 [benign=31, malignant/HGD=19] | MRI | manual | Fukoka Guidelines | Internal | 0.98 [0.94-1.00] | Not reported |
| Lee et al. (2024)<sup>56</sup> | Korea | Retrospective, monocentric | 194 [benign=83, malignant/HGD=111] | CT | semi-automated | Histopathology | Internal | 0.85 [0.79-0.91] | 0.85 [0.75-0.96] |
| Lou et al. (2024)<sup>57</sup> | China | Retrospective, monocentric | 121 [benign=61, malignant/HGD=60] | CT | manual | Histopathology | Internal | 0.93 [0.88-0.99] | 0.90 [0.75-0.96] |
| Cheng Si et al. (2025)<sup>58</sup> | China | Retrospective, multicentric | 362 [benign=214, malignant/HGD=148] | CT | manual | Histopathology | External | 1.00 [1.00-1.00] | 0.96 [0.93-1.00] |

### 2.3 Data extraction

Two reviewers independently extracted data using a predefined, structured template. From each study, baseline data on study design and setup, imaging acquisition and image processing, model development and validation, as well as the best-performing results from the training and validation datasets were extracted. The complete overview of additional extracted variables can be found in Supplementary Information (Table S1). Only studies that reported the area under the receiver operating characteristic curve (AUC), together with the corresponding sensitivity and specificity, were eligible for quantitative synthesis. Where multiple models or validation cohorts were reported, the data were extracted in a hierarchical manner to preserve comparability between the development and validation stages. Priority was given to the most external validation dataset, where available. All entries were verified by consensus, and any potential overlap in cohorts was addressed by retaining the most comprehensive dataset. This database was then used as the basis for the quantitative meta-analyses and four-dimensional quality assessments.

### 2.4 Statistical analysis

#### 2.4.1 Diagnostic test accuracy meta-analysis (DTA-MA)

DTA-MA was conducted to evaluate the performance of AI-based radiomics in:

(Q1) cyst type differentiation between serous and mucinous pancreatic cystic lesions (SCN vs MCN/IPMN)
(Q2) malignancy or high-grade dysplasia prediction in mucinous and IPMN lesions.

Across both research questions, the included studies reported the sensitivity and specificity of AI-based radiomics models using separate training and validation datasets. Accordingly, our meta-analysis presents results separately for training and validation datasets.

The training and validation datasets were separated in advance, and this approach was applied consistently throughout all analyses. Subsequent results refer to this framework without reiterating the rationale behind it. For studies that did not implement cross-validation, we incorporated their reported estimates of sensitivity and specificity into the analysis of the training dataset. These studies usually provided one pair of diagnostic accuracy estimates based on their total sample size without distinguishing between the training and validation subsets. As training datasets are generally larger than validation datasets, we considered estimates from non-cross-validated studies to be most comparable to those derived from training datasets in cross-validated studies.

To perform the DTA-MA for validation datasets, we used the sensitivity and specificity estimates explicitly reported for validation datasets in cross-validated studies and the overall sensitivity and specificity estimates from the non-cross validated studies. This pragmatic assignment does not imply equivalence between training and validation performance. Instead, it represents methodological approximation, the influence of which was explicitly assessed in sensitivity analyses. Because validation estimates from studies addressing (Q2) were also incorporated into the training-based DTA-MA, we performed a sensitivity analysis excluding these studies to avoid potential double contribution and to assess the robustness of the pooled estimates. This approach enabled us to assess the influence of this methodological choice on the robustness of our results. All DTA-MA were conducted within a Bayesian framework using a bivariate hierarchical model which is also recommended from the Cochrane handbook^24,25^. In hierarchical summary receiver operating characteristic (HSROC) plots, confidence regions summarize uncertainty around the pooled sensitivity and specificity estimates, whereas prediction regions reflect between-study heterogeneity and expected variability in future study settings. Where confidence intervals were not reported, 95% confidence intervals were derived using standard statistical methods based on the reported point estimates and sample sizes, where applicable.

#### 2.4.2 Subgroup analysis

To explore potential sources of heterogeneity, prespecified subgroup analyses were conducted separately for Q1 and Q2, as well as for the training and validation datasets. These analyses were based on the following factors: study design (monocentric vs. multicentric); validation strategy; imaging modality; segmentation approach; preprocessing; model category (handcrafted, hybrid, or deep-learning–based radiomics); inclusion of non-radiomics features; geographic region; and year of publication. Subgroup-specific pooled sensitivity and specificity estimates were calculated using the same Bayesian bivariate hierarchical modelling framework as in the primary analyses. Due to the limited number of studies within several subgroups, no formal adjustment for multiple comparisons was applied and the subgroup analyses were considered exploratory in nature. A comprehensive tabular representation was employed to ensure optimal transparency, in which the detailed subgroup results were summarized.

#### 2.4.3 Study inclusion for performance summaries

The inclusion criteria for meta-analyses and performance–quality spider plots were restricted to studies reporting Area Under the Curve (AUC), sensitivity, and specificity for the respective dataset. Studies reporting complete metrics exclusively for validation were included in validation analyses but excluded from training-based summaries. Conversely, all eligible studies were included in quality assessments based on RQS 2.0, METRICS, TRIPOD+AI, and PROBAST+AI, irrespective of performance reporting completeness.

#### 2.4.4 Inter-rater agreement

Methodological and reporting quality assessments were independently performed by two reviewers, and inter-rater agreement was quantified using weighted Cohen’s kappa^26^. Disagreements were weighted according to the squared distance from perfect agreement, with larger discrepancies between ratings being penalized more heavily. Weighted Cohen’s kappa values were calculated separately for each assessment tool and for both research questions, where applicable. Agreement levels were interpreted using conventional qualitative categories (e.g. moderate, substantial or almost perfect agreement). This analysis was performed to assess the consistency and reliability of study evaluations across different quality assessment frameworks. All analyses were performed in R (version 4.5.2).

### 2.5 Quality assessment framework

Each study underwent a structured evaluation across four complementary dimensions. RQS 2.0 and METRICS were used to capture different aspects of radiomics methodology^22,23^. Reporting transparency was assessed using TRIPOD+AI, while clinical risk of bias and applicability were evaluated using PROBAST+AI^20,21^. RQS 2.0 conceptualizes the stepwise maturation of radiomics models from exploratory analysis to clinical deployment, adapted to the architectural framework in terms of handcrafted, deep learning or hybrid radiomics^22^. Early levels (RRL1–3) cover foundational exploration, data preparation, and prototype model development, whereas intermediate levels (RRL4–5) focus on internal validation and capability testing. Higher-order levels (RRL6–9) require evidence of trustworthiness, prospective validity, real-world applicability, and sustained clinical deployment, representing the transition from technical feasibility to true clinical integration. All studies were additionally re-evaluated using RQS 1.0, which served as methodological baseline control.

METRICS was used as a complementary radiomics-specific assessment for reproducibility and methodological rigor^27,28^. Domain-level scores were calculated as the ratio of achieved to maximal weighted points per domain and normalized to percentages (0–100%), preserving the original weighting while enabling interpretable domain-specific comparisons.

Reporting transparency was assessed using the TRIPOD+AI checklist. Each item was scored as 1 (fulfilled), 0.5 (partially fulfilled), or 0 (not fulfilled).

Risks of bias and applicability were evaluated using PROBAST+AI. Items were scored as yes (1), probably yes (0.75), probably no (0.25), no (0) or no information (0). Risks of bias and applicability concerns were graded as low, unclear or high. Scores were calculated at the domain level and separately for model development and evaluation phases to enable structured comparison and visualization of methodological weaknesses. QUADAS-2 was not applied because it was developed for classical diagnostic accuracy studies and an official AI-adapted extension was not available at the time the included studies were conducted or this review was designed^29^.

Each study included in the analysis was subjected to four structured assessments, resulting in a comprehensive item-level evaluation (174 items per study; 5046 items per reviewer). All assessments were performed independently by two reviewers (a visceral surgeon and a senior board-certified radiologist with expertise in abdominal imaging). Discrepancies were resolved by consensus. If required, adjudication was performed by a senior board-certified pancreatic surgeon.

### 2.6 Reporting and reproducibility

This work was conducted in accordance with the PRISMA 2020 guidelines. The complete checklists and detailed search strategies can be found in the Supplementary Information. All analyses and figures were generated using reproducible R code. The code used for data processing, analysis and figure generation can be obtained from the corresponding author upon request. Quality assessment frameworks and scoring rules were applied consistently across all included studies as predefined in the Methods. Domain-level results for all assessments, including independent ratings by both reviewers, are provided in the Supplementary Information.

## 3. Results

### 3.1 Study selection and characteristics

The study selection process is summarized in the PRISMA flowchart (Figure 1). A total of 1.974 records were identified through database searches. After removal of duplicates and screening of titles and abstracts, full-text articles were assessed for eligibility, resulting in the inclusion of 29 studies in the final analysis. Of these, 15 studies addressed cyst type differentiation between serous and mucinous pancreatic cystic lesions (SCN vs. MCN/IPMN; Q1), and 14 studies evaluated malignancy or high-grade dysplasia prediction in mucinous and IPMN lesions (Q2).

**Figure 1.**
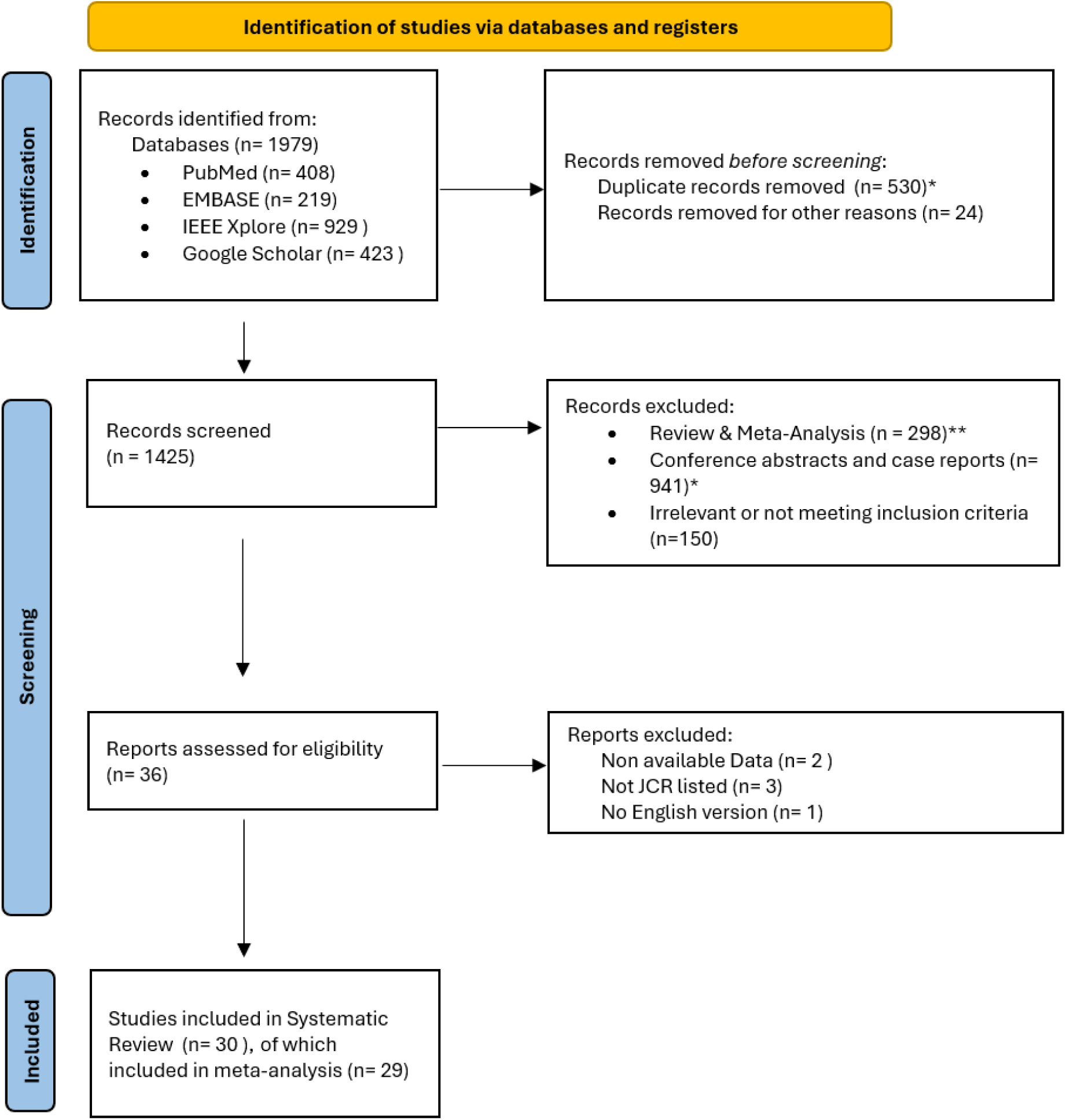
PRISMA 2020 flowchart. Study selection for the systematic review and meta-analysis of AI-based radiomics for pancreatic cystic lesions. Records were identified through database searches (PubMed, Embase, IEEE Xplore, and Google Scholar). Duplicate records were identified and removed using reference management software prior to screening (*).

Included studies were published between 2015 and 2025 and originated predominantly from East Asia, followed by North America and Europe. Most studies employed retrospective, monocentric designs, with only a minority using multicentric cohorts. Computed tomography was the most frequently used imaging modality, followed by magnetic resonance imaging; only a small number of studies combined both modalities. Reference standards primarily consisted of histopathological evaluation of surgical specimens, supplemented by clinical follow-up in selected studies. In one study, the reference standard relied on follow-up alone. Key study characteristics for Q1 and Q2 are summarized in Table 1. Additional methodological details and expanded study-level characteristics are provided in the Supplementary Information (Tables S1 and S2).

### 3.2 Diagnostic test accuracy meta-analysis

#### 3.2.1 Diagnostic accuracy for cyst type differentiation (Q1)

Training-based analyses across 14 studies (median sample size per study: 153) demonstrated high apparent diagnostic performance. Pooled sensitivity and specificity were 0.89 [0.85, 0.92] and 0.90 [0.85, 0.93], respectively. Between-study heterogeneity (τ²) was substantial for both sensitivity (0.56) and specificity (0.78), with a moderate correlation between the two (ρ = 0.38). A comparison of the confidence and prediction regions in the SROC space also indicated significant variability between studies.

Although exploratory subgroup analyses did not identify any statistically significant differences across the prespecified variables, several subgroups comprised only one or two studies, which limited statistical power. Training-based forest plots, SROC analyses and subgroup results can be found in the supplementary information (Figures S1–S2; Table S3).

Validation-based analyses revealed a slightly lower, yet still clinically comparable level of diagnostic performance. The pooled sensitivity and specificity were 0.86 [0.82, 0.89] and 0.88 [0.81, 0.93], respectively (Figure 2). Exclusion of non–cross-validated studies yielded pooled sensitivity and specificity estimates of 0.81 [0.73, 0.88] and 0.84 [0.78, 0.89], demonstrating robustness of the primary analysis with substantial overlap of confidence intervals (Figure S3). The heterogeneity estimates for the validation dataset were 0.34 for sensitivity and 1.26 for specificity, indicating a moderate correlation between sensitivity and specificity (ρ = 0.44). SROC analysis again demonstrated a substantially larger prediction region compared with the confidence region, consistent with marked between-study heterogeneity (Figure 3). In validation-based exploratory subgroup analyses, no consistent differences were observed across most variables. However, segmentation strategy was associated with significant differences in specificity. Nevertheless, subgroup sizes were small, so the findings should be interpreted cautiously (Table S4).

**Figure 2.**
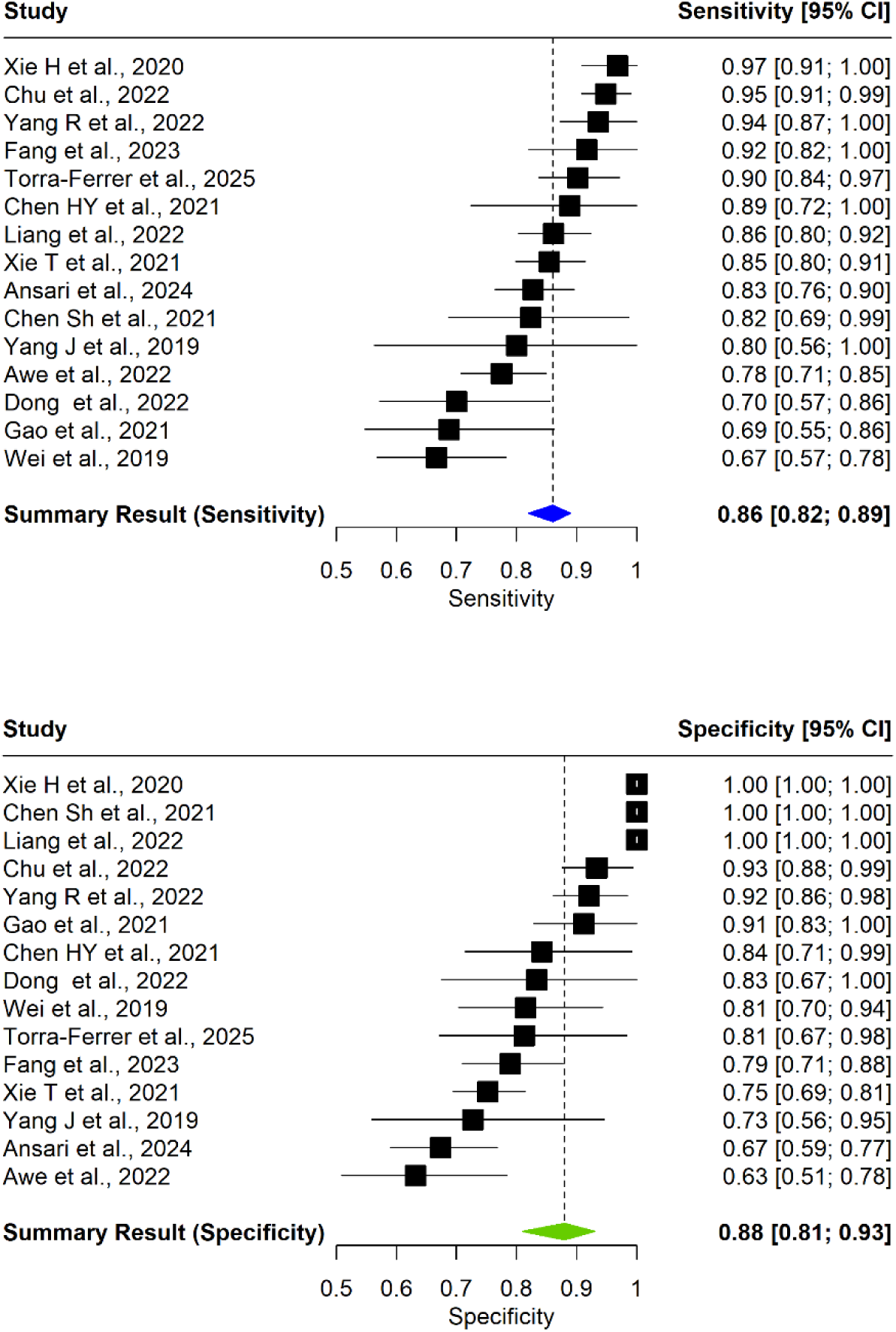
Sensitivity and Specificity summary results for the Q1 validation dataset. Studies that do not report sensitivity and specificity for the training dataset are not included in training-based analyses.

**Figure 3.**
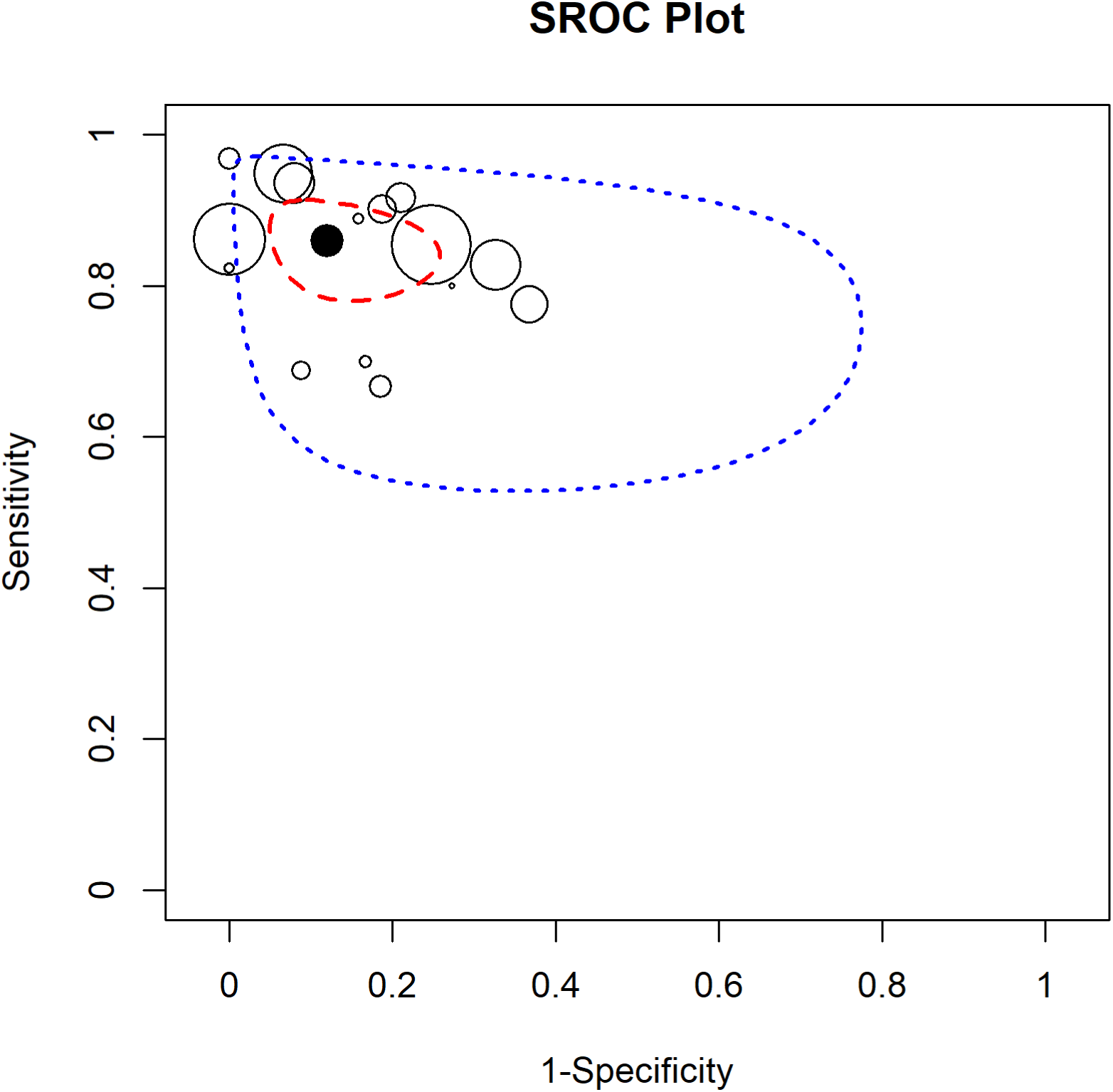
SROC plot in terms of the validation dataset for Q1. The unfilled points represent pairs of sensitivity and 1-specificity estimates for each study. The black filled point represents the pair of summary sensitivity and specificity estimates. The region shown by the red dashed line represents the confidence region, and the region shown by the blue dotted line represents the prediction region.

#### 3.2.2 Diagnostic accuracy for malignancy or high-grade dysplasia prediction (Q2)

Across 12 studies, with a median total sample size of 105 participants, training-based analyses similarly demonstrated a high apparent diagnostic performance. The pooled sensitivity and specificity were 0.88 [0.79, 0.95] and 0.89 [0.81, 0.94], respectively. The heterogeneity estimates for sensitivity and specificity were 1.98 and 1.61, indicating substantial heterogeneity between studies. A moderate to strong correlation was found between sensitivity and specificity (*ρ =* 0.63). A comparison of confidence and prediction regions in SROC space provided further support for marked heterogeneity. Exploratory subgroup analyses revealed no statistically significant differences across the prespecified variables. Despite this, several subgroups comprised only one or two studies, which limited statistical power. Training-based forest plots, SROC analyses and subgroup results are provided in the Supplementary Information (Figures S4–S5 ; Table S5).

Validation-based analyses showed slightly lower but clinically comparable diagnostic performance. The pooled sensitivity and specificity were 0.82 [0.75, 0.89] and 0.86 [0.80, 0.91], respectively (Figure 4). A sensitivity analysis excluding non–cross-validated studies yielded pooled estimates of 0.73 [0.64, 0.83] for sensitivity and 0.86 [0.76, 0.94] for specificity, demonstrating robustness of the primary analysis with substantial overlap of confidence intervals (Figure S6). Heterogeneity estimates for the validation dataset were 0.71 for sensitivity and 0.43 for specificity, with a low correlation between sensitivity and specificity (*ρ =* 0.15). SROC analysis again demonstrated a substantially larger prediction region compared with the confidence region, consistent with marked between-study heterogeneity (Figure 5). In validation-based exploratory subgroup analyses, we observed no consistent differences across most variables; regional differences in sensitivity emerged, although subgroup sizes were small and we should interpret findings cautiously (Table S6).

**Figure 4.**
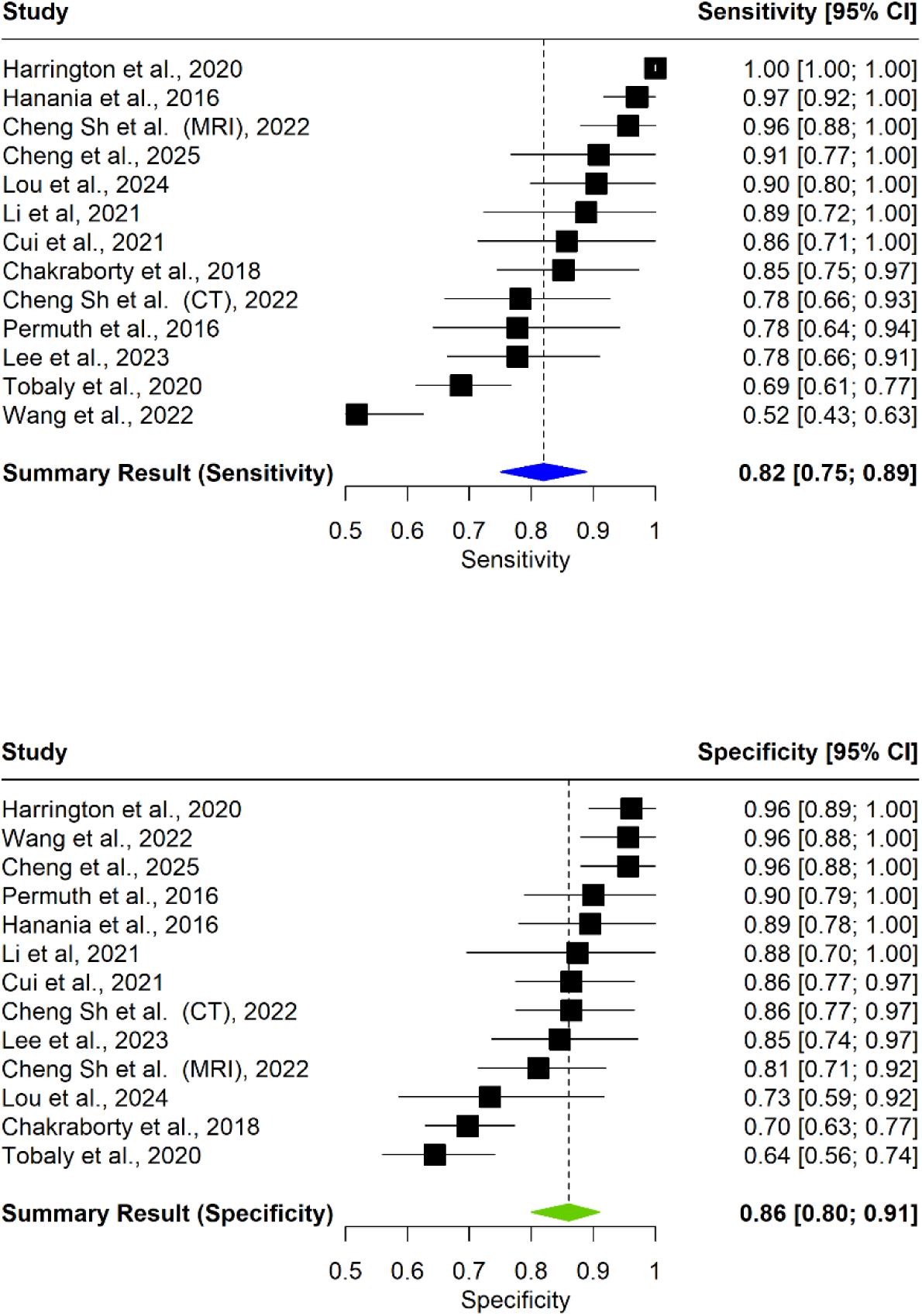
Sensitivity and Specificity summary results for the Q2 validation dataset. Studies not reporting sensitivity and specificity for the training dataset are not included in training-based analyses.

**Figure 5.**
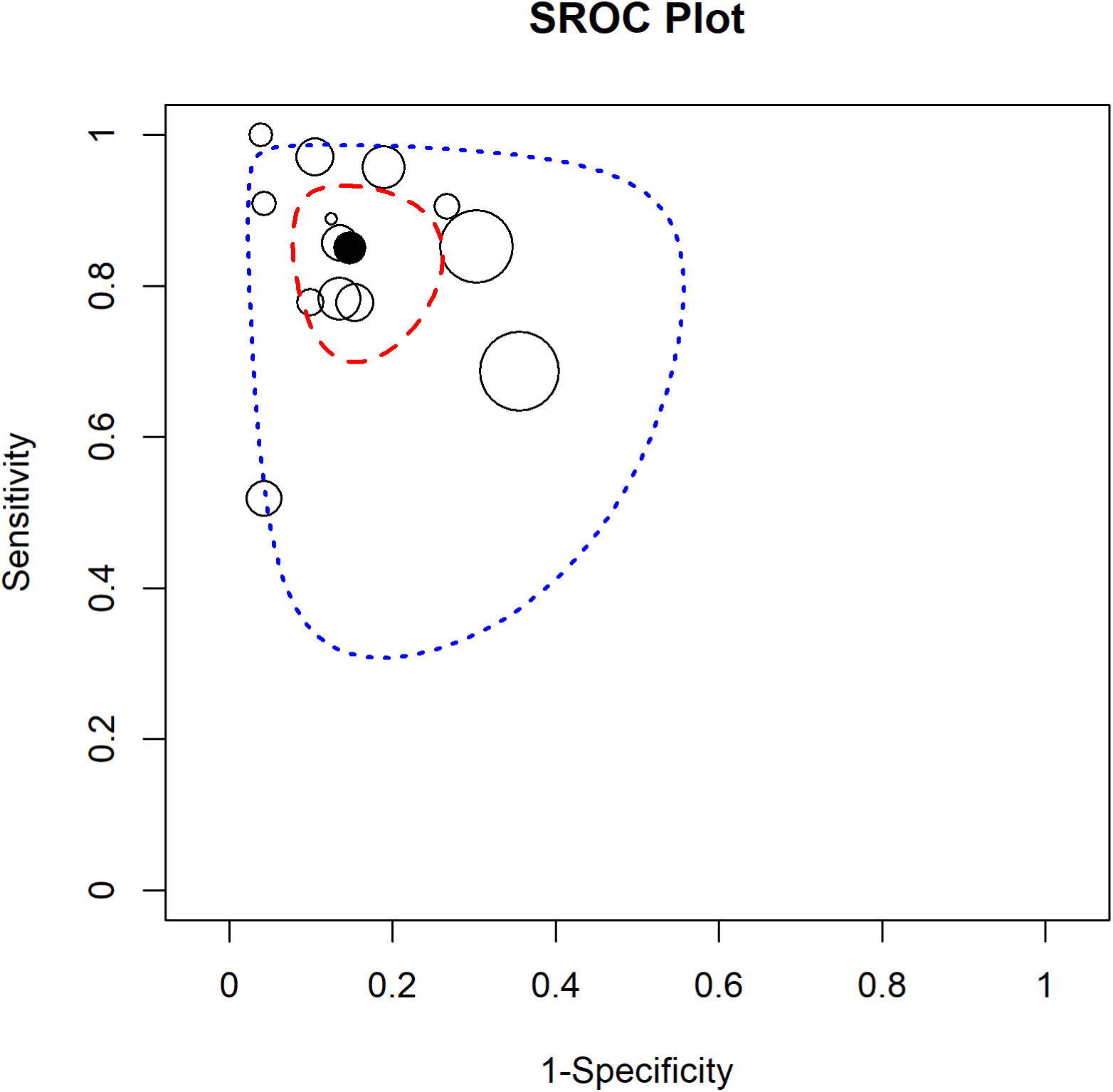
SROC plot in terms of the validation dataset for Q2. The unfilled points represent pairs of sensitivity and 1-specificity estimates for each study. The black filled point represents the pair of summary sensitivity and specificity estimates. The region shown by the red dashed line represents the confidence region, and the region shown by the blue dotted line represents the prediction region

### 3.3 Quality, risk of bias and radiomics-specific assessments

#### 3.3.1 Assessments for cyst type differentiation (Q1)

METRICS analysis revealed marked heterogeneity in methodological completeness. While domains related to study design and feature processing achieved comparatively high mean adherence (≥75%), substantial shortcomings were observed in imaging data, segmentation, image processing, and feature extraction, with fulfilment rates at or below 50% during the early stages of the radiomics pipeline. At the end of the workflow, only 33% of items were satisfied in the testing domain, and no items were fulfilled in the open science domain (Figure S7; Table S7).

RQS 2.0 scores increased cumulatively across early Radiomics Readiness Levels (RRLs), with the median score rising from 4 points at RRL1 to 15 points at RRL5. Beyond this intermediate stage, no further increase in the median cumulative score was observed, despite a continued rise in the maximum achievable score from 34 points at RRL5 to 56 points at RRL9. In detail, RRL1 (Foundational Exploration), RRL2 (Data Preparation), and RRL3 (Prototype Model Development) showed fulfilment rates just above 50%, whereas RRL4 (Internal Validation) and RRL5 (Capability Testing) declined to approximately 25%. All higher readiness levels, RRL6 (Trustworthiness Assessment), RRL7 (Prospective Validity), RRL8 (Applicability and Sustainability), and RRL9 (Clinical Deployment), showed 0% fulfilment across the assessed studies (Figure S9; Table S8). These findings were concordant with the RQS 1.0 assessment, which identified the same structural weaknesses, albeit with lower granularity (Table S9).

TRIPOD+AI assessment revealed a heterogeneous reporting profile across domains (Figure 6A; Table S10). Complete reporting was consistently achieved for the title and abstract domains (100% each). Reporting quality declined across subsequent manuscript sections, with mean fulfilment rates of 78.9% for the introduction and 74.5% for the methods domain. Domains directly relevant to transparency and clinical accountability showed substantially lower adherence. Open science items were fulfilled in only 53.1% of cases, while patient and public involvement demonstrated the lowest overall adherence at 46.9%. Reporting of results and discussion sections reached 56.6% and 71.9% fulfilment, respectively, indicating moderate completeness but marked variability across studies. Inspection of individual study-level data confirmed pronounced heterogeneity within these domains, with fulfilment rates ranging widely between studies, particularly for open science practices, patient and public involvement, and results reporting (Table S10). Overall, no domain beyond the title and abstract achieved uniformly high adherence across studies, indicating that reporting quality remained inconsistent even in core methodological and interpretative sections.

**Figure 6A.**
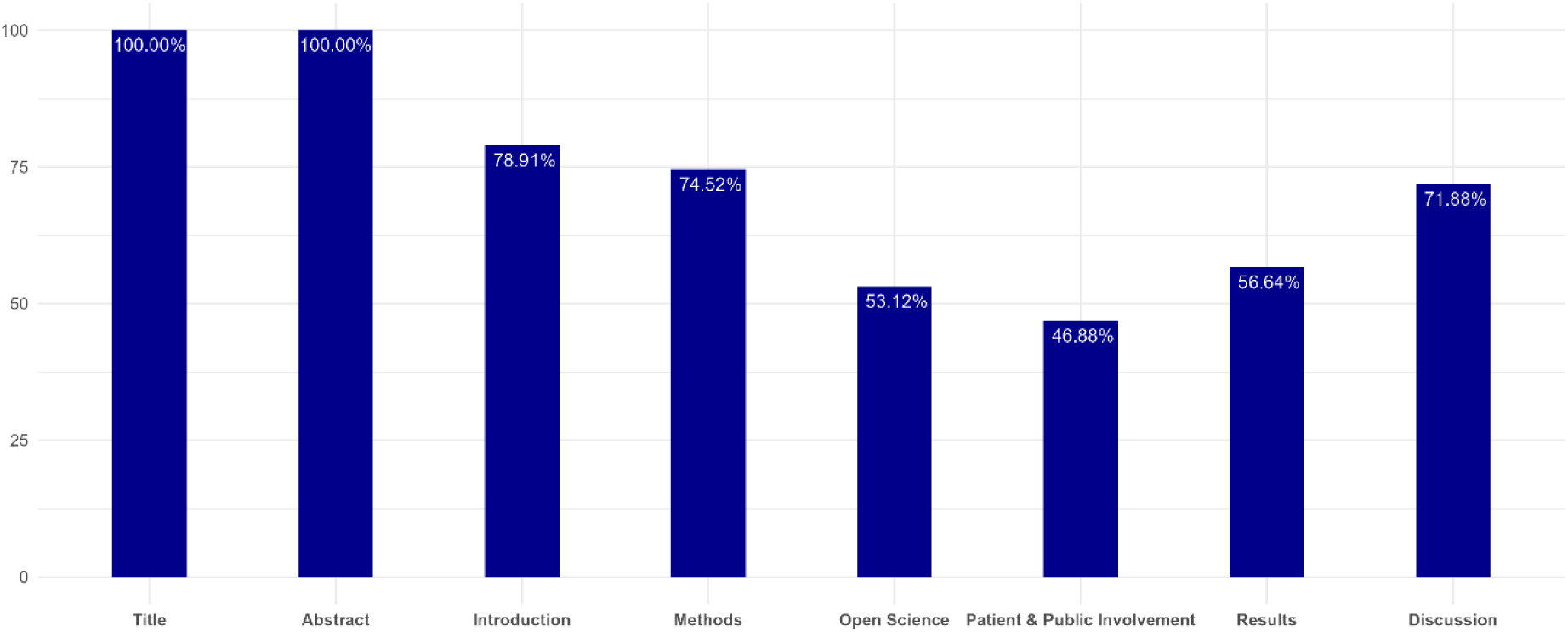
TRIPOD+AI for studies addressing Q1. Domain-level reporting completeness is shown as mean percentages across included studies. Detailed domain ratings are in the supplementary information.

PROBAST+AI assessment identified a consistent imbalance between low applicability concerns and elevated risk-of-bias across studies (Figure 6B; Table S11). At the domain level, risk of bias related to participant selection was consistently rated as low across studies, reflecting well-defined inclusion criteria and clinically plausible target populations. In contrast, bias related to predictors was frequently rated as unclear or high, most commonly due to insufficient reporting of feature selection procedures, handling of correlated radiomics features, and lack of safeguards against information leakage between training and validation datasets. The analysis domain showed the highest concentration of high risk-of-bias ratings. Common deficiencies included inadequate handling of overfitting, limited use of appropriate internal validation strategies, and insufficient reporting of model calibration and uncertainty. Outcome-related risk of bias was predominantly rated as unclear, driven by incomplete reporting of reference standards, variable definitions of malignancy or high-grade dysplasia, and limited transparency regarding outcome adjudication procedures. While overall applicability was generally rated as low across domains, overall risk-of-bias judgements were predominantly unclear or high.

**Figure 6B.**
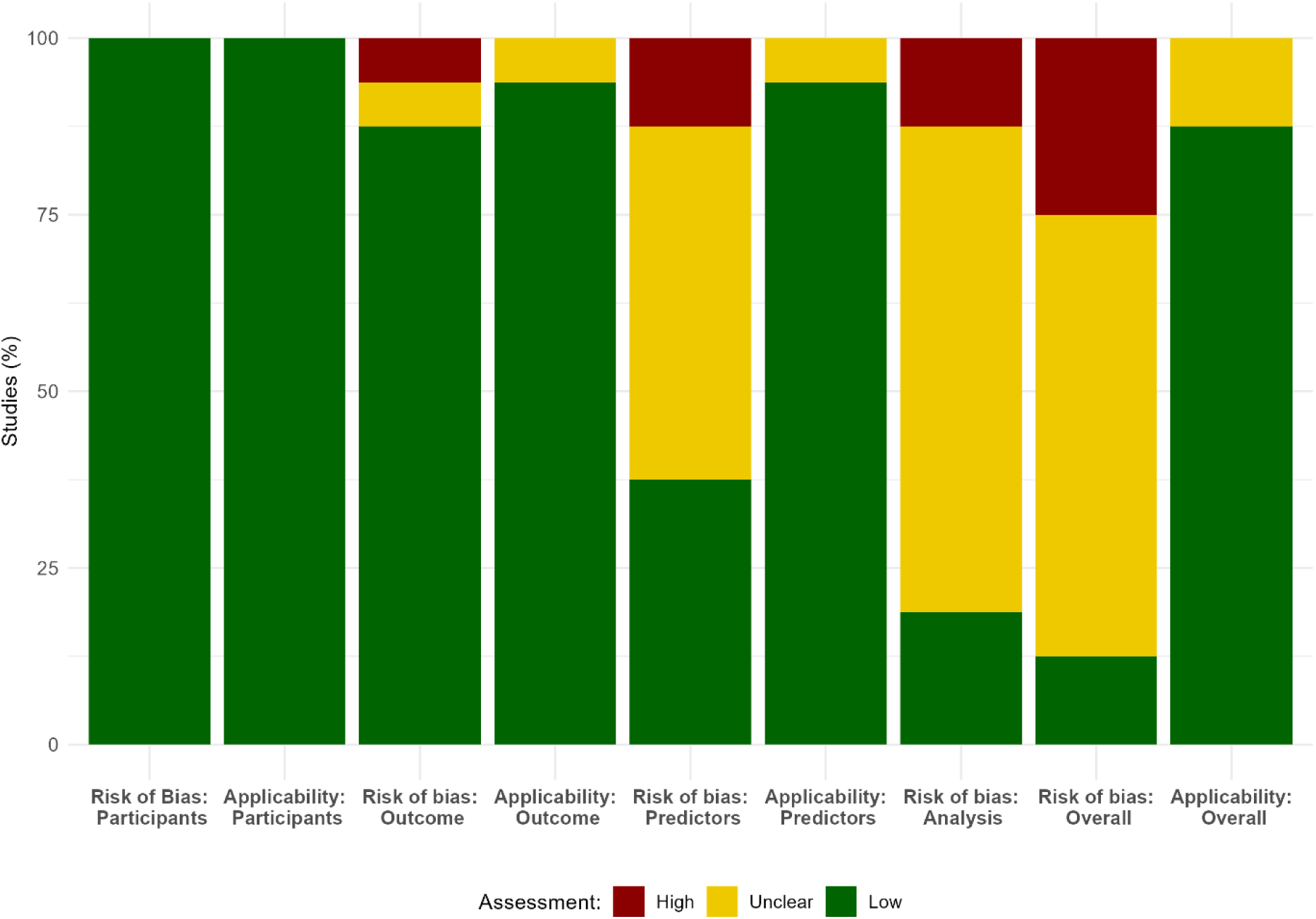
PROBAST+AI for studies addressing Q1. Domain-level distributions of risk of bias are shown as the percentage of studies classified as low, unclear, or high risk for each domain. Detailed domain ratings are provided in the Supplementary Information.

Multidimensional spider plot analysis demonstrated a consistent dissociation between diagnostic performance and methodological robustness across studies. High performance metrics (AUC, sensitivity, specificity), observed in both training and validation datasets, were frequently accompanied by lower scores for radiomics-specific assessments (RQS 2.0 and METRICS), reporting quality (TRIPOD+AI), and risk of bias (PROBAST+AI). This pattern was consistent across individual studies (Figure 7).

**Figure 7.**
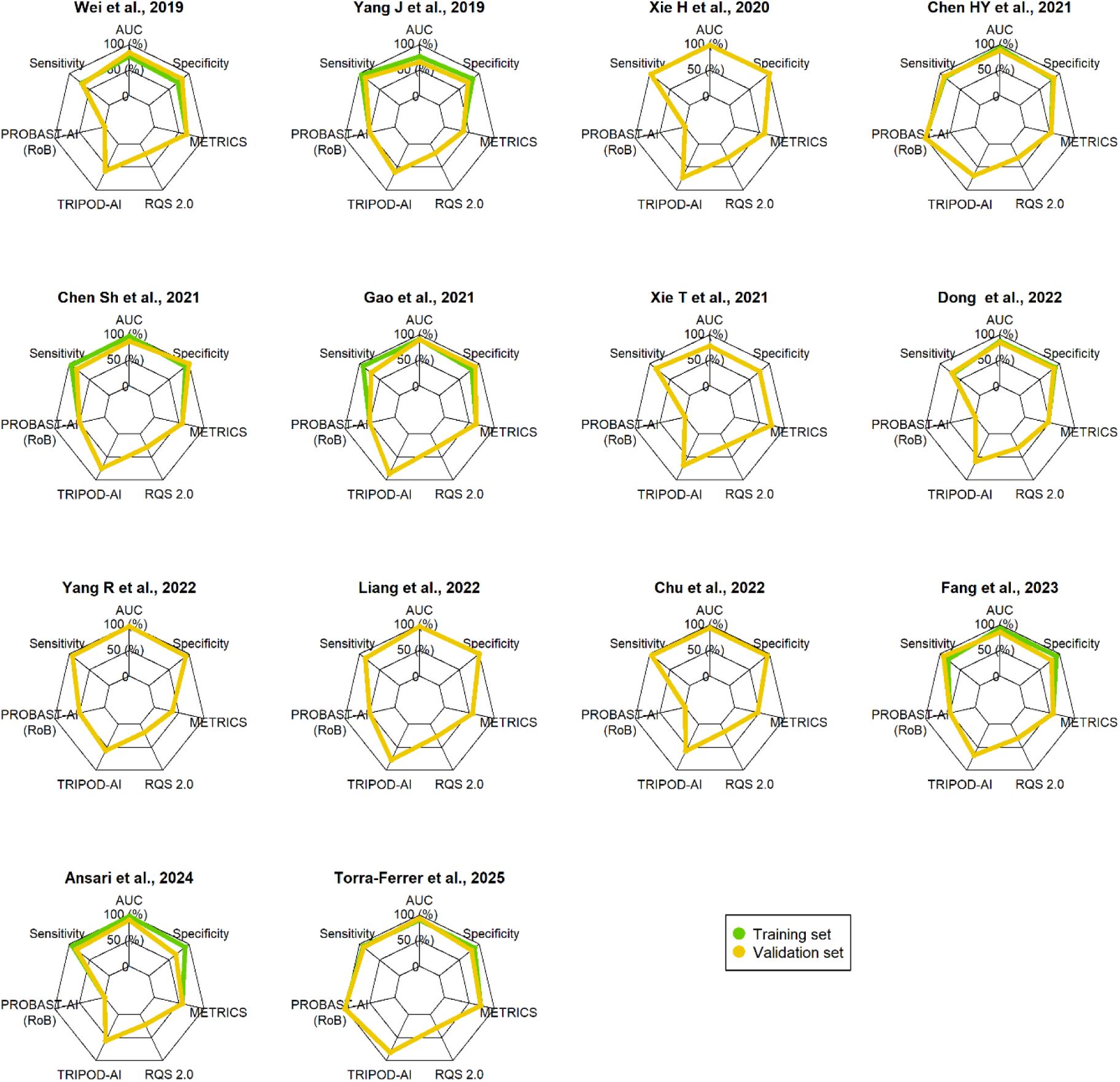
Study-level spider plots illustrate diagnostic performance and four-dimensional assessment metrics (Q1). Green lines represent training datasets and yellow lines represent validation datasets. Spider plots include only studies reporting complete performance metrics for both training and validation datasets.

#### 3.3.2 Assessments for malignancy or high-grade dysplasia prediction (Q2)

METRICS analysis similarly demonstrated substantial methodological heterogeneity for Q2. The study design domain was the only domain in which more than 75% of items were fulfilled (82%). Deficiencies were again observed in the imaging data and segmentation domains, with only marginally better adherence in image processing and feature extraction as well as feature processing, where scores ranged from 50% to 56%. The domains of preparation for modelling and metrics and comparison showed ratings comparable to Q1, whereas the domains of testing and open science exhibited identical ratings (33% and 0%) (Figure S7; Table S7).

RQS 2.0 scores increased cumulatively across early Radiomics Readiness Levels (RRLs), with the median score rising from 4 points at RRL1 to 13 points at RRL5. As observed in Q1, beyond this intermediate stage, no further increase in the median cumulative score was detected. In detail, only RRL1 (Foundational Exploration) showed fulfilment rates just above 50%. In Q2 RRL2 (Data Preparation) and RRL3 (Prototype Model Development) already fall below 50%, whereas RRL4 (Internal Validation) and RRL5 (Capability Testing) declined to approximately 25%. Similar to Q1, all higher readiness levels showed 0% overall fulfilment (Figure S10; Table S8). These findings are consistent with the results of RQS 1.0 (Table S9).

TRIPOD+AI assessment demonstrated a consistently incomplete and heterogeneous reporting profile across multiple domains (Figure 8A; Table S10). As in Q1, complete reporting was achieved for the title and abstract domains (100% each). Reporting completeness declined across the remaining manuscript sections, with fulfilment rates of 77.7% for the introduction and 70.3% for the methods domain. Domains related to transparency and participation showed particularly low adherence. Open science items were fulfilled in only 47.6% of cases, while patient and public involvement demonstrated the lowest reporting completeness at 28.6%, indicating a marked deficit in participatory and transparency-oriented reporting. Reporting results and discussion sections reached 64.7% and 57.1% fulfilment, respectively, reflecting moderate completeness but substantial variability across studies. Examination of study-level data confirmed pronounced heterogeneity within all domains below the 75% threshold, particularly for open science practices and patient and public involvement, where fulfilment ranged widely between studies (Table S10). Compared with Q1, both domains showed a further decline in overall adherence, indicating that increased clinical stakes in malignancy prediction were not accompanied by improved transparency or stakeholder involvement.

**Figure 8A.**
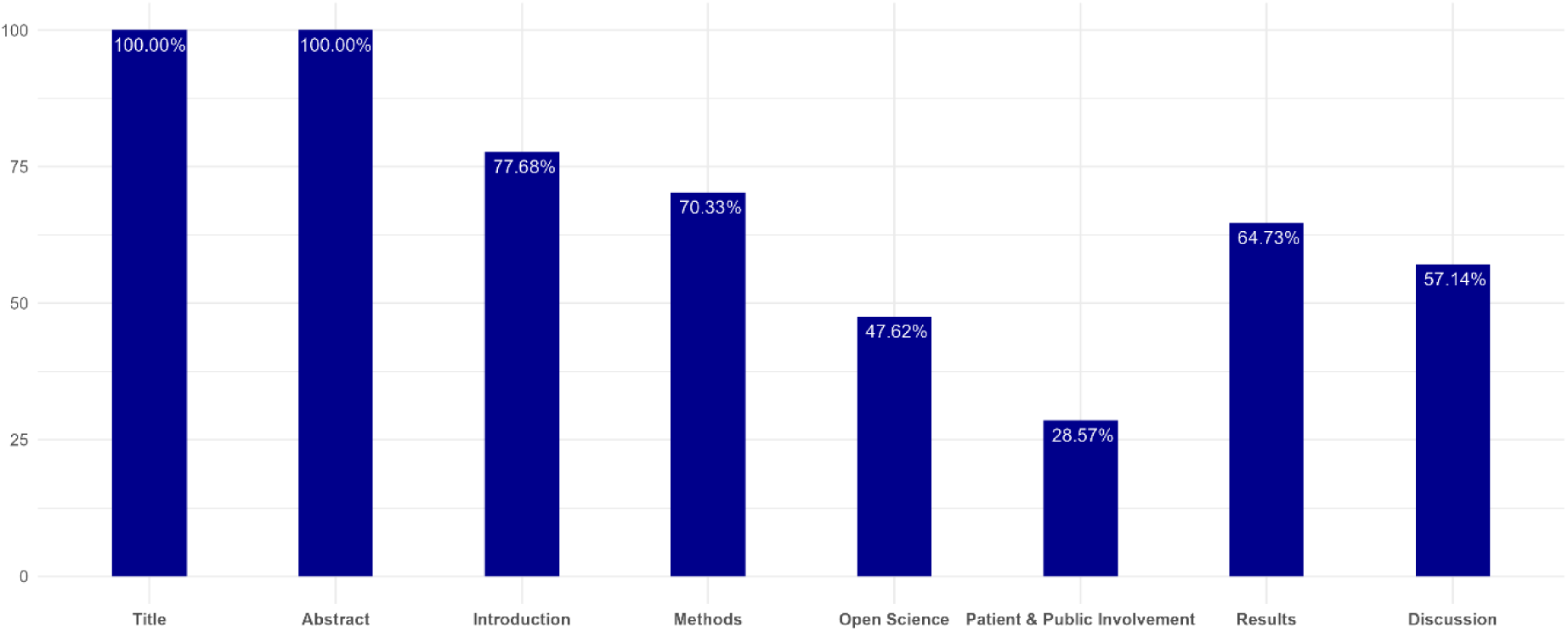
TRIPOD+AI for studies addressing Q2. Domain-level reporting completeness is shown as mean percentages across included studies. Detailed domain ratings are in the supplementary information.

PROBAST+AI revealed domain-specific risk-of-bias patterns in Q2, with generally low applicability concerns but substantial variability in risk-of-bias assessments across studies (Figure 8B; Table S11). At the domain level, participant selection again demonstrated predominantly low risk-of-bias ratings, indicating stable cohort definitions and clinically plausible target populations across Q2 studies. Consistent with the applicability assessment, concerns regarding applicability for participants and outcomes remained low throughout (Figure 8B). In contrast, the predictors domain showed a marked shift toward higher risk-of-bias ratings compared with Q1, driven by insufficient reporting on feature selection strategies, inadequate handling of high-dimensional radiomics feature spaces, and limited transparency regarding predictor preprocessing. Several studies were rated as high risk of bias in this domain contributing substantially to the increased proportion of overall high risk-of-bias classifications in Q2. The analysis domain exhibited the highest concentration of high risk-of-bias ratings in Q2, exceeding that observed in Q1. Frequent deficiencies included inadequate strategies to mitigate overfitting, limited use of robust internal validation procedures, and insufficient reporting of model calibration and uncertainty estimates. These shortcomings were consistently identified by both reviewers and represented the dominant driver of elevated overall risk-of-bias assessments in Q2 (Table S11). Compared with Q1, Q2 showed a higher proportion of overall high risk-of-bias classifications, while overall applicability concerns largely remained low.

**Figure 8B.**
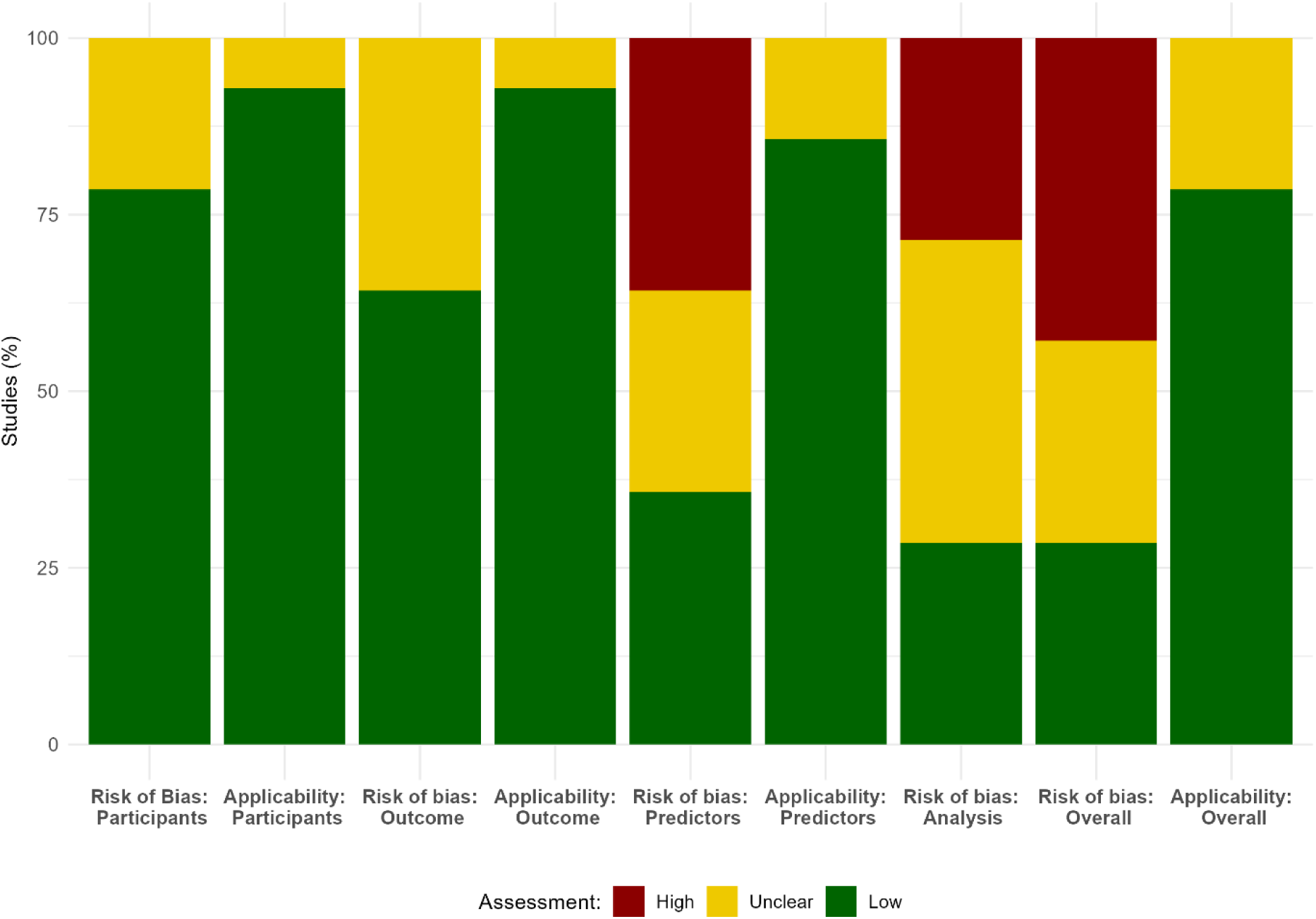
PROBAST+AI for studies addressing Q2. Domain-level distributions of risk of bias are shown as the percentage of studies classified as low, unclear, or high risk for each domain. Detailed domain ratings are provided in the Supplementary Information.

**Figure 9.**
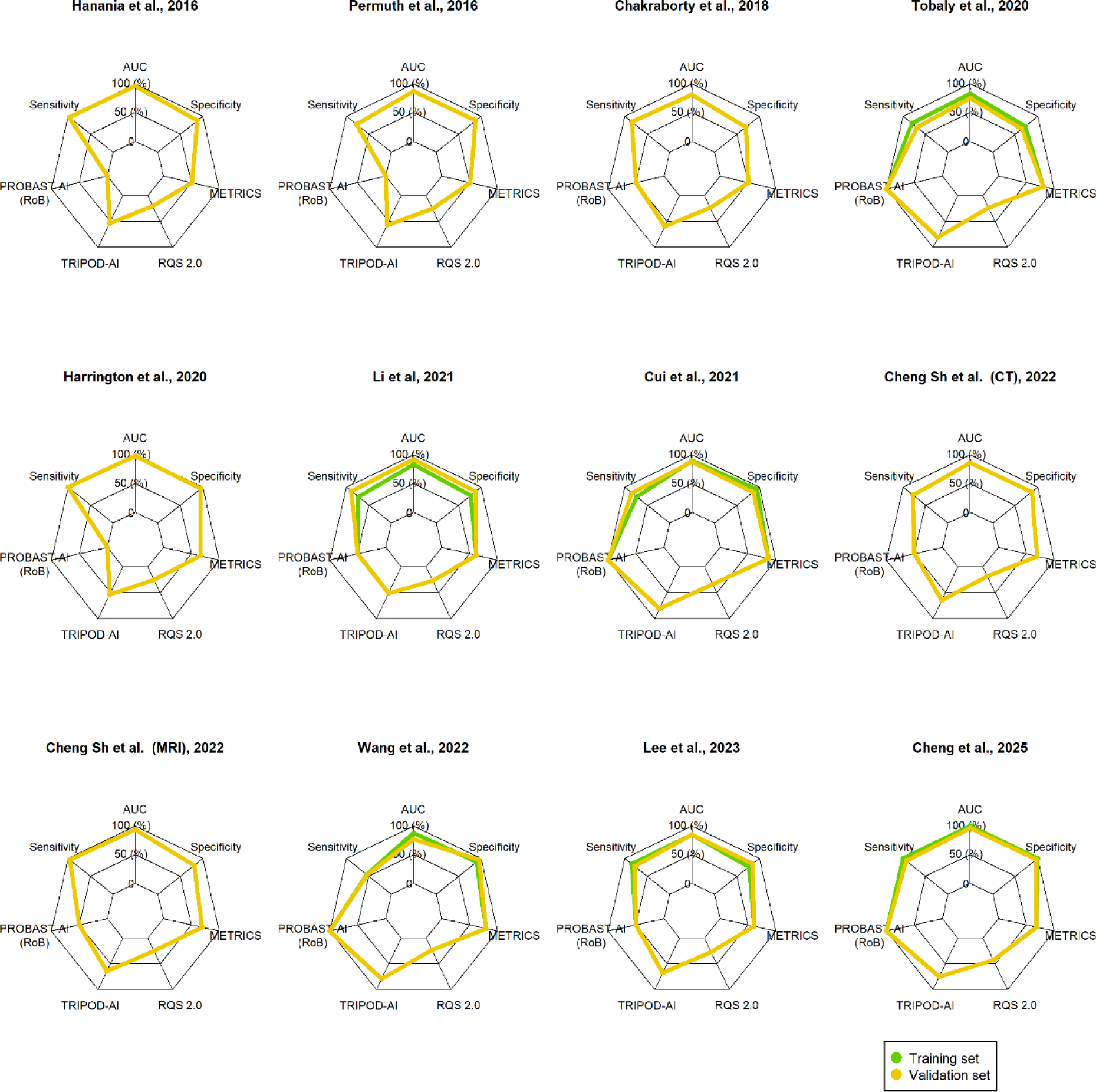
Study-level spider plots illustrate diagnostic performance and four-dimensional assessment metrics (Q2). Green lines represent training datasets and yellow lines represent validation datasets. Spiderplots include only studies reporting complete performance metrics for both training and validation datasets.

Multidimensional spider plot analysis demonstrated a consistent dissociation between diagnostic performance and methodological robustness across studies. As in Q1, high performance metrics observed in both training and validation datasets were frequently accompanied by lower scores for radiomics-specific assessments (RQS 2.0 and METRICS), reporting quality (TRIPOD+AI) and risk of bias (PROBAST+AI), despite the higher clinical stakes associated with malignancy prediction (Figure 8).

### 3.4 Inter-rater agreement

The inter-rater agreement between the two evaluators is summarized in Table 6. Agreement was quantified using weighted Cohen’s kappa with squared weights, which assigns greater penalties to larger rating discrepancies.

**Table 6.** Inter-rater agreement between reviewers using weighted Cohen’s kappa. Agreement was evaluated for METRICS, TRIPOD+AI, PROBAST+AI and RQS 2.0 separately for Q1 and Q2. Interpretation categories follow established kappa thresholds. For PROBAST+AI applicability in Q1, Cohen’s kappa was not estimable due to zero variance in one rater (all ratings classified as “low concern”); percent agreement is therefore reported instead (93.5%).

| Criterion | Q1 | Interpretation (Q1) | Q2 | Interpretation (Q2) |
| --- | --- | --- | --- | --- |
| METRICS | 0.48 | Moderate | 0.63 | Substantial |
| TRIPOD | 0.52 | Moderate | 0.49 | Moderate |
| PROBAST applicability | — | — | 0.41 | Moderate |
| PROBAST risk of bias | 0.84 | Almost perfect | 0.90 | Almost perfect |
| RQS2 | 0.84 | Almost perfect | 0.92 | Almost perfect |

The level of overall agreement varied across the assessment tools. METRICS and TRIPOD+AI demonstrated moderate to substantial agreement, indicating reasonable consistency alongside some inter-rater variability. For PROBAST+AI applicability, moderate agreement was observed for Q2. For Q1, Cohen’s kappa could not be meaningfully interpreted due to zero variance in one rater who rated all items as ‘low’; in this case, the percentage agreement was high (93.5%).

By contrast, PROBAST+AI risk of bias and RQS 2.0 showed almost perfect agreement for both Q1 and Q2, reflecting a high level of consistency between evaluators in these areas.

## 4. Discussion

### Principal findings and conceptual contribution

Over a decade, high diagnostic accuracy has not translated into clinical impact. Across two clinically distinct but related questions of cyst type differentiation (Q1) and malignancy or high-grade dysplasia prediction (Q2), our synthesis reveals a consistent pattern. AI-based radiomics models achieve high pooled discrimination with only modest declines from training to validation, yet substantial between-study heterogeneity persists, indicating limited transportability across clinical settings. Crucially, four-dimensional quality assessment identifies a field-wide performance–quality decoupling, where performance alone is insufficient for translation without attention to bias control and evaluation beyond isolated metrics^20,21,59^.

## Accuracy and heterogeneity

### Q1 – Cyst type differentiation: High discrimination with limited transportability

For Q1, pooled discrimination was high in both training and validation datasets, with only modest performance decrements. However, substantial between-study heterogeneity persisted, with prediction regions consistently exceeding confidence regions in SROC space, indicating limited transportability. Sensitivity analyses excluding non–cross-validated studies yielded overlapping credible intervals, supporting stability of the pooled signal but not its generalizability. Segmentation strategy emerged as a consistent translational bottleneck: manually segmented models showed numerically higher validation specificity than semi-automated approaches, underscoring the tension between apparent robustness under controlled conditions and scalability in clinical deployment, consistent with decade-long evidence from pancreas segmentation research^60^.

### Q2 – Malignancy or high-grade dysplasia prediction: Comparable performance with higher translational risk

For Q2, pooled discrimination remained high in both training and validation datasets, confirming the feasibility of AI-based radiomics malignancy risk stratification. Nonetheless, heterogeneity was more pronounced than in Q1, with wide prediction regions and marked differences in correlation structure between training and validation, suggesting threshold instability and calibration variability across studies. Unlike Q1, this heterogeneity has direct clinical consequences, as even small shifts in sensitivity or specificity may alter decisions between surveillance and surgery.

### The translational gap informed by multidimensional quality assessment

Building on these task-specific findings, our data identify a persistent proof-of-concept plateau separating reported performance from clinical application. Although cyst type differentiation (Q1) and malignancy or high-grade dysplasia prediction (Q2) represent distinct but sequential clinical decisions, assessment results converge on recurring clusters. Deficits systematically appeared at key transition points of the modelling pipeline: from data acquisition through preprocessing and segmentation, from model development to validation, and from performance reporting to transparency and governance. These breakpoints were consistently reflected across assessment frameworks. METRICS identified limitations in image acquisition, segmentation, and testing, RQS 2.0 indicated limited advancement beyond intermediate readiness levels. TRIPOD+AI revealed reduced reporting completeness beyond the methods section and PROBAST+AI demonstrated increased risk of bias related to predictors and model analysis. This underscores that a singular focus on technical barriers to implementation, such as segmentation or limited pipeline scalability alone, cannot bring about a systemic increase in clinical application. Notably, the transition from Q1 to the clinically more relevant Q2 setting was not accompanied by any systematic improvement in reporting completeness, radiomics readiness, or risk-of-bias control. On the contrary, assessment results for Q2 were consistently poorer across all dimensions, with a particularly pronounced increase in overall risk of bias, despite stable diagnostic performance.

This pattern suggests that escalating clinical stakes have not been translated into more rigorous study design or evaluation strategies, indicating that current AI-based radiomics research remains anchored in proof-of-concept paradigms even as clinical consequences become more substantial. Methodologically rigorous proof-of-concept studies with external validation demonstrate that bias reduction and robustness are, in principle, achievable^33,44,52,58^.

However, these advances have largely occurred within study designs that prioritize technical feasibility and performance optimization rather than downstream clinical integration. Multimodal models that are externally validated and combine radiomics with established clinical markers, including MRI-based approaches in an otherwise CT-dominated field, increase robustness and proximity to guidelines^52,56^. Nevertheless, increases in architectural and biological complexity, including deep ensemble models, end-to-end learning, direct benchmarking against classical radiomics, and multi-omics integration have predominantly extended the proof-of-concept paradigm without fundamentally altering study design, validation strategy, or clinical decision anchoring^40,46,54,58,61,62^. As a result, methodological innovation in this field has predominantly accumulated within existing evaluation paradigms, rather than prompting a corresponding evolution in how radiomics studies are framed, assessed, and interpreted in a clinical context.

### Limitations

First, subgroup analyses were exploratory and often underpowered. Quantitative synthesis was restricted to studies reporting AUC, sensitivity, and specificity, reflecting standard meta-analytical requirements. Studies lacking these metrics were included in qualitative and quality-based assessments but could not be pooled quantitatively due to incomplete reporting. Second, the scope was limited to AI-based radiomics approaches. Pure end-to-end convolutional neural network models were excluded due to heterogeneous reporting and fundamentally different modelling, improving internal coherence at the expense of coverage. Furthermore, the biological and clinical heterogeneity of pancreatic cystic neoplasms required abstraction to shared decision tasks, particularly for IPMNs, which does not fully reflect real-world complexity but enabled quantitative synthesis. The evidence base remains constrained by the predominance of CT-based models despite MRI being the clinical reference standard. Finally, while the four-dimensional assessment framework is resource-intensive, it is essential to avoid interpreting diagnostic performance in isolation: overlap between assessment domains reflects complementary perspectives across the model lifecycle rather than redundancy^27,28^. Inter-rater agreement was moderate for more interpretative assessment frameworks such as METRICS and TRIPOD+AI, reflecting inherent subjectivity in judgement-based reporting evaluations rather than a limitation of the analytical approach.

### Field-level claim: redefining progress in AI-based radiomics for pancreatic cysts

Existing evidence of AI-based radiomics in pancreatic cystic neoplasms has largely defined progress by improved diagnostic performance. High AUC values, sensitivities, and specificities are insufficient unless explicitly linked to predefined treatment goals, acceptable error trade-offs, and actionable distinctions between surveillance and surgical management. The findings of this study demonstrate that overcoming the translational gap requires a redefinition of progress. Rather than the further performance-driven optimization of model architectures, meaningful advancement is contingent on a shift towards decision-anchored, clinically applicable study designs that translate performance into operational clinical value. In the context of increasingly industry-driven AI development in healthcare, this underscores the continued importance of clinically grounded and academically engaged innovation to ensure effective translation^63^.

### Implications for future research and clinical translation

Consequently, models must be designed to support real clinical decision-making by radiologists and, ultimately, surgeons. Illustrative examples from clinically embedded AI research demonstrate how explicit decision anchoring and workflow integration can be operationalized at the level of study design, even if such approaches appear to be not yet broadly scalable^64^. Rather than model class, the critical determinant appears to be the alignment between model development and clinical decision-making processes. This requires stratifying cystic lesions according to clinical threat rather than simplified dichotomies that obscure biological and clinical heterogeneity, as misclassification at the level of outcome definition already constrains the validity of subsequent surgical risk assessment. External validation is necessary but does not guarantee clinical implementation. In the absence of actionable decision thresholds, explicit communication of uncertainties, and benchmarking against established clinical standards, including guidelines or expert interpretations, validation restricted to retrospective, resection-selected cohorts have limited translational value. In addition, challenges exist in particular in the multi-center harmonization of data sets and robust, reliable automatic segmentation^60,65^.

From a clinical perspective, future research should focus on MRI-based models, as MRI is the gold standard for monitoring pancreatic cysts and for perioperative assessment^5^. To ensure clinical interpretability and explainability, model architecture, especially in the case of hybrid and end-to-end deep learning approaches, should be evaluated against clinical reference standards, with radiomics serving as a transparent benchmark framework. Ultimately, model output must remain interpretable to patients. Future field evaluations should systematically analyze training and validation datasets separately, as they address different questions of model optimization versus robustness and transferability, while combining them obscures the performance degradation and decision instability that are critical for clinical interpretation. The development of AI-specific assessment frameworks highlights that performance gains must be interpreted in conjunction with methodological quality and bias control. Linking these assessments to meta-analytical performance data allows a more comprehensive evaluation of translational relevance.

## Conclusion

This study identifies a structural performance plateau in AI-based radiomics for pancreatic cystic neoplasms. By demonstrating significant heterogeneity and convergent limitations across multiple independent dimensions of methodological quality, it shows that further performance improvements alone are unlikely to lead to clinical benefits. These results provide empirical justification for a paradigm shift from performance-oriented optimization to study designs that explicitly target clinical translation.

## Data Availability

All data analyzed during this study are included in this published article and its Supplementary Information files. Further information are available from the corresponding author on reasonable request.

## Code Availability

The code used for data processing and meta-analytical analyses is available from the corresponding author upon reasonable request.

## Author Contributions

J.D.L. was responsible for conceptualization, methodology, data curation, visualization, original draft writing, review and editing, supervision, and project administration. T.E. was responsible for formal analysis, visualization, and review and editing. H.B. was responsible for review and editing. S.F.-F. was responsible for review and editing. C.N. was responsible for conceptualization, data curation, review and editing, and supervision. D.A.R. was responsible for conceptualization, review and editing, and supervision. All authors read and approved the final manuscript.

## Acknowledgements

This study was funded by the Research Commission of the University of Freiburg as the first phase of the RADIANCE project, which is being led by JD Lettner. The funder played no role in study design, data collection, analysis and interpretation of data, or the writing of this manuscript. T. Evrenoglou is supported German Research Foundation Deutsche Forschungsgemeinschaft; Project ID: 554095932. DA Ruess is supported by the German Research Foundation Deutsche Forschungsgemeinschaft; CRC1479 P17; Project ID: 441891347 and by the German Cancer Aid, Deutsche Krebshilfe; Project ID: 70113697.

## Competing Interests

All authors declare no financial or non-financial competing interests.

## Notes

### Competing Interest Statement

The authors have declared no competing interest.

