## Supplementary Information for "AI-based radiomics for pancreatic cysts: high diagnostic performance amid a persistent translational gap"

A four-dimensional framework review and Meta-analysis

### Supplementary Information

[1]Department of General and Visceral Surgery, Center for Surgery, Faculty of Medicine, Medical Center - University of Freiburg, Freiburg, Germany

[2]Department of Interventional and Diagnostic Radiology. University Hospital Freiburg, Freiburg, Germany

[3]Institute of Medical Biometry and Statistics, Faculty of Medicine and Medical Center—University of Freiburg, Germany

[†]Shared last authorship.

#### Abstract Checklist Prisma 2020

| Section and Topic | Item # | Checklist item | Reported (Yes/No) |
|---|---|---|---|
| **TITLE** | | | |
| Title | 1 | Identify the report as a systematic review. | Yes |
| **BACKGROUND** | | | |
| Objectives | 2 | Provide an explicit statement of the main objective(s) or question(s) the review addresses. | Yes |
| **METHODS** | | | |
| Eligibility criteria | 3 | Specify the inclusion and exclusion criteria for the review. | Yes |
| Information sources | 4 | Specify the information sources (e.g. databases, registers) used to identify studies and the date when each was last searched. | Yes |
| Risk of bias | 5 | Specify the methods used to assess risk of bias in the included studies. | Yes |
| Synthesis of results | 6 | Specify the methods used to present and synthesise results. | Yes |
| **RESULTS** | | | |
| Included studies | 7 | Give the total number of included studies and participants and summarise relevant characteristics of studies. | Yes |
| Synthesis of results | 8 | Present results for main outcomes, preferably indicating the number of included studies and participants for each. If meta-analysis was done, report the summary estimate and confidence/credible interval. If comparing groups, indicate the direction of the effect (i.e. which group is favoured). | Yes |
| **DISCUSSION** | | | |
| Limitations of evidence | 9 | Provide a brief summary of the limitations of the evidence included in the review (e.g. study risk of bias, inconsistency and imprecision). | Yes |
| Interpretation | 10 | Provide a general interpretation of the results and important implications. | Yes |
| **OTHER** | | | |
| Funding | 11 | Specify the primary source of funding for the review. | Yes |
| Registration | 12 | Provide the register name and registration number. | Yes |

From: Page MJ, McKenzie JE, Bossuyt PM, Boutron I, Hoffmann TC, Mulrow CD, et al. The PRISMA 2020 statement: an updated guideline for reporting systematic reviews. BMJ 2021;372:n71. doi: 10.1136/bmj.n71. This work is licensed under CC BY 4.0. To view a copy of this license, visit https://creativecommons.org/licenses/by/4.0/

#### Manuscript Checklist Prisma 2020

| Section and Topic | Item # | Checklist item | Location where item is reported |
|---|---|---|---|
| **TITLE** | | | |
| Title | 1 | Identify the report as a systematic review. | Title page |
| **ABSTRACT** | | | |
| Abstract | 2 | See the PRISMA 2020 for Abstracts checklist. | Supplementary Information |
| **INTRODUCTION** | | | |
| Rationale | 3 | Describe the rationale for the review in the context of existing knowledge. | 1. Introduction |
| Objectives | 4 | Provide an explicit statement of the objective(s) or question(s) the review addresses. | 1. Introduction |
| **METHODS** | | | |
| Eligibility | 5 | Specify the inclusion and exclusion criteria for the review and how studies were grouped for the syntheses. | 2.1 Study design and eligibility |
| Information | 6 | Specify all databases, registers, websites, organisations, reference lists and other sources searched or consulted to identify studies. | 2.2 Literature search and selection |
| Search | 7 | Present the full search strategies for all databases, registers and websites, including any filters and limits used. | Supplementary Information |
| Selection | 8 | Specify the methods used to decide whether a study met the inclusion criteria of the review, including how many reviewers screened | 2.2 Literature search and selection |
| Data collection | 9 | Specify the methods used to collect data from reports, including how many reviewers collected data from each report, whether they | 2.2 Literature search and selection |
| Data items | 10a | List and define all outcomes for which data were sought. Specify whether all results that were compatible with each outcome domain | 2.3. Data extraction |
| | 10b | List and define all other variables for which data were sought (e.g. participant and intervention characteristics, funding sources). | 2.1.-2.3 and Supplementary Info. |
| Study risk of | 11 | Specify the methods used to assess risk of bias in the included studies, including details of the tool(s) used, how many reviewers | 2.5 Quality assessment |
| Effect | 12 | Specify for each outcome the effect measure(s) (e.g. risk ratio, mean difference) used in the synthesis or presentation of results. | 2.41 DTA-MA |
| Synthesis methods | 13a | Describe the processes used to decide which studies were eligible for each synthesis (e.g. tabulating the study intervention | 2.4 Statistical analysis |
| | 13b | Describe any methods required to prepare the data for presentation or synthesis, such as handling of missing summary statistics, or | 2.4 Statistical analysis |
| | 13c | Describe any methods used to tabulate or visually display results of individual studies and syntheses. | 2.4 Statistical analysis |
| | 13d | Describe any methods used to synthesize results and provide a rationale for the choice(s). If meta-analysis was performed, describe | 2.4 Statistical analysis |
| | 13e | Describe any methods used to explore possible causes of heterogeneity among study results (e.g. subgroup analysis, meta- | 2.4 Statistical analysis |
| | 13f | Describe any sensitivity analyses conducted to assess robustness of the synthesized results. | 2.4 Statistical analysis |
| Reporting bias | 14 | Describe any methods used to assess risk of bias due to missing results in a synthesis (arising from reporting biases). | 2.4 Statistical analysis |
| Certainty | 15 | Describe any methods used to assess certainty (or confidence) in the body of evidence for an outcome. | 2.4 Statistical analysis |
| **RESULTS** | | | |
| Study | 16a | Describe the results of the search and selection process, from the number of records identified in the search to the number of studies | 3. Results, Figure 1 |

| Section and Topic | Item # | Checklist item | Location where item is reported |
|---|---|---|---|
| selection | 16b | Cite studies that might appear to meet the inclusion criteria, but which were excluded, and explain why they were excluded. | 3. Results, 3.1. |
| Study | 17 | Cite each included study and present its characteristics. | 3. Results, 3.1 |
| Risk of bias in | 18 | Present assessments of risk of bias for each included study. | 3.3. Results, 3.3.1 |
| Results of | 19 | For all outcomes, present, for each study: (a) summary statistics for each group (where appropriate) and (b) an effect estimate and its | 3. Results, 3.3.1 |
| Results of syntheses | 20a | For each synthesis, briefly summarise the characteristics and risk of bias among contributing studies. | 3. Results, 3.3.1 |
| | 20b | Present results of all statistical syntheses conducted. If meta-analysis was done, present for each the summary estimate and its | 3. Results, 3.2.1/ 3.2.2 |
| | 20c | Present results of all investigations of possible causes of heterogeneity among study results. | 3. Results, 3.2.1/ 3.2.2/ Supp |
| | 20d | Present results of all sensitivity analyses conducted to assess the robustness of the synthesized results. | 3. Results, 3.2.1/ 3.2.2/ Supp |
| Reporting | 21 | Present assessments of risk of bias due to missing results (arising from reporting biases) for each synthesis assessed. | 3. Results, 3.3.1 |
| Certainty of | 22 | Present assessments of certainty (or confidence) in the body of evidence for each outcome assessed. | 3. Results, 3.3.1 |
| **DISCUSSION** | | | |
| Discussion | 23a | Provide a general interpretation of the results in the context of other evidence. | 4. Discussion |
| | 23b | Discuss any limitations of the evidence included in the review. | 4. Discussion |
| | 23c | Discuss any limitations of the review processes used. | 4. Discussion |
| | 23d | Discuss implications of the results for practice, policy, and future research. | 4. Discussion |
| **OTHER INFORMATION** | | | |
| Registration and protocol | 24a | Provide registration information for the review, including register name and registration number, or state that the review was not | 2. Methods, 2.1. |
| | 24b | Indicate where the review protocol can be accessed, or state that a protocol was not prepared. | 2. Methods, 2.1. |
| | 24c | Describe and explain any amendments to information provided at registration or in the protocol. | 2. Methods, 2.1. |
| Support | 25 | Describe sources of financial or non-financial support for the review, and the role of the funders or sponsors in the review. | Acknowledgements |
| Competing | 26 | Declare any competing interests of review authors. | Competing interests |
| Availability of data, code and other materials | 27 | Report which of the following are publicly available and where they can be found: template data collection forms; data extracted from included studies; data used for all analyses; analytic code; any other materials used in the review. | Data- / Code Availability |

From: Page MJ, McKenzie JE, Bossuyt PM, Boutron I, Hoffmann TC, Mulrow CD, et al. The PRISMA 2020 statement: an updated guideline for reporting systematic reviews. BMJ 2021;372:n71. doi: 10.1136/bmj.n71. This work is licensed under CC BY 4.0. To view a copy of this license, visit https://creativecommons.org/licenses/by/4.0/

**Literature search strategy**

A comprehensive literature search was performed covering the period from 1 January 2015 to 31 July 2025. The complete database-specific search strings are reported to ensure transparency and reproducibility. For Google Scholar, a restricted title-based search was used to improve specificity. Search strategies were adapted to the syntax and indexing system of each database and applied as follows.

**PubMed**

“Radiomics”[MeSH Terms] OR “Artificial Intelligence”[MeSH Terms] OR “Machine Learning”[MeSH Terms] OR “Deep Learning”[MeSH Terms]) AND “Pancreas”[MeSH Terms] AND “Neoplasms”[MeSH Terms]

**Embase**

Radiomics OR Artificial Intelligence OR Machine Learning OR Deep Learning AND Pancreas AND Neoplasms

**IEEE Xplore**

Radiomics OR Artificial Intelligence AND Pancreas AND Neoplasms AND Cyst

**Google Scholar**

allintitle: radiomics pancreatic OR cysts OR artificial intelligence OR prediction OR serous OR neoplasm OR mucinous

#### Results

**Table S1. Imaging and Segmentation Characteristics of studies included in the meta-analysis.** Fifteen studies addressed cyst type differentiation (Q1) and fourteen studies addressed malignancy or high-grade dysplasia prediction in mucinous and IPMN lesions (Q2). Abbreviations: ROI, Region of Interest; VOI, Voxel of Interest

| Study | Modality | Phase used for analysis | Phases used for modeling (1= single,2= multiphase) | Minimum slice thickness (mm) | Maximum slice thickness (mm) | Preprocessing (yes= 1, no= 0) | Segmented Structure for modeling (1= Cyst only, 2= whole organ) | Segmentation unit (ROI/VOI) |
|---|---|---|---|---|---|---|---|---|
| **(Q1), n= 15** | | | | | | | | |
| Wei et al. (2019)[31] | CT | venous | 1 | 1 | 3 | 0 | 1 | ROI |
| Yang J et al. (2019)[32] | CT | venous | 1 | 2 | 5 | 0 | 1 | ROI |
| Xie H et al. (2020)[33] | CT | venous | 1 | 3 | 3 | 1 | 1 | VOI |
| Chen HY et al. (2021)[34] | CT | portal venous | 1 | 5 | 5 | 1 | 1 | ROI |
| Chen Sh et al. (2021)[35] | CT | noncontrast, arterial | 2 | 2 | 2 | 1 | 1 | VOI |
| Gao et al. (2021)[36] | CT | noncontrast, arterial | 2 | 1.5 | 1.5 | 0 | 1 | VOI |
| Xie T et al. (2021)[37] | CT | venous | 1 | 1 | 1 | 1 | 1 | VOI |
| Awe et al. (2022)[38] | CT | venous | 1 | 1.25 | 7 | 1 | 1 | VOI |
| Dong et al. (2022)[39] | CT | venous | 1 | 0.33 | 0.33 | 1 | 1 | VOI |
| Yang R et al. (2022)[40] | CT | arterial and venous | 2 | 3 | 3 | 0 | 1 | ROI |
| Liang et al. (2022)[21] | CT | arterial | 1 | 5 | 5 | 1 | 1 | ROI |
| Chu et al. (2022)[41] | CT | venous | 1 | 0.75 | 0.75 | 0 | 2 | VOI |
| Fang et al. (2023)[42] | MRI | T2WI | 1 | 5 | 5 | 0 | 1 | VOI |

| | | | | | | | | |
|---|---|---|---|---|---|---|---|---|
| Ansari et al. (2024)[43] | MRI | ADC | 1 | 5 | 5 | 1 | 1 | VOI |
| Torra-Ferrer et al. (2025)[44] | CT | Not reported | Not reported | Not reported | Not reported | 1 | 2 | ROI |
| **(Q2), n= 14** | | | | | | | | |
| Hanania et al. (2016)[45] | CT | arterial | 1 | Not reported | Not reported | 1 | 2 | ROI |
| Permuth et al. (2016)[46] | CT | arterial and venous | 1 | 3 | 3 | 0 | 1 | ROI |
| Chakraborty et al. (2018)[47] | CT | venous | 1 | 2.5 | 2.5 | 0 | 2 | ROI |
| Polk et al. (2020)[48] | CT | noncontrast, arterial, venous | 2 | 3 | 3 | 0 | 1 | ROI |
| Tobaly et al. (2020)[49] | CT | arterial or venous | 1 | 0.625 | 2.5 | 1 | 1 | VOI |
| Harrington et al. (2020)[50] | CT | venous | 1 | 2.5 | 2.5 | 0 | 1 | ROI |
| Li et al (2021)[51] | CT | venous | 1 | 1.5 | 1.5 | 1 | 1 | ROI |
| Cui et al. (2021)[52] | MRI | T2WI, noncontrast T1WI, T1WI arterial and venous | 2 | 2.5 | 6 | 1 | 1 | ROI |
| Cheng Sh et al. (2022)[53] | CT | arterial and venous | 2 | 5 | 5 | 1 | 1 | ROI |
| | MRI | T2WI | 1 | 5 | 5 | 1 | 1 | ROI |
| Wang et al. (2022)[54] | CT | arterial and venous | 2 | 1 | 1 | 1 | 1 | ROI |
| Flammia et al. (2023)[55] | MRI | T2WI, noncontrast T1WI, T1WI venous, ADC | 2 | Not reported | Not reported | 0 | 1 | ROI |
| Lee et al. (2024)[56] | CT | venous | 1 | <2.5 | 7 | 1 | 1 | VOI |
| Lou et al. (2024)[57] | CT | arterial and venous | 2 | 1 | 1 | 1 | 2 | VOI |
| Cheng Si et al. (2025)[58] | CT | venous | 1 | Not reported | Not reported | 0 | 2 | ROI |

**Table S2. Feature Characteristics of studies included in the meta-analysis.** Fifteen studies addressed cyst type differentiation (Q1) and fourteen studies addressed malignancy or high-grade dysplasia prediction in mucinous and IPMN lesions (Q2). Abbreviations: LBP, local binary pattern, ; DL, deep learning.

| Study | AI-Classifier (yes= 1, no= 0) | Radiomics feature composition (feature class; transformations; source) | Extracted radiomics features (n) | Significant radiomics features (n) | Non-radiomics features included in combined model |
|---|---|---|---|---|---|
| **(Q1), n= 15** | | | | | |
| Wei et al. (2019)[31] | 1 | Second; none; handcrafted | 409 | 22 | Clinical variables |
| Yang J et al. (2019)[32] | 1 | shape, first, second; none; handcrafted | Not reported | 7 | None |
| Xie H et al. (2020)[33] | 0 | shape, first, second; none; handcrafted | 1942 | 18 | Radiologic variables |
| Chen HY et al. (2021)[34] | 1 | shape, first, second; none; handcrafted | 271 | 13 | Radiologic variables |
| Chen Sh et al. (2021)[35] | 1 | shape, first, second; none; handcrafted | 710 | 15 | Clinical variables |
| Gao et al. (2021)[36] | 1 | shape, first, second; none; handcrafted | 1218 | 14 | Clinical variables |
| Xie T et al. (2021)[37] | 1 | shape, first, second; wavelet; handcrafted | 764 | 10 | None |
| Awe et al. (2022)[38] | 1 | shape, first, second; none; handcrafted | 95 | 95 | Radiologic variables |
| Dong et al. (2022)[39] | 1 | shape, first, second; wavelet; handcrafted | 944 | 5 | Radiologic variables |
| Yang R et al. (2022)[40] | 1 | shape, first, second; wavelet; handcrafted and DL | Not reported | Not reported | None |
| Liang et al. (2022)[21] | 1 | shape, first, second; wavelet, LBP; handcrafted and DL | 1067 | 73 | None |
| Chu et al. (2022)[41] | 1 | shape, first, second; none; handcrafted | 488 | 30 | Clinical variables |
| Fang et al. (2023)[42] | 1 | shape, first, second; none; handcrafted | 1409 | 4 | Clinical and radiologic variables |
| Ansari et al. (2024)[43] | 1 | shape, first, second; wavelet; handcrafted | 851 | 4 | None |
| Torra-Ferrer et al. (2025)[44] | 1 | shape, first, second; none; handcrafted | 214 | 38 | Radiologic variables |
| **(Q2), n= 14** | | | | | |
| Hanania et al. (2016)[45] | 0<br>0 | shape, first, second; none; handcrafted | 360 | 10 | None |
| Permuth et al. (2016)[46] | 1 | shape, first, second; none; handcrafted | 112 | 14 | Clinical variables |
| Chakraborty et al. (2018)[47] | 0 | second; none; handcrafted | 131 | 4 | Clinical variables |
| Polk et al. (2020)[48] | 1 | shape, first, second; none; handcrafted | 39 | 3 | Clinical and radiologic variables |
| Tobaly et al. (2020)[49] | 1 | shape, first, second; none; handcrafted | 107 | 85 | Clinical variables |

| | | | | | |
|---|---|---|---|---|---|
| Harrington et al. (2020)[50] | 1 | shape, first, second; wavelet; handcrafted | 850 | 13 | Clinical variables |
| Li et al (2021)[51] | 1 | shape, first, second; none; handcrafted | 436 | 8 | Clinical variables |
| Cui et al. (2021)[52] | 1 | shape, first, second; wavelet; handcrafted | 1312 | 9 | Clinical and radiologic variables |
| Cheng Sh et al. (2022)[53] - CT | 1 | shape, first, second; wavelet; handcrafted | 1320 | 8 | None |
| Cheng Sh et al. (2022)[53] - MRI | 1 | shape, first, second; none; handcrafted | 1037 | 10 | None |
| Wang et al. (2022)[54] | 1 | shape, first, second; none; handcrafted | 2436 | 18 | None |
| Flammia et al. (2023)[55] | 1 | shape, first, second; none; handcrafted | 107 | 67 | None |
| Lee et al. (2024)[56] | 1 | shape, first, second; none; handcrafted | 1218 | 11 | None |
| Lou et al. (2024)[57] | 1 | shape, first, second; wavelet; handcrafted | 1427 | 17 | Clinical variables |
| Cheng Si et al. (2025)[58] | 1 | shape, first, second; none; handcrafted | 321 | 19 | Clinical variables |

#### Diagnostic accuracy for cyst type differentiation (Q1)

#### Training datasets (Q1)

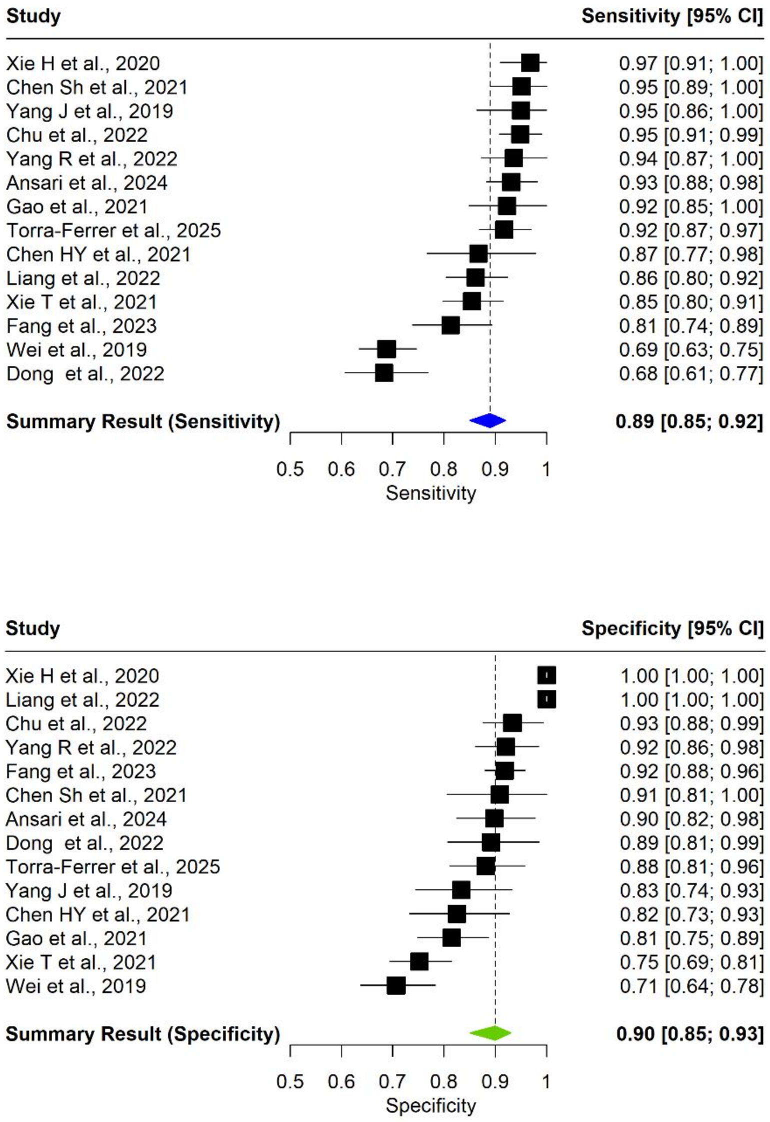


**Figure S1. Sensitivity and Specificity summary results of training dataset (Q1).** Studies not reporting sensitivity and specificity for the training dataset are not included in training-based analyses.

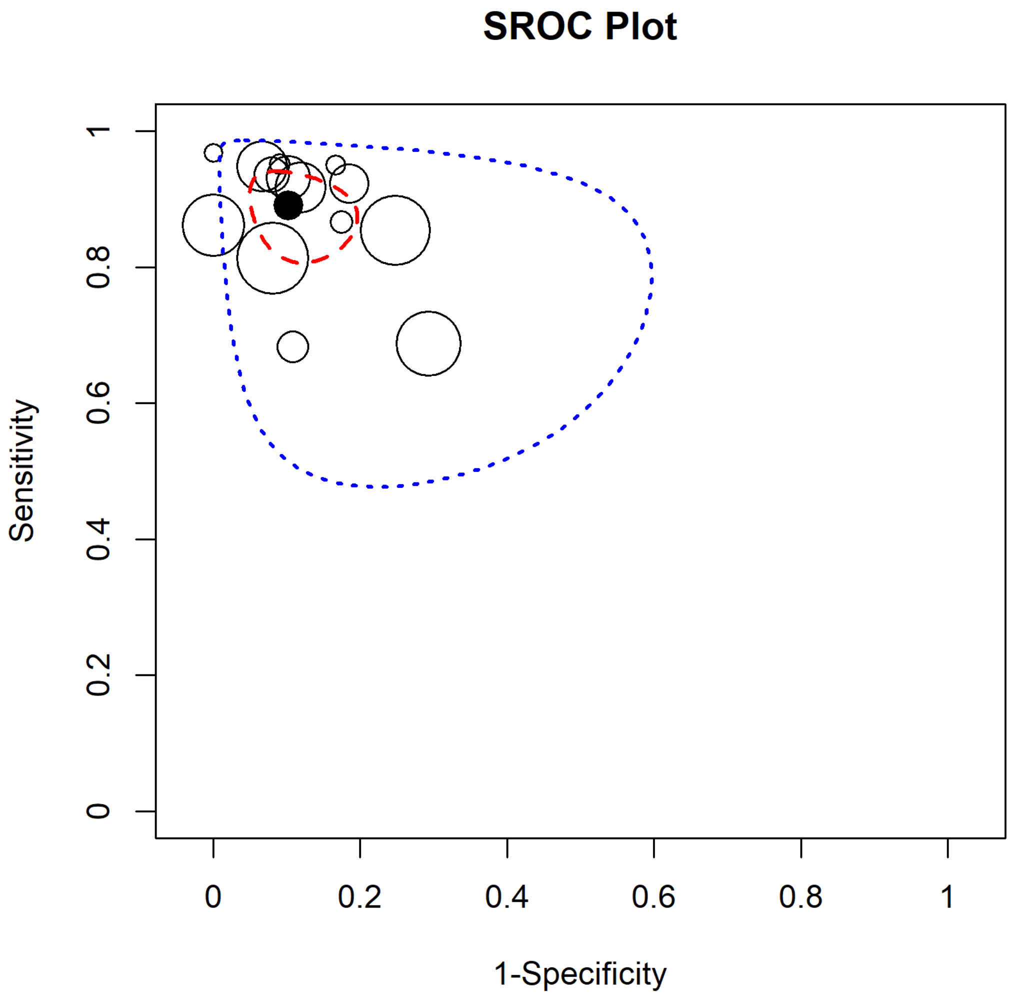


**Figure S2. SROC plot in terms of the training dataset for Q1.** The non-filled points represent the study-specific pairs of sensitivity and 1-specificity. The black filled point represents the pair of summary sensitivity and 1-specificity estimates. The region shown in terms of the red dashed line represents the confidence region while the region represented with a blue dotted line represent the prediction region.

| Characteristic | Subgroups (no. studies) | Sensitivity [95% CrI] | Specificity [95% CrI) |
|---|---|---|---|
| Type of center | Monocentric (12) | 0.89 [0.83, 0.94] | 0.90 [0.85, 0.95] |
| | Multicentric (2) | 0.90 [0.80, 0.96] | 0.86 [0.73, 0.93] |
| Type of validation | Internal (7) | 0.84 [0.76, 0.92] | 0.89 [0.80, 0.96] |
| | Internal Cross (3) | 0.92 [0.85, 0.97] | 0.91 [0.73, 0.99] |
| | External (3) | 0.91 [0.86, 0.95] | 0.87 [0.80, 0.93] |
| | None (1) | 0.94 [0.84, 0.98] | 0.92 [0.84, 0.97] |
| Deep Learning radiomics | Yes (2) | 0.89 [0.78, 0.95] | 0.97 [0.68, 0.99] |
| | No (12) | 0.89 [0.84, 0.93] | 0.87 [0.83, 0.91] |
| AI classifier | Yes (13) | 0.88 [0.84, 0.92] | 0.88 [0.84, 0.92] |
| | No (1) | 0.97 [0.87, 1.00] | 1.00 [1.00, 1.00] |
| Segmentation | Manual (12) | 0.90 [0.86, 0.97] | 0.91 [0.86, 0.95] |
| | Semi-Automated (2) | 0.77 [0.57, 0.91] | 0.81 [0.65, 0.93] |
| Image Modality | MRI (2) | 0.87 [0.69, 0.97] | 0.91 [0.84, 0.95] |
| | CT (12) | 0.89 [0.84, 0.93] | 0.89 [0.84, 0.94] |
| Number of phases | Uniphase (10) | 0.87 [0.81, 0.92] | 0.90 [0.84, 0.96] |
| | Multiphase (3) | 0.94 [0.88, 0.97] | 0.87 [0.78, 0.94] |
| | Not specified (1) | 0.92 [0.85, 0.96] | 0.88 [0.78, 0.95] |
| Segmented structure | Cysts Only (12) | 0.88 [0.83, 0.93] | 0.89 [0.84, 0.94] |
| | Cysts+Pancreas/Whole Organ (2) | 0.93 [0.87, 0.97] | 0.91 [0.81, 0.96] |
| Pre-Processing | Yes (8) | 0.88 [0.83, 0.93] | 0.92 [0.84, 0.97] |
| | No (6) | 0.88 [0.81, 0.94] | 0.87 [0.80, 0.92] |
| Non radiomics features | Clinical Variables (4) | 0.89 [0.79, 0.96] | 0.84 [0.75, 0.92] |
| | None (5) | 0.90 [0.85, 0.94] | 0.91 [0.76, 0.99] |
| | Radiological Variables (5) | 0.87 [0.74, 0.96] | 0.90 [0.83, 0.96] |
| | Clinical+Radiological Variables (1) | 0.81 [0.74, 0.89] | 0.92 [0.88, 0.96] |
| Region | USA (2) | 0.94 [0.88, 0.97] | 0.92 [0.83, 0.97] |
| | Asia (11) | 0.87 [0.81, 0.92] | 0.90 [0.83, 0.95] |
| | Europe (1) | 0.92 [0.85, 0.96] | 0.88 [0.78, 0.95] |
| Year | < 2022 (7) | 0.90 [0.83, 0.95] | 0.83 [0.77, 0.89] |
| | >= 2022 (7) | 0.88 [0.82, 0.93] | 0.93 [0.89, 0.96] |

**Table S3. Subgroup analysis results in terms of training dataset for Q1.** Numbers in bold indicate statistically significant results.

#### Validation datasets (Q1)

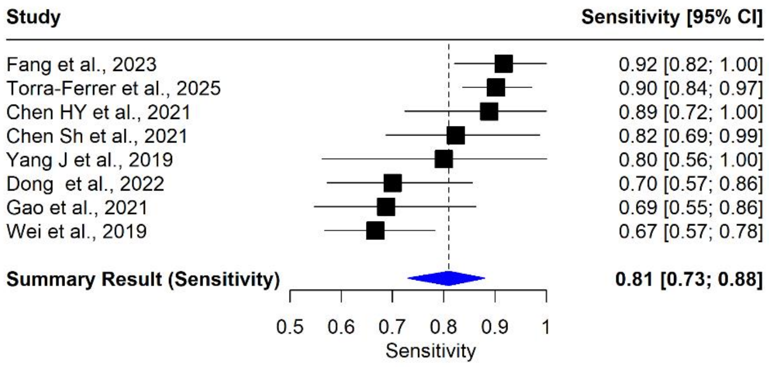


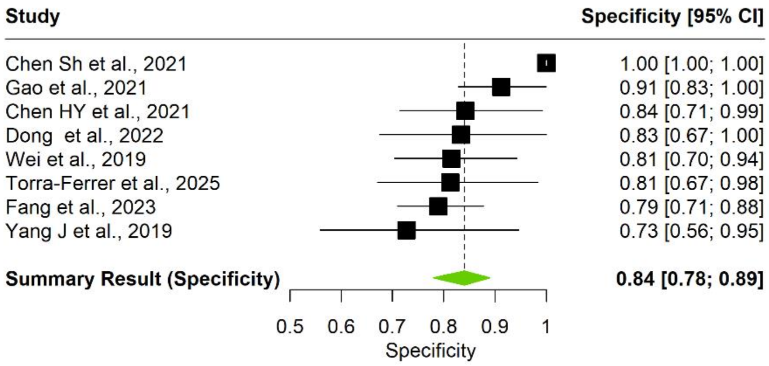


**Figure S3. Sensitivity analysis results for Sensitivity and Specificity of validation dataset (Q1).** Non-cross validated studies were excluded from the analysis. Studies not reporting sensitivity and specificity for the training dataset are not included in training-based analyses.

| Characteristic | Subgroups (no. studies) | Sensitivity [95% CrI] | Specificity [95% CrI) |
|---|---|---|---|
| Type of center | Monocentric (13) | 0.85 [0.80, 0.89] | 0.88 [0.81, 0.94] |
| | Multicentric (2) | 0.90 [0.77, 0.97] | 0.83 [0.65, 0.94] |
| Type of validation | Internal (7) | 0.79 [0.71, 0.86] | 0.91 [0.80, 0.98] |
| | Internal Cross (4) | 0.90 [0.81, 0.96] | 0.87 [0.70, 0.97] |
| | External (3) | 0.87 [0.78, 0.93] | 0.76 [0.62, 0.88] |
| | None (1) | 0.94 [0.87, 1.00] | 0.92 [0.86, 0.98] |
| Deep Learning radiomics | Yes (2) | 0.89 [0.78, 0.95] | 0.97 [0.68, 0.99] |
| | No (13) | 0.85 [0.80, 0.90] | 0.83 [0.77, 0.89] |
| AI classifier | Yes (13) | 0.85 [0.81, 0.89] | 0.86 [0.79, 0.92] |
| | No (1) | 0.97 [0.91, 1.00] | 1.00 [1.00, 1.00] |
| Segmentation | Manual (12) | 0.87 [0.82, 0.91] | **0.90 [0.84, 0.95]** |
| | Semi-Automated (3) | 0.80 [0.69, 0.87] | **0.74 [0.62, 0.84]** |
| Image Modality | MRI (2) | 0.85 [0.71, 0.96] | 0.73 [0.56, 0.86] |
| | CT (13) | 0.86 [0.80, 0.90] | 0.90 [0.83, 0.95] |
| Number of phases | Uniphase (10) | 0.85 [0.80, 0.90] | 0.86 [0.77, 0.94] |
| | Multiphase (3) | 0.84 [0.66, 0.95] | 0.93[0.86, 0.97] |
| | Not specified (1) | 0.90 [0.84, 0.97] | 0.81 [0.67, 0.98] |
| Segmented structure | Cysts Only (13) | 0.84 [0.79, 0.88] | 0.88 [0.80, 0.94] |
| | Cysts+Pancreas/Whole Organ (2) | 0.93 [0.84, 0.97] | 0.89 [0.72, 0.97] |
| Pre-Processing | Yes (9) | 0.85 [0.81, 0.89] | 0.90 [0.77, 0.98] |
| | No (6) | 0.86 [0.75, 0.95] | 0.87 [0.81, 0.92] |
| Non radiomics features | Clinical Variables (4) | 0.82 [0.63, 0.93] | 0.91 [0.84, 0.96] |
| | None (5) | 0.86 [0.82, 0.91] | 0.87 [0.62, 0.99] |
| | Radiological Variables (5) | 0.86 [0.77, 0.91] | 0.85 [0.71, 0.95] |
| | Clinical+Radiological Variables (1) | 0.92 [0.82, 1.00] | 0.79 [0.71, 0.88] |
| Region | USA (3) | 0.86 [0.75, 0.94] | 0.77 [0.55, 0.92] |
| | Asia (11) | 0.85 [0.79, 0.89] | 0.90 [0.83, 0.96] |
| | Europe (1) | 0.90 [0.84, 0.97] | 0.81 [0.67, 0.98] |
| Year | < 2022 (7) | 0.83 [0.74, 0.90] | 0.87 [0.78, 0.95] |
| | >= 2022 (8) | 0.87 [0.82, 0.91] | 0.87 [0.75, 0.95] |

**Table S4. Subgroup analysis results in terms of validation dataset (Q1).** Numbers in bold indicate statistically significant results.

#### Training Dataset (Q2)

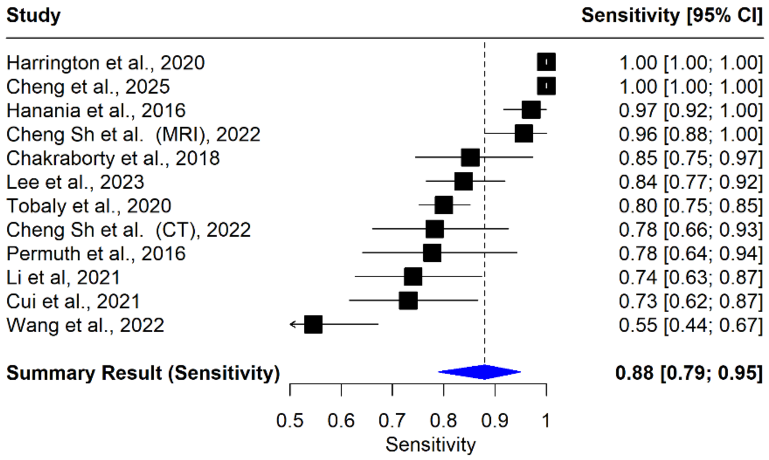


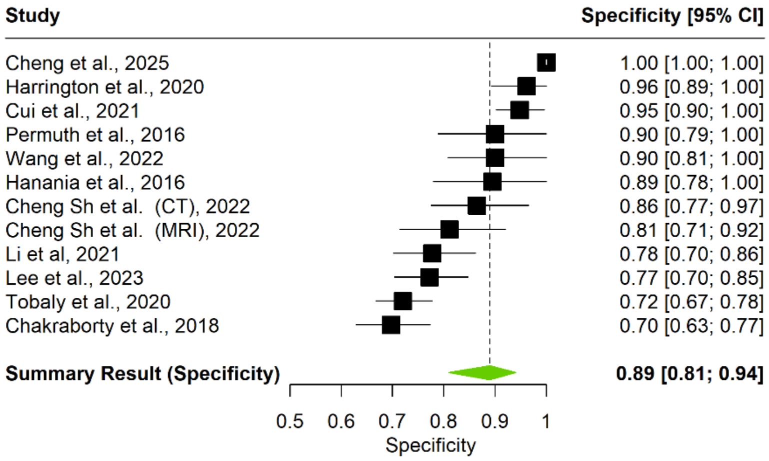


**Figure S4. Sensitivity and Specificity summary results for training dataset (Q2).** Studies not reporting sensitivity and specificity for the training dataset are not included in training-based analyses.

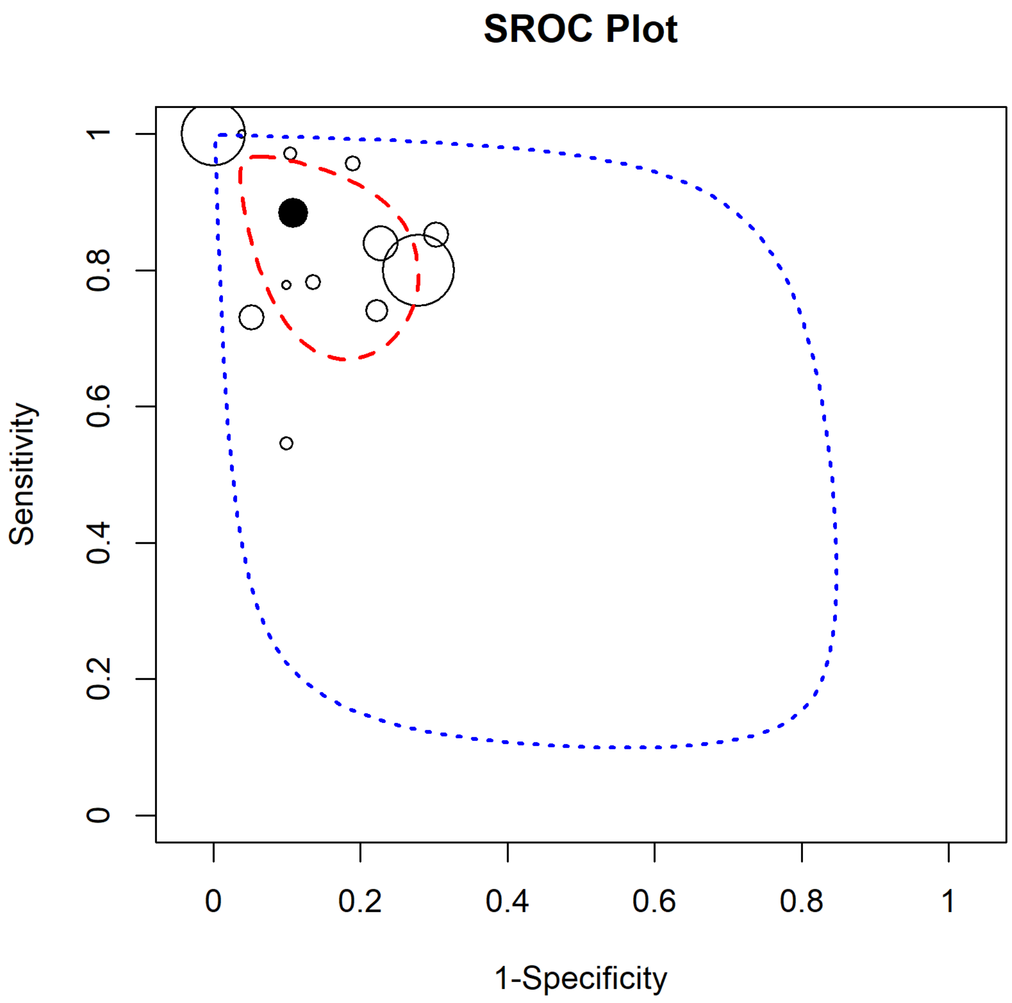


**Figure S5. SROC plot in terms of the training dataset (Q2).** The non-filled points represent the study-specific pairs of sensitivity and 1-specificity. The black filled point represents the pair of summary sensitivity and 1-specificity estimates. The region shown in terms of the red dashed line represent the confidence region while the region represented with a blue dotted line represent the prediction region

| Characteristic | Subgroups (no. studies) | Sensitivity [95% CrI] | Specificity [95% CrI) |
|---|---|---|---|
| Type of center | Monocentric (8) | 0.87 [0.80, 0.92] | 0.82 [0.76, 0.88] |
| | Multicentric (4) | 0.86 [0.47, 1.00] | 0.94 [0.72, 1.00] |
| Type of validation | Internal (2) | 0.80 [0.64, 0.91] | 0.77 [0.66, 0.86] |
| | Internal Cross (5) | 0.88 [0.80, 0.95] | 0.82 [0.74, 0.90] |
| | External (4) | 0.86 [0.47, 1.00] | 0.94 [0.72, 1.00] |
| | None (1) | 1.00 [1.00, 1.00] | 0.96 [0.89, 1.00] |
| AI classifier | Yes (10) | 0.87 [0.76, 0.95] | 0.88 [0.79, 0.95] |
| | No (2) | 0.88 [0.52, 0.99] | 0.90 [0.76, 0.97] |
| Segmentation | Manual (9) | 0.90 [0.78, 0.98] | 0.91 [0.83, 0.97] |
| | Semi-Automated (3) | 0.81 [0.73, 0.87] | 0.77 [0.67, 0.88] |
| Image Modality | MRI (2) | 0.84 [0.56, 0.98] | 0.89 [0.72, 0.97] |
| | CT (10) | 0.88 [0.78, 0.96] | 0.88 [0.79, 0.95] |
| Number of phases | Uniphase (9) | 0.92 [0.84, 0.98] | 0.88 [0.78, 0.95] |
| | Multiphase (3) | 0.69 [0.53, 0.82] | 0.91 [0.84, 0.96] |
| Segmented structure | Cysts Only (9) | 0.79 [0.73, 0.85] | 0.85 [0.79, 0.91] |
| | Cysts+Pancreas/Whole Organ (3) | 0.96 [0.81, 1.00] | 0.92 [0.59, 1.00] |
| Pre-Processing | Yes (8) | 0.81 [0.73, 0.89] | 0.83 [0.77, 0.89] |
| | No (4) | 0.95 [0.80, 1.00] | 0.94 [0.73, 1.00] |
| Non radiomics features | None (5) | 0.85 [0.69, 0.95] | 0.83 [0.77, 0.89] |
| | Clinical Variables (6) | 0.91 [0.78, 0.99] | 0.90 [0.74, 0.98] |
| | Clinical+Radiological Variables (1) | 0.73 [0.62, 0.87] | 0.95 [0.90, 1.00] |
| Region | USA (4) | 0.91 [0.80, 0.98] | 0.86 [0.73, 0.96] |
| | Asia (7) | 0.86 [0.69, 0.97] | 0.90 [0.79, 0.97] |
| | Europe (1) | 0.80 [0.75, 0.85] | 0.72 [0.67, 0.78] |
| Year | < 2022 (7) | 0.83 [0.76, 0.90] | 0.85 [0.76, 0.93] |
| | >= 2022 (5) | 0.90 [0.66, 1.00] | 0.91 [0.75, 0.99] |

**Table S5. Subgroup analysis results in terms of training dataset (Q2).** Numbers in bold indicate statistically significant results.

#### Validation Datasets (Q2)

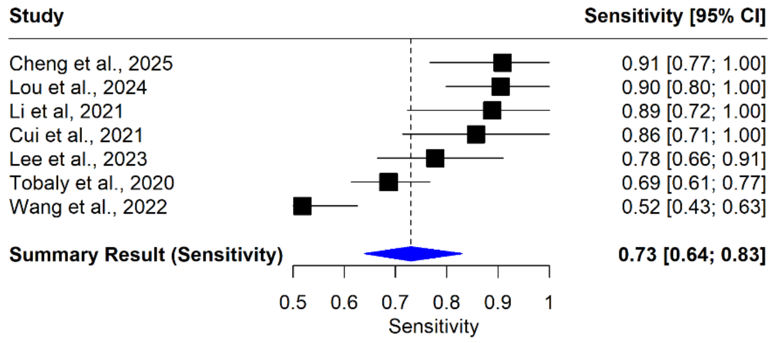


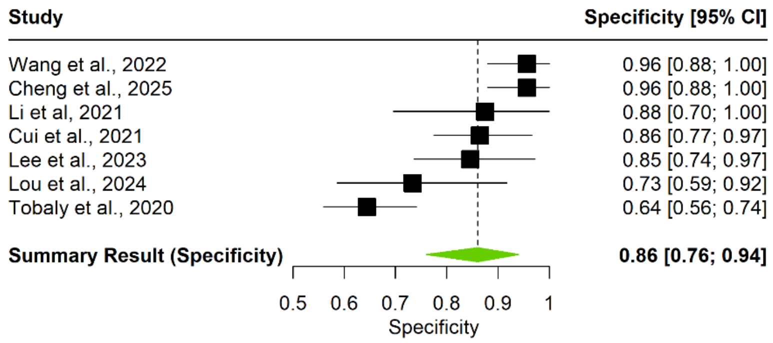


**Figure S6. Sensitivity analysis results for Sensitivity and Specificity of validation dataset (Q2).** Non-cross validated studies were excluded from the analysis. Studies not reporting sensitivity and specificity for the training dataset are not included in training-based analyses.

| Characteristic | Subgroups (no. studies) | Sensitivity [95% CrI] | Specificity [95% CrI) |
|---|---|---|---|
| Type of center | Monocentric (9) | 0.88 [0.82, 0.93] | 0.84 [0.78,0.90] |
| | Multicentric (4) | 0.72 [0.56, 0.88] | 0.87 [0.71, 0.97] |
| Type of validation | Internal (2) | 0.85 [0.72, 0.94] | 0.82 [0.68,0.92] |
| | Internal Cross (5) | 0.88 [0.80, 0.95] | 0.82 [0.74, 0.90] |
| | External (4) | 0.72 [0.56, 0.88] | 0.87 [0.71, 0.97] |
| | None (1) | 1.00 [1.00, 1.00] | 0.96 [0.89, 1.00] |
| AI classifier | Yes (11) | 0.83 [0.75, 0.90] | 0.84 [0.78, 0.90] |
| | No (2) | 0.88 [0.52, 0.99] | 0.90 [0.76, 0.97] |
| Segmentation | Manual (10) | 0.88 [0.80, 0.94] | 0.86 [0.80, 0.92] |
| | Semi-Automated (3) | 0.74 [0.63, 0.83] | 0.79 [0.62, 0.92] |
| Image Modality | MRI (2) | 0.92 [0.76, 0.98] | 0.84 [0.70, 0.93] |
| | CT (11) | 0.83 [0.75, 0.90] | 0.86 [0.79, 0.92] |
| Number of phases | Uniphase (9) | 0.88 [0.80, 0.94] | 0.84 [0.77, 0.91] |
| | Multiphase (4) | 0.77 [0.60, 0.91] | 0.87 [0.79, 0.94] |
| Segmented structure | Cysts Only (9) | 0.80 [0.71, 0.88] | 0.86 [0.80, 0.91] |
| | Cysts+Pancreas/Whole Organ (4) | 0.92 [0.84, 0.97] | 0.82 [0.68, 0.94] |
| Pre-Processing | Yes (9) | 0.83 [0.74, 0.92] | 0.83 [0.77, 0.89] |
| | No (4) | 0.87 [0.76, 0.95] | 0.88 [0.74, 0.97] |
| Non radiomics features | None (5) | 0.84 [0.66, 0.95] | 0.87 [0.81, 0.92] |
| | Clinical Variables (7) | 0.84 [0.76, 0.92] | 0.84 [0.73, 0.92] |
| | Clinical+Radiological Variables (1) | 0.86 [0.71, 1.00] | 0.86 [0.77, 0.97] |
| Region | USA (4) | **0.91 [0.80, 0.98]** | **0.86 [0.74, 0.96]** |
| | Asia (8) | 0.83 [0.74, 0.91] | 0.87 [0.82, 0.91] |
| | Europe (1) | **0.69 [0.61, 0.77]** | **0.64 [0.56, 0.74]** |
| Year | < 2022 (7) | 0.87 [0.78, 0.94] | 0.84 [0.75, 0.92] |
| | >= 2022 (6) | 0.82 [0.70, 0.92] | 0.87 [0.80, 0.92] |

**Table S6. Subgroup analysis results in terms of validation dataset and Q2.** Numbers in bold indicate statistically significant results.

#### Quality, risk of bias and radiomics specific assessments

##### Scoring approach for quality assessment frameworks

To enable structured comparison, aggregation, and visualization across studies and assessment frameworks, all quality instruments were operationalized using predefined numeric scoring systems. Checklist items were translated into ordinal numeric scores reflecting the degree of fulfillment, rather than being reported as narrative item-level comments, to facilitate cross-study comparability and domain-level synthesis.

For TRIPOD-AI, items were scored as 1 (fulfilled), 0.5 (partially fulfilled), or 0 (not fulfilled). For PROBAST-AI, items were scored as yes (1), probably yes (0.75), probably no (0.25), no (0), or no information (0), consistent with the original response options of the tool. PROBAST-AI scores were summarized at the domain level and separately for model development and evaluation phases.

While TRIPOD-AI and PROBAST-AI allow narrative item-level justification, free-text comments were deliberately not reported to avoid subjective weighting and to maintain a consistent quantitative framework across all applied instruments. This numeric operationalization does not replace qualitative judgment but provides a transparent and reproducible abstraction of checklist-based assessments aligned with the quantitative synthesis and visualization strategy of the present review.

Item-level assessments were performed independently by two reviewers according to predefined scoring rules. Given their volume, item-level data are not reported in full but are available from the authors upon reasonable request. Domain-level results and final framework-level judgments for each reviewer are fully reported in the Supplementary Tables.

#### METRICS (Q1)

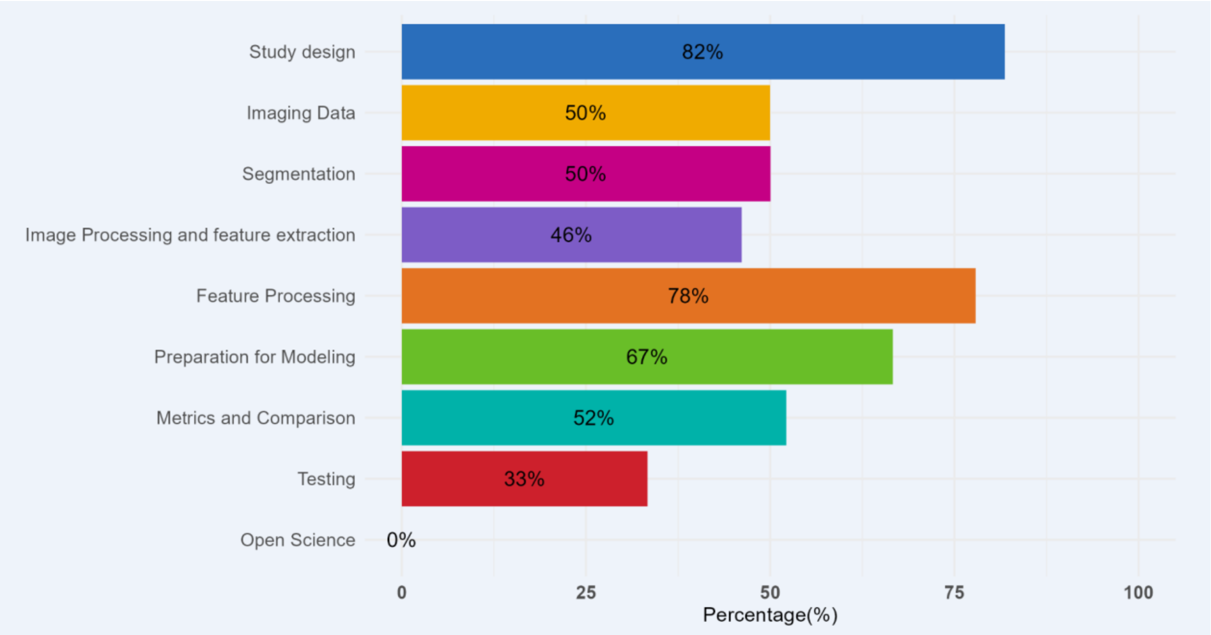


**Figure S7. Domain-level METRICS fulfillment rates for studies addressing cyst type differentiation (Q1).** Values represent the percentage of fulfilled items per domain across included studies.

#### METRICS (Q2)

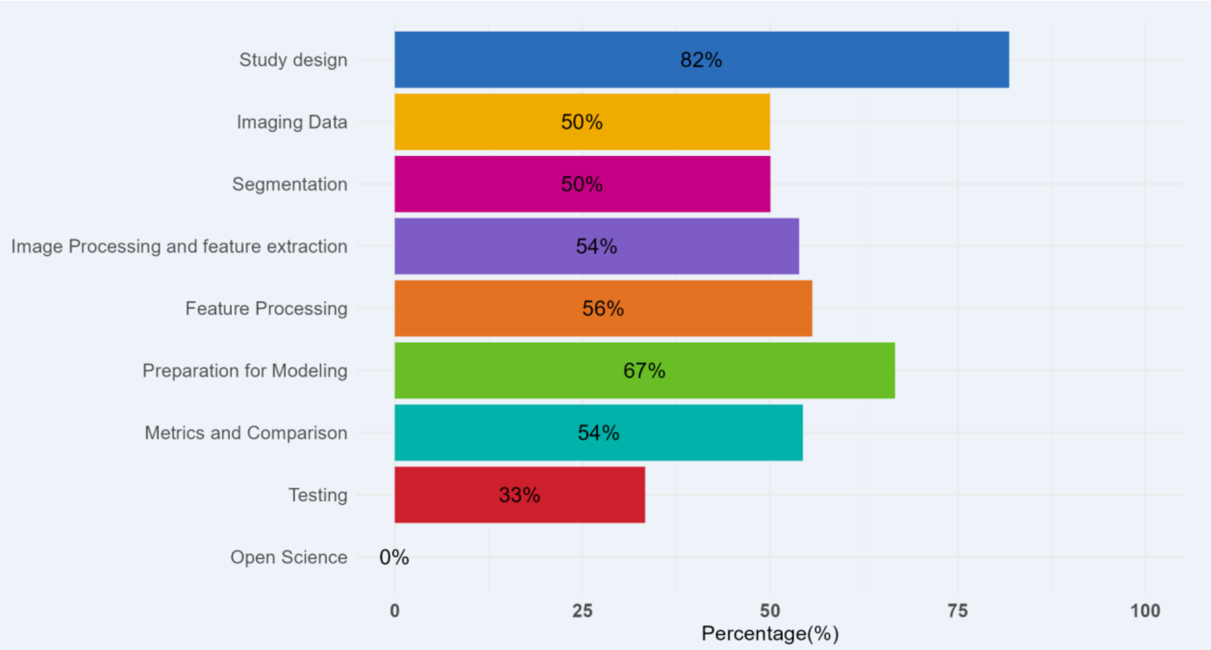


**Figure S8. Domain-level METRICS fulfillment rates for studies addressing malignancy prediction.** Values represent the percentage of fulfilled items per domain across included studies.

**Table S7. METRICS assessment results for studies addressing Q1 and Q2, Domain-levels reported separately for both independent reviewers.** The table summarizes METRICS domain scores for each included study, shown independently for Reviewer 1 and Reviewer 2 to ensure transparency of the assessment process.

Reviewer 1

| Question | Author | Study desigr | Imaging Data | Segmentatior | Image Processing and feature extractic | Feature Processing |
|---|---|---|---|---|---|---|
| Q1 | Wei et al. | 0.818 | 0.500 | 0.500 | 0.539 | 0.779 |
| Q1 | Yang J et al. | 0.818 | 0.500 | 0.000 | 0.000 | 0.779 |
| Q1 | Xie H et al. | 0.818 | 0.700 | 0.666 | 0.769 | 0.000 |
| Q1 | Chen HY et al. | 0.455 | 0.999 | 0.000 | 1.000 | 0.000 |
| Q1 | Chen Sh et al. | 0.818 | 0.700 | 0.500 | 1.000 | 0.000 |
| Q1 | Gao et al. | 0.818 | 0.500 | 0.666 | 0.539 | 0.000 |
| Q1 | Xie T et al. | 0.818 | 0.700 | 0.666 | 1.000 | 0.779 |
| Q1 | Awe et al. | 0.818 | 0.500 | 0.666 | 1.000 | 0.222 |
| Q1 | Dong et al. | 0.818 | 0.700 | 0.500 | 0.692 | 0.000 |
| Q1 | Yang R et al. | 0.818 | 0.500 | 0.500 | 0.461 | 0.000 |
| Q1 | Liang et al. | 0.818 | 0.200 | 0.000 | 1.000 | 0.667 |
| Q1 | Chu et al. | 0.818 | 0.500 | 0.500 | 0.000 | 0.000 |
| Q1 | Fang et al. | 0.818 | 0.700 | 0.000 | 0.231 | 0.779 |
| Q1 | Ansari et al. | 0.818 | 0.500 | 0.666 | 1.000 | 0.000 |
| Q1 | Torra-Ferrer et al. | 0.818 | 0.500 | 0.500 | 1.000 | 0.667 |
| Q2 | Hanania et al. | 0.818 | 0.200 | 0.500 | 0.692 | 0.334 |
| Q2 | Permuth et al. | 0.818 | 0.700 | 0.500 | 0.000 | 0.000 |
| Q2 | Chakraborty et al. | 0.818 | 0.500 | 0.166 | 0.769 | 0.334 |
| Q2 | Polk et al. | 0.818 | 0.500 | 0.166 | 0.000 | 0.222 |
| Q2 | Tobaly et al. | 0.818 | 0.999 | 0.666 | 0.692 | 0.556 |
| Q2 | Harrington et al. | 0.818 | 0.500 | 0.666 | 0.231 | 0.556 |
| Q2 | Li et al | 0.818 | 0.500 | 0.500 | 0.308 | 0.556 |
| Q2 | Cui et al. | 0.818 | 0.999 | 0.500 | 1.000 | 0.779 |
| Q2 | Cheng Sh et al. | 0.818 | 0.700 | 0.666 | 0.539 | 0.779 |
| Q2 | Wang et al. | 0.818 | 0.999 | 0.666 | 0.692 | 1.001 |
| Q2 | Flammia et al. | 0.818 | 0.700 | 0.500 | 0.231 | 0.000 |
| Q2 | Lee et al. | 0.818 | 0.700 | 0.666 | 1.000 | 1.001 |
| Q2 | Lou et al. | 0.818 | 0.500 | 0.500 | 0.692 | 0.000 |
| Q2 | Cheng Si et al. | 0.818 | 0.700 | 0.666 | 0.539 | 0.556 |

Reviewer 1

| Question | Author | Preparation for Modeling | Metrics and Comparison | Testing | Open Science | Total % | Category |
|---|---|---|---|---|---|---|---|
| Q1 | Wei et al. | 1.000 | 0.478 | 0.334 | 0.000 | 0.670 | Good |
| Q1 | Yang J et al. | 0.000 | 0.348 | 0.000 | 0.000 | 0.371 | Low |
| Q1 | Xie H et al. | 0.000 | 1.000 | 0.334 | 0.333 | 0.672 | Good |
| Q1 | Chen HY et al. | 0.666 | 0.478 | 1.000 | 0.000 | 0.726 | Good |
| Q1 | Chen Sh et al. | 0.666 | 1.000 | 0.334 | 0.000 | 0.753 | Good |
| Q1 | Gao et al. | 0.666 | 0.869 | 0.334 | 0.000 | 0.643 | Good |
| Q1 | Xie T et al. | 0.334 | 0.739 | 0.334 | 0.000 | 0.710 | Good |
| Q1 | Awe et al. | 0.000 | 0.522 | 0.334 | 0.333 | 0.578 | Moderate |
| Q1 | Dong et al. | 0.000 | 0.348 | 0.334 | 0.000 | 0.540 | Moderate |
| Q1 | Yang R et al. | 0.000 | 0.000 | 0.334 | 0.000 | 0.393 | Low |
| Q1 | Liang et al. | 0.666 | 0.869 | 0.334 | 0.000 | 0.618 | Good |
| Q1 | Chu et al. | 0.000 | 0.565 | 0.334 | 0.000 | 0.435 | Moderate |
| Q1 | Fang et al. | 0.666 | 1.000 | 0.334 | 0.000 | 0.628 | Good |
| Q1 | Ansari et al. | 0.000 | 0.739 | 0.334 | 0.000 | 0.626 | Good |
| Q1 | Torra-Ferrer et al. | 1.000 | 0.261 | 1.000 | 0.000 | 0.721 | Good |
| Q2 | Hanania et al. | 0.666 | 0.435 | 0.334 | 0.000 | 0.572 | Moderate |
| Q2 | Permuth et al. | 0.000 | 0.869 | 0.334 | 0.000 | 0.514 | Moderate |
| Q2 | Chakraborty et al. | 0.000 | 0.565 | 0.334 | 0.000 | 0.519 | Moderate |
| Q2 | Polk et al. | 0.000 | 0.739 | 0.334 | 0.000 | 0.425 | Moderate |
| Q2 | Tobaly et al. | 1.000 | 0.522 | 1.000 | 0.000 | 0.806 | Excellent |
| Q2 | Harrington et al. | 0.000 | 0.565 | 0.334 | 0.000 | 0.499 | Moderate |
| Q2 | Li et al | 0.666 | 0.478 | 0.334 | 0.000 | 0.549 | Moderate |
| Q2 | Cui et al. | 0.666 | 1.000 | 1.000 | 0.000 | 0.883 | Excellent |
| Q2 | Cheng Sh et al. | 0.334 | 0.869 | 0.334 | 0.333 | 0.662 | Good |
| Q2 | Wang et al. | 0.666 | 0.478 | 1.000 | 0.000 | 0.794 | Good |
| Q2 | Flammia et al. | 0.000 | 0.435 | 0.334 | 0.000 | 0.483 | Moderate |
| Q2 | Lee et al. | 0.666 | 0.869 | 0.334 | 0.000 | 0.764 | Good |
| Q2 | Lou et al. | 1.000 | 0.522 | 0.334 | 0.000 | 0.634 | Good |
| Q2 | Cheng Si et al. | 0.666 | 0.652 | 1.000 | 0.000 | 0.726 | Good |

Reviewer 2

| Question | Author | Study design | Imaging Data | Segmentation | Image Processing and feature extraction | Feature Processing |
|---|---|---|---|---|---|---|
| Q1 | Wei et al. | 1.000 | 0.500 | 0.500 | 0.308 | 0.779 |
| Q1 | Yang J et al. | 0.364 | 0.300 | 0.000 | 0.231 | 0.000 |
| Q1 | Xie H et al. | 0.818 | 0.700 | 0.666 | 0.308 | 0.000 |
| Q1 | Chen HY et al. | 0.818 | 0.799 | 0.000 | 0.461 | 0.779 |
| Q1 | Chen Sh et al. | 1.000 | 0.500 | 0.500 | 0.461 | 0.779 |
| Q1 | Gao et al. | 0.818 | 0.500 | 0.500 | 0.769 | 0.779 |
| Q1 | Xie T et al. | 0.818 | 0.700 | 0.500 | 0.769 | 0.779 |
| Q1 | Awe et al. | 1.000 | 0.300 | 0.666 | 1.000 | 0.779 |
| Q1 | Dong et al. | 0.818 | 0.200 | 0.500 | 0.000 | 0.779 |
| Q1 | Yang R et al. | 0.818 | 0.500 | 0.500 | 0.461 | 0.000 |
| Q1 | Liang et al. | 1.000 | 0.000 | 0.666 | 1.000 | 0.779 |
| Q1 | Chu et al. | 0.818 | 0.500 | 0.666 | 0.231 | 0.000 |
| Q1 | Fang et al. | 0.818 | 0.300 | 0.500 | 0.000 | 0.779 |
| Q1 | Ansari et al. | 0.818 | 0.500 | 0.500 | 0.461 | 0.556 |
| Q1 | Torra-Ferrer et | 0.818 | 0.500 | 0.500 | 0.769 | 0.779 |
| Q2 | Hanania et al. | 0.818 | 0.000 | 0.500 | 1.000 | 0.334 |
| Q2 | Permuth et al. | 0.818 | 0.300 | 0.500 | 0.308 | 0.000 |
| Q2 | Chakraborty et | 0.818 | 0.500 | 0.000 | 0.308 | 0.779 |
| Q2 | Polk et al. | 0.364 | 0.700 | 0.500 | 0.539 | 0.000 |
| Q2 | Tobaly et al. | 1.000 | 0.999 | 0.666 | 1.000 | 0.556 |
| Q2 | Harrington et a | 1.000 | 0.300 | 0.500 | 0.769 | 0.000 |
| Q2 | Li et al | 0.818 | 0.600 | 0.500 | 0.539 | 0.779 |
| Q2 | Cui et al. | 0.818 | 0.799 | 0.666 | 1.000 | 0.779 |
| Q2 | Cheng Sh et al. | 0.818 | 0.500 | 0.666 | 0.769 | 0.779 |
| Q2 | Wang et al. | 0.818 | 0.799 | 0.166 | 0.461 | 0.667 |
| Q2 | Flammia et al. | 0.818 | 0.500 | 0.500 | 0.308 | 0.556 |
| Q2 | Lee et al. | 0.818 | 0.300 | 0.500 | 0.539 | 0.779 |
| Q2 | Lou et al. | 0.818 | 0.300 | 0.500 | 0.769 | 0.222 |
| Q2 | Cheng Si et al. | 0.818 | 0.300 | 0.500 | 0.308 | 0.556 |

Reviewer 2

| Question | Author | Preparation for Modeling | Metrics and Comparison | Testing | Open Science | Total % | Category |
|---|---|---|---|---|---|---|---|
| Q1 | Wei et al. | 1.000 | 0.435 | 0.334 | 0.000 | 65.500 | Good |
| Q1 | Yang J et al. | 1.000 | 0.435 | 0.334 | 0.000 | 0.377 | Low |
| Q1 | Xie H et al. | 1.000 | 1.000 | 0.334 | 0.333 | 0.601 | Good |
| Q1 | Chen HY et al. | 0.000 | 0.565 | 0.000 | 0.000 | 0.530 | Moderate |
| Q1 | Chen Sh et al. | 0.000 | 0.565 | 0.000 | 0.333 | 0.566 | Moderate |
| Q1 | Gao et al. | 0.666 | 0.565 | 0.000 | 0.333 | 0.636 | Good |
| Q1 | Xie T et al. | 1.000 | 0.565 | 0.334 | 0.333 | 0.741 | Good |
| Q1 | Awe et al. | 1.000 | 0.435 | 0.334 | 0.667 | 0.728 | Good |
| Q1 | Dong et al. | 0.666 | 0.261 | 0.334 | 0.333 | 0.473 | Moderate |
| Q1 | Yang R et al. | 0.000 | 0.000 | 0.000 | 0.000 | 0.361 | Low |
| Q1 | Liang et al. | 0.000 | 0.392 | 0.334 | 0.000 | 0.566 | Moderate |
| Q1 | Chu et al. | 0.000 | 0.478 | 0.334 | 0.000 | 0.469 | Moderate |
| Q1 | Fang et al. | 1.000 | 0.652 | 0.334 | 0.000 | 0.570 | Moderate |
| Q1 | Ansari et al. | 0.666 | 0.261 | 0.334 | 0.333 | 0.566 | Moderate |
| Q1 | Torra-Ferrer et al. | 0.666 | 0.696 | 0.666 | 0.000 | 0.728 | Good |
| Q2 | Hanania et al. | 0.666 | 0.609 | 0.334 | 0.000 | 0.513 | Moderate |
| Q2 | Permuth et al. | 0.666 | 0.174 | 0.334 | 0.000 | 0.513 | Moderate |
| Q2 | Chakraborty et al. | 0.666 | 0.174 | 0.334 | 0.000 | 0.508 | Moderate |
| Q2 | Polk et al. | 0.666 | 0.174 | 0.334 | 0.333 | 0.429 | Moderate |
| Q2 | Tobaly et al. | 1.000 | 0.435 | 0.334 | 0.333 | 0.804 | Excellent |
| Q2 | Harrington et al. | 1.000 | 0.435 | 0.334 | 0.000 | 0.665 | Good |
| Q2 | Li et al | 0.666 | 0.478 | 0.334 | 0.000 | 0.617 | Good |
| Q2 | Cui et al. | 1.000 | 0.696 | 1.000 | 0.000 | 0.865 | Excellent |
| Q2 | Cheng Sh et al. | 0.666 | 0.652 | 0.334 | 0.000 | 0.693 | Good |
| Q2 | Wang et al. | 1.000 | 0.652 | 1.000 | 0.000 | 0.790 | Good |
| Q2 | Flammia et al. | 0.000 | 0.435 | 0.334 | 0.000 | 0.496 | Moderate |
| Q2 | Lee et al. | 0.666 | 0.783 | 0.334 | 0.000 | 0.615 | Good |
| Q2 | Lou et al. | 0.666 | 0.652 | 0.334 | 0.000 | 0.596 | Moderate |
| Q2 | Cheng Si et al. | 1.000 | 0.696 | 1.000 | 0.000 | 0.681 | Good |

#### RQS 2.0 (Q1)

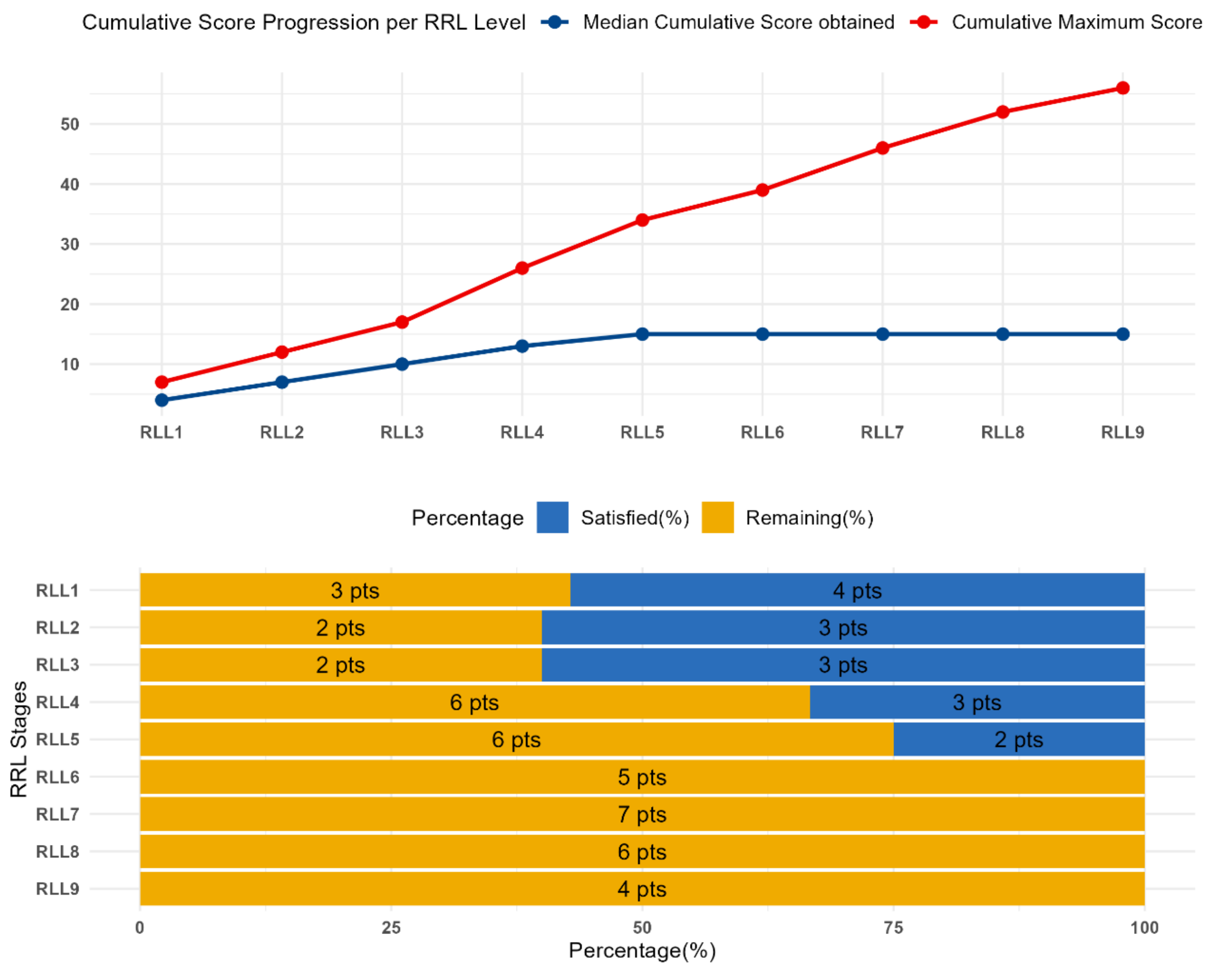


**Figure S9. Radiomics Quality Score (RQS) 2.0 cumulative score progression across radiomics readiness levels (RRL) for studies addressing cyst type differentiation (Q1).** The blue line represents the median cumulative score achieved per readiness level, while the red line indicates the cumulative maximum achievable score. The graphical representation follows the original RQS 2.0 framework as proposed by Lambin et al. (Lambin P, Woodruff HC, Mali SA, Zhong X, Kuang S, Lavrova Eet al.; Radiomics Quality Score 2.0: towards radiomics readiness levels and clinical translation for personalized medicine. Nat Rev Clin Oncol. 2025 Nov;22(11):831-846. doi: 10.1038/s41571-025-01067-1. Epub 2025 Sep 3. PMID: 40903523.)

#### RQS 2.0 (Q2)

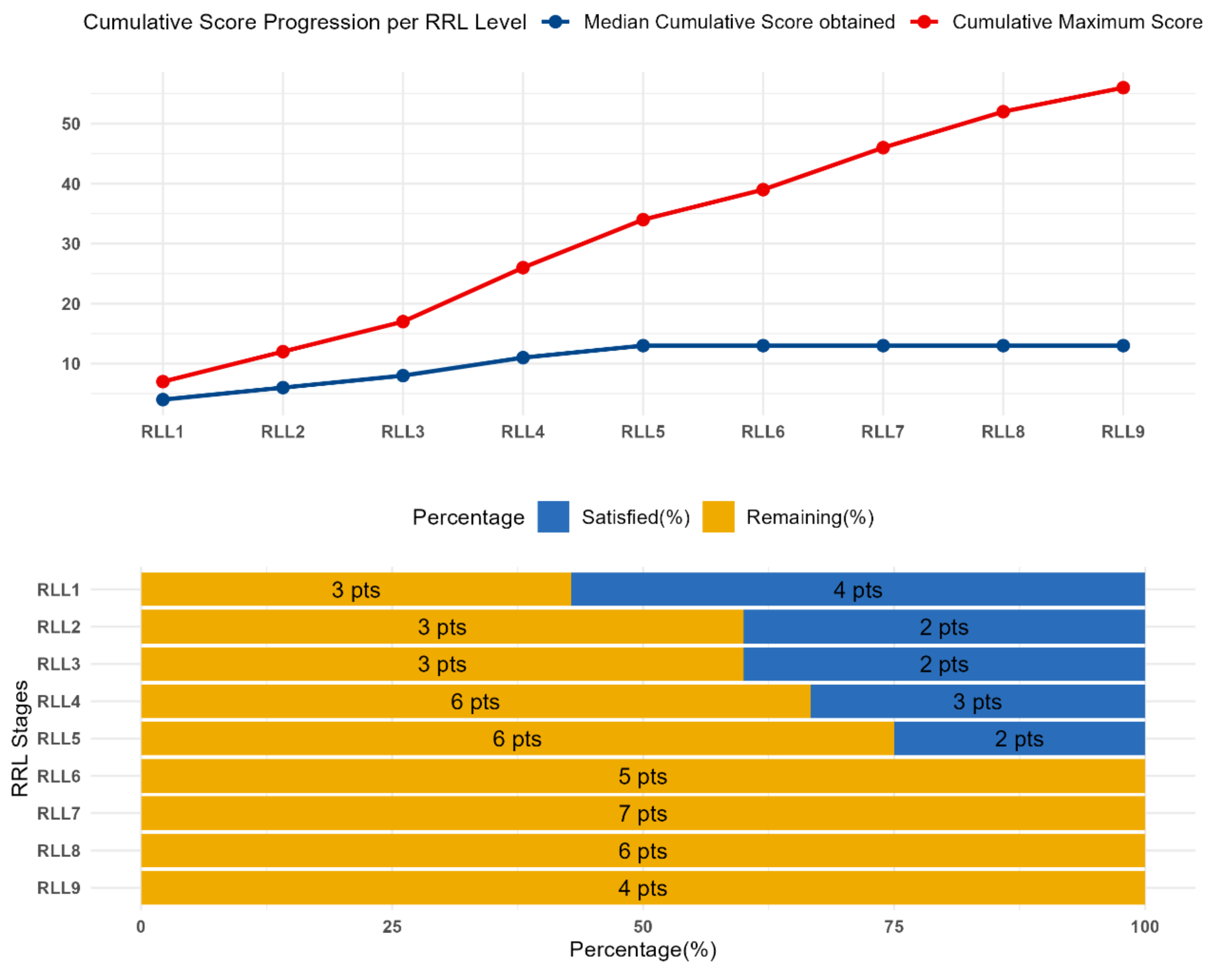


**Figure S10. Radiomics Quality Score (RQS) 2.0 cumulative score progression across radiomics readiness levels (RRL) for studies addressing malignancy prediction (Q2).** The blue line represents the median cumulative score achieved per readiness level, while the red line indicates the cumulative maximum achievable score. The graphical representation follows the original RQS 2.0 framework as proposed by Lambin et al. (Lambin P, Woodruff HC, Mali SA, Zhong X, Kuang S, Lavrova Eet al.; Radiomics Quality Score 2.0: towards radiomics readiness levels and clinical translation for personalized medicine. Nat Rev Clin Oncol. 2025 Nov;22(11):831-846. doi: 10.1038/s41571-025-01067-1. Epub 2025 Sep 3. PMID: 40903523.)

**Table S8. RQS 2.0 assessment**. Results for Q1 and Q2, reported separately for both independent reviewers.

Reviewer 1

| Question | Autor | RLL1 | RLL2 | RLL3 | RLL4 | RLL5 | RLL6 | RLL7 | RLL8 | RLL9 | Total | % |
|---|---|---|---|---|---|---|---|---|---|---|---|---|
| Q1 | Wei et al. | 3 | 2 | 1 | 3 | 2 | 0 | 0 | 1 | 0 | 12 | 0.21 |
| Q1 | Yang J et al. | 3 | 2 | 2 | 2 | 2 | 0 | 0 | 0 | 0 | 11 | 0.20 |
| Q1 | Xie H et al. | 4 | 3 | 4 | 4 | 2 | 0 | 0 | 0 | 0 | 17 | 0.30 |
| Q1 | Chen HY et al. | 4 | 3 | 3 | 4 | 4 | 1 | 0 | 0 | 1 | 20 | 0.36 |
| Q1 | Chen Sh et al. | 4 | 3 | 3 | 4 | 4 | 1 | 0 | 0 | 0 | 19 | 0.34 |
| Q1 | Gao et al. | 4 | 2 | 4 | 4 | 4 | 0 | 0 | 0 | 0 | 18 | 0.32 |
| Q1 | Xie T et al. | 4 | 3 | 2 | 3 | 2 | 0 | 0 | 0 | 0 | 14 | 0.25 |
| Q1 | Awe et al. | 3 | 2 | 2 | 3 | 0 | 1 | 0 | 0 | 0 | 11 | 0.20 |
| Q1 | Dong et al. | 4 | 4 | 4 | 3 | 2 | 0 | 0 | 0 | 0 | 17 | 0.30 |
| Q1 | Yang R et al. | 4 | 1 | 0 | 3 | 2 | 0 | 0 | 0 | 0 | 10 | 0.18 |
| Q1 | Liang et al. | 4 | 3 | 3 | 3 | 2 | 0 | 0 | 0 | 0 | 15 | 0.27 |
| Q1 | Chu et al. | 5 | 2 | 1 | 2 | 2 | 1 | 0 | 0 | 0 | 13 | 0.23 |
| Q1 | Fang et al. | 5 | 2 | 3 | 4 | 4 | 2 | 0 | 0 | 0 | 20 | 0.36 |
| Q1 | Ansari et al. | 4 | 3 | 2 | 2 | 1 | 0 | 0 | 0 | 0 | 12 | 0.21 |
| Q1 | Torra-Ferrer et al. | 4 | 3 | 4 | 2 | 3 | 0 | 0 | 0 | 0 | 16 | 0.29 |
| Q2 | Hanania et al. | 3 | 2 | 1 | 3 | 2 | 0 | 0 | 0 | 0 | 11 | 0.20 |
| Q2 | Permuth et al. | 4 | 1 | 2 | 4 | 2 | 1 | 0 | 0 | 0 | 14 | 0.25 |
| Q2 | Chakraborty et al. | 4 | 1 | 4 | 2 | 2 | 0 | 0 | 0 | 0 | 13 | 0.23 |
| Q2 | Polk et al. | 4 | 2 | 3 | 3 | 2 | 0 | 0 | 0 | 0 | 14 | 0.25 |
| Q2 | Tobaly et al. | 3 | 3 | 1 | 3 | 3 | 0 | 0 | 0 | 0 | 13 | 0.23 |
| Q2 | Harrington et al. | 4 | 1 | 3 | 2 | 2 | 0 | 0 | 0 | 0 | 12 | 0.21 |
| Q2 | Li et al | 3 | 1 | 3 | 3 | 1 | 2 | 0 | 0 | 0 | 13 | 0.23 |
| Q2 | Cui et al. | 4 | 2 | 3 | 4 | 6 | 0 | 0 | 0 | 0 | 19 | 0.34 |
| Q2 | Cheng Sh et al. | 4 | 4 | 1 | 2 | 2 | 0 | 0 | 0 | 0 | 13 | 0.23 |
| Q2 | Wang et al. | 4 | 3 | 1 | 4 | 3 | 0 | 0 | 0 | 0 | 15 | 0.27 |
| Q2 | Flammia et al. | 4 | 2 | 2 | 3 | 2 | 0 | 0 | 0 | 0 | 13 | 0.23 |
| Q2 | Lee et al. | 4 | 3 | 2 | 4 | 2 | 0 | 0 | 0 | 0 | 15 | 0.27 |
| Q2 | Lou et al. | 4 | 3 | 3 | 2 | 2 | 1 | 0 | 0 | 0 | 15 | 0.27 |
| Q2 | Cheng Si et al. | 3 | 2 | 4 | 4 | 6 | 2 | 3 | 0 | 0 | 24 | 0.43 |

Reviewer 2

| Question | Autor | RLL1 | RLL2 | RLL3 | RLL4 | RLL5 | RLL6 | RLL7 | RLL8 | RLL9 | Total | % |
|---|---|---|---|---|---|---|---|---|---|---|---|---|
| Q1 | Wei et al. | 3 | 1 | 2 | 3 | 2 | 0 | 0 | 0 | 0 | 11 | 0.20 |
| Q1 | Yang J et al. | 5 | 2 | 1 | 3 | 0 | 0 | 0 | 0 | 0 | 11 | 0.20 |
| Q1 | Xie H et al. | 5 | 3 | 3 | 3 | 2 | 2 | 0 | 0 | 0 | 18 | 0.32 |
| Q1 | Chen HY et al. | 5 | 3 | 4 | 3 | 5 | 1 | 0 | 0 | 0 | 21 | 0.38 |
| Q1 | Chen Sh et al. | 5 | 3 | 2 | 5 | 4 | 2 | 0 | 0 | 0 | 21 | 0.38 |
| Q1 | Gao et al. | 5 | 2 | 4 | 4 | 4 | 1 | 0 | 0 | 0 | 20 | 0.36 |
| Q1 | Xie T et al. | 5 | 2 | 4 | 2 | 0 | 1 | 0 | 0 | 0 | 14 | 0.25 |
| Q1 | Awe et al. | 4 | 1 | 2 | 3 | 0 | 1 | 0 | 0 | 0 | 11 | 0.20 |
| Q1 | Dong et al. | 5 | 3 | 4 | 3 | 1 | 0 | 0 | 0 | 0 | 16 | 0.29 |
| Q1 | Yang R et al. | 5 | 1 | 0 | 3 | 1 | 0 | 0 | 0 | 0 | 10 | 0.18 |
| Q1 | Liang et al. | 2 | 3 | 4 | 3 | 4 | 1 | 0 | 0 | 0 | 17 | 0.30 |
| Q1 | Chu et al. | 6 | 2 | 2 | 2 | 2 | 1 | 0 | 0 | 0 | 15 | 0.27 |
| Q1 | Fang et al. | 5 | 2 | 4 | 4 | 4 | 2 | 0 | 0 | 0 | 21 | 0.36 |
| Q1 | Ansari et al. | 4 | 3 | 2 | 4 | 0 | 0 | 0 | 0 | 0 | 13 | 0.23 |
| Q1 | Torra-Ferrer et al. | 3 | 3 | 4 | 4 | 4 | 0 | 0 | 0 | 0 | 18 | 0.32 |
| Q2 | Hanania et al. | 3 | 2 | 1 | 3 | 0 | 0 | 0 | 0 | 0 | 9 | 0.16 |
| Q2 | Permuth et al. | 5 | 1 | 3 | 2 | 0 | 1 | 0 | 0 | 0 | 12 | 0.21 |
| Q2 | Chakraborty et al. | 4 | 1 | 3 | 2 | 2 | 0 | 0 | 0 | 0 | 12 | 0.21 |
| Q2 | Polk et al. | 5 | 1 | 3 | 2 | 2 | 0 | 0 | 0 | 0 | 13 | 0.23 |
| Q2 | Tobaly et al. | 5 | 2 | 2 | 3 | 2 | 0 | 0 | 0 | 0 | 14 | 0.56 |
| Q2 | Harrington et al. | 4 | 0 | 3 | 2 | 2 | 0 | 0 | 0 | 0 | 11 | 0.20 |
| Q2 | Li et al | 3 | 1 | 3 | 3 | 1 | 1 | 0 | 0 | 0 | 12 | 0.21 |
| Q2 | Cui et al. | 5 | 2 | 4 | 4 | 6 | 0 | 0 | 0 | 0 | 21 | 0.38 |
| Q2 | Cheng Sh et al. | 5 | 4 | 2 | 1 | 2 | 0 | 0 | 0 | 0 | 14 | 0.25 |
| Q2 | Wang et al. | 5 | 3 | 1 | 4 | 3 | 0 | 0 | 0 | 0 | 16 | 0.29 |
| Q2 | Flammia et al. | 5 | 2 | 1 | 2 | 0 | 0 | 0 | 0 | 0 | 10 | 0.18 |
| Q2 | Lee et al. | 5 | 3 | 1 | 4 | 2 | 1 | 0 | 0 | 0 | 16 | 0.29 |
| Q2 | Lou et al. | 5 | 3 | 3 | 3 | 0 | 0 | 0 | 0 | 0 | 14 | 0.25 |
| Q2 | Cheng Si et al. | 3 | 3 | 3 | 5 | 6 | 2 | 3 | 0 | 0 | 25 | 0.45 |

**Table S9. RQS 1.0 assessment**. Results for Q1 and Q2, reported separately for both independent reviewers.

Reviewer 1

| Question | Author | Image protocol quality | Multiple Segmentations | Phantom Study | Imaging at multiple time | Feature reduction or adjustment for multiple testing | Multivariable analysis | Biological correlates |
|---|---|---|---|---|---|---|---|---|
| Q1 | Wei et al. | 0.5 | 0 | 0 | 0 | 1 | 1 | 1 |
| Q1 | Yang J et al. | 0.5 | 1 | 0 | 0 | 1 | 0 | 0 |
| Q1 | Xie H et al. | 0.5 | 0 | 0 | 0 | 1 | 1 | 1 |
| Q1 | Chen HY et al. | 0.5 | 0 | 0 | 0 | 1 | 1 | 1 |
| Q1 | Chen Sh et al. | 0.5 | 1 | 0 | 1 | 1 | 1 | 1 |
| Q1 | Gao et al. | 0.5 | 0 | 0 | 1 | 1 | 1 | 1 |
| Q1 | Xie T et al. | 0.5 | 0 | 0 | 0 | 1 | 1 | 1 |
| Q1 | Awe et al. | 0.5 | 0 | 0 | 0 | 1 | 0 | 0 |
| Q1 | Dong et al. | 0.5 | 1 | 0 | 0 | 1 | 1 | 0 |
| Q1 | Yang R et al. | 0.5 | 1 | 0 | 1 | 1 | 0 | 0 |
| Q1 | Liang et al. | 0 | 1 | 0 | 0 | 1 | 1 | 1 |
| Q1 | Chu et al. | 0.5 | 1 | 0 | 0 | 1 | 1 | 0 |
| Q1 | Fang et al. | 0.5 | 1 | 0 | 0 | 1 | 0 | 1 |
| Q2 | Ansari et al. | 0.5 | 0 | 0 | 0 | 1 | 0 | 0 |
| Q2 | Torra-Ferrer et al. | 0 | 1 | 0 | 0 | 1 | 1 | 1 |
| Q2 | Hanania et al. | 0 | 1 | 0 | 1 | 1 | 0 | 1 |
| Q2 | Permuth et al. | 0.5 | 1 | 0 | 0 | 1 | 1 | 1 |
| Q2 | Chakraborty et al. | 0.5 | 0 | 0 | 1 | 1 | 1 | 0 |
| Q2 | Polk et al. | 0.5 | 0 | 0 | 1 | 1 | 1 | 0 |
| Q2 | Tobaly et al. | 0.5 | 0 | 0 | 0 | 1 | 1 | 1 |
| Q2 | Harrington et al. | 0.5 | 0 | 0 | 0 | 1 | 1 | 1 |
| Q2 | Li et al | 0 | 1 | 0 | 0 | 1 | 1 | 1 |
| Q2 | Cui et al. | 0.5 | 0 | 0 | 1 | 1 | 1 | 1 |
| Q2 | Cheng Sh et al. | 0.5 | 0 | 0 | 0 | 1 | 0 | 1 |
| Q2 | Wang et al. | 0.5 | 0 | 0 | 1 | 1 | 0 | 0 |
| Q2 | Flammia et al. | 0.5 | 1 | 0 | 1 | 1 | 0 | 0 |
| Q2 | Lee et al. | 0.5 | 0 | 0 | 0 | 1 | 0 | 1 |
| Q2 | Lou et al. | 0.5 | 1 | 0 | 0 | 1 | 1 | 1 |
| Q2 | Cheng Si et al. | 0 | 0 | 0 | 0 | 1 | 1 | 1 |

Reviewer 1

| Question | Author | Cut-off analysis | Discrimination statistics | Calibration statistics | Prospective study | Validation | Comparison to gold standard | Potential clinical applications | Cost-effectiveness analysis |
|---|---|---|---|---|---|---|---|---|---|
| Q1 | Wei et al. | 0 | 1 | 0 | 0 | 0.4 | 1 | 0 | 0 |
| Q1 | Yang J et al. | 0 | 0.5 | 0 | 0 | 0.4 | 0 | 1 | 0 |
| Q1 | Xie H et al. | 0 | 0.5 | 1 | 0 | 0 | 0 | 1 | 0 |
| Q1 | Chen HY et al. | 0 | 1 | 0 | 0 | 0.6 | 1 | 1 | 0 |
| Q1 | Chen Sh et al. | 0 | 0.5 | 0.5 | 0 | 0.4 | 1 | 1 | 0 |
| Q1 | Gao et al. | 0 | 0.5 | 0 | 0 | 0.4 | 1 | 1 | 0 |
| Q1 | Xie T et al. | 1 | 1 | 0 | 0 | 0.4 | 1 | 0 | 0 |
| Q1 | Awe et al. | 0 | 1 | 0 | 0 | 0.4 | 0 | 0 | 0 |
| Q1 | Dong et al. | 0 | 0.5 | 0 | 0 | 0.4 | 0 | 1 | 0 |
| Q1 | Yang R et al. | 0 | 0 | 0 | 0 | 0.4 | 0 | 1 | 0 |
| Q1 | Liang et al. | 0 | 1 | 0.5 | 0 | 0.4 | 0 | 1 | 0 |
| Q1 | Chu et al. | 0 | 0.5 | 0 | 0 | 0.4 | 0 | 1 | 0 |
| Q1 | Fang et al. | 0 | 0.5 | 1 | 0 | 0.4 | 1 | 1 | 0 |
| Q2 | Ansari et al. | 0 | 0.5 | 0 | 0 | 0.4 | 0 | 1 | 0 |
| Q2 | Torra-Ferrer et al. | 0 | 0.5 | 0 | 0 | 0.8 | 0 | 1 | 0 |
| Q2 | Hanania et al. | 0 | 1 | 0 | 0 | 0.4 | 1 | 1 | 0 |
| Q2 | Permuth et al. | 0 | 1 | 0 | 0 | 0.4 | 1 | 0 | 0 |
| Q2 | Chakraborty et al. | 0 | 1 | 0 | 0 | 0.4 | 1 | 0 | 0 |
| Q2 | Polk et al. | 0 | 1 | 0 | 0 | 0.4 | 1 | 0 | 0 |
| Q2 | Tobaly et al. | 1 | 1 | 0 | 0 | 0.6 | 0 | 1 | 0 |
| Q2 | Harrington et al. | 0 | 0 | 0 | 0 | 0.4 | 1 | 0 | 0 |
| Q2 | Li et al | 0 | 1 | 0 | 0 | 0.4 | 0 | 0 | 0 |
| Q2 | Cui et al. | 1 | 1 | 0.5 | 0 | 0.8 | 1 | 1 | 0 |
| Q2 | Cheng Sh et al. | 0 | 1 | 0 | 0 | 0.4 | 1 | 0 | 0 |
| Q2 | Wang et al. | 0 | 1 | 0 | 0 | 0.8 | 1 | 0 | 0 |
| Q2 | Flammia et al. | 0 | 0.5 | 0 | 0 | 0.4 | 0 | 1 | 0 |
| Q2 | Lee et al. | 0 | 1 | 0 | 0 | 0.4 | 1 | 0 | 0 |
| Q2 | Lou et al. | 0 | 1 | 0 | 0 | 0.4 | 0 | 0 | 0 |
| Q2 | Cheng Si et al. | 1 | 1 | 0.5 | 0 | 1 | 0 | 1 | 1 |

Reviewer 1

| Question | Author | Open sciene and | Total | % |
|---|---|---|---|---|
| Q1 | Wei et al. | 0 | 12 | 0.333 |
| Q1 | Yang J et al. | 0 | 10 | 0.278 |
| Q1 | Xie H et al. | 0 | 11 | 0.306 |
| Q1 | Chen HY et al. | 0 | 15 | 0.417 |
| Q1 | Chen Sh et al. | 0 | 16 | 0.444 |
| Q1 | Gao et al. | 0 | 14 | 0.389 |
| Q1 | Xie T et al. | 0 | 13 | 0.361 |
| Q1 | Awe et al. | 0.25 | 9 | 0.250 |
| Q1 | Dong et al. | 0 | 11 | 0.306 |
| Q1 | Yang R et al. | 0 | 10 | 0.278 |
| Q1 | Liang et al. | 0 | 13 | 0.361 |
| Q1 | Chu et al. | 0 | 11 | 0.306 |
| Q1 | Fang et al. | 0 | 15 | 0.417 |
| Q2 | Ansari et al. | 0 | 9 | 0.250 |
| Q2 | Torra-Ferrer et al. | 0 | 13 | 0.361 |
| Q2 | Hanania et al. | 0 | 14 | 0.389 |
| Q2 | Permuth et al. | 0 | 13 | 0.361 |
| Q2 | Chakraborty et al. | 0 | 12 | 0.333 |
| Q2 | Polk et al. | 0 | 12 | 0.333 |
| Q2 | Tobaly et al. | 0 | 14 | 0.389 |
| Q2 | Harrington et al. | 0 | 10 | 0.278 |
| Q2 | Li et al | 0 | 10 | 0.278 |
| Q2 | Cui et al. | 0 | 19 | 0.528 |
| Q2 | Cheng Sh et al. | 0 | 11 | 0.306 |
| Q2 | Wang et al. | 0 | 13 | 0.361 |
| Q2 | Flammia et al. | 0 | 11 | 0.306 |
| Q2 | Lee et al. | 0 | 11 | 0.306 |
| Q2 | Lou et al. | 0 | 11 | 0.306 |
| Q2 | Cheng Si et al. | 0 | 17 | 0.472 |

Reviewer 2

| Question | Author | Image protocol | Multiple Segmentations | Phantom Study | Imaging at multiple time | Feature reduction or adjustment for multiple testing | Multivariable analysis | Biological correlates |
|---|---|---|---|---|---|---|---|---|
| Q1 | Wei et al. | 0.5 | 0 | 0 | 0 | 1 | 1 | 1 |
| Q1 | Yang J et al. | 0.5 | 1 | 0 | 0 | 1 | 0 | 0 |
| Q1 | Xie H et al. | 0.5 | 0 | 0 | 0 | 1 | 1 | 1 |
| Q1 | Chen HY et al. | 0.5 | 0 | 0 | 0 | 1 | 1 | 1 |
| Q1 | Chen Sh et al. | 0.5 | 1 | 0 | 1 | 1 | 1 | 1 |
| Q1 | Gao et al. | 0.5 | 0 | 0 | 1 | 1 | 1 | 1 |
| Q1 | Xie T et al. | 0.5 | 0 | 0 | 0 | 1 | 0 | 1 |
| Q1 | Awe et al. | 0.5 | 0 | 0 | 0 | 1 | 0 | 0 |
| Q1 | Dong et al. | 0.5 | 1 | 0 | 0 | 1 | 1 | 0 |
| Q1 | Yang R et al. | 0.5 | 1 | 0 | 1 | 1 | 0 | 0 |
| Q1 | Liang et al. | 0 | 1 | 0 | 0 | 1 | 1 | 1 |
| Q1 | Chu et al. | 0.5 | 1 | 0 | 0 | 1 | 1 | 0 |
| Q1 | Fang et al. | 0.5 | 1 | 0 | 0 | 1 | 0 | 1 |
| Q2 | Ansari et al. | 0.5 | 0 | 0 | 0 | 1 | 0 | 0 |
| Q2 | Torra-Ferrer et al. | 0 | 1 | 0 | 0 | 1 | 1 | 1 |
| Q2 | Hanania et al. | 0 | 1 | 0 | 1 | 1 | 0 | 1 |
| Q2 | Permuth et al. | 0.5 | 1 | 0 | 0 | 1 | 1 | 1 |
| Q2 | Chakraborty et al. | 0.5 | 0 | 0 | 0 | 1 | 1 | 1 |
| Q2 | Polk et al. | 0.5 | 1 | 0 | 1 | 1 | 1 | 0 |
| Q2 | Tobaly et al. | 0.5 | 0 | 0 | 0 | 1 | 1 | 1 |
| Q2 | Harrington et al. | 0.5 | 0 | 0 | 0 | 1 | 1 | 1 |
| Q2 | Li et al | 0.5 | 1 | 0 | 0 | 1 | 1 | 1 |
| Q2 | Cui et al. | 0.5 | 0 | 0 | 1 | 1 | 1 | 1 |
| Q2 | Cheng Sh et al. | 0.5 | 0 | 0 | 0 | 1 | 0 | 1 |
| Q2 | Wang et al. | 0.5 | 1 | 0 | 1 | 1 | 0 | 0 |
| Q2 | Flammia et al. | 0.5 | 1 | 0 | 1 | 1 | 0 | 0 |
| Q2 | Lee et al. | 0.5 | 0 | 0 | 0 | 1 | 0 | 1 |
| Q2 | Lou et al. | 0.5 | 1 | 0 | 0 | 1 | 1 | 1 |
| Q2 | Cheng Si et al. | 0 | 1 | 0 | 0 | 1 | 1 | 1 |

Reviewer 2

| Question | Author | Cut-off analysis | Discrimination statistics | Calibration statistics | Prospective study | Validation | Comparison to gold standard | Potential clinical applications | Cost-analysis |
|---|---|---|---|---|---|---|---|---|---|
| Q1 | Wei et al. | 0 | 1 | 0.5 | 0 | 0.4 | 1 | 0 | 0 |
| Q1 | Yang J et al. | 0 | 0.5 | 0 | 0 | 0.4 | 0 | 1 | 0 |
| Q1 | Xie H et al. | 0 | 0.5 | 1 | 0 | 0 | 0 | 1 | 0 |
| Q1 | Chen HY et al. | 0 | 1 | 0 | 0 | 0.6 | 1 | 1 | 0 |
| Q1 | Chen Sh et al. | 0 | 0.5 | 0.5 | 0 | 0.4 | 1 | 1 | 0 |
| Q1 | Gao et al. | 0 | 0.5 | 0.5 | 0 | 0.4 | 1 | 1 | 0 |
| Q1 | Xie T et al. | 1 | 1 | 0 | 0 | 0.4 | 1 | 0 | 0 |
| Q1 | Awe et al. | 0 | 1 | 0.5 | 0 | 0.4 | 0 | 0 | 0 |
| Q1 | Dong et al. | 0 | 0.5 | 0.5 | 0 | 0.4 | 0 | 1 | 0 |
| Q1 | Yang R et al. | 0 | 1 | 0 | 0 | 0.4 | 0 | 1 | 0 |
| Q1 | Liang et al. | 0 | 1 | 1 | 0 | 0.4 | 0 | 1 | 0 |
| Q1 | Chu et al. | 0 | 0.5 | 0 | 0 | 0.4 | 1 | 1 | 0 |
| Q1 | Fang et al. | 0 | 0.5 | 1 | 0 | 0.4 | 0 | 1 | 0 |
| Q2 | Ansari et al. | 0 | 0.5 | 0 | 0 | 0.4 | 0 | 1 | 0 |
| Q2 | Torra-Ferrer et al. | 0 | 1 | 0.5 | 0 | 0.8 | 0 | 1 | 0 |
| Q2 | Hanania et al. | 0 | 1 | 0.5 | 0 | 0.4 | 1 | 1 | 0 |
| Q2 | Permuth et al. | 0 | 1 | 0 | 0 | 0.4 | 1 | 0 | 0 |
| Q2 | Chakraborty et al. | 0 | 1 | 0 | 0 | 0.4 | 1 | 0 | 0 |
| Q2 | Polk et al. | 0 | 1 | 0 | 0 | 0.4 | 1 | 0 | 0 |
| Q2 | Tobaly et al. | 1 | 1 | 0.5 | 0 | 0.6 | 0 | 1 | 0 |
| Q2 | Harrington et al. | 0 | 0.5 | 0 | 0 | 0.4 | 1 | 0 | 0 |
| Q2 | Li et al | 0 | 1 | 0.5 | 0 | 0.4 | 0 | 0 | 0 |
| Q2 | Cui et al. | 1 | 1 | 1 | 0 | 0.8 | 1 | 1 | 0 |
| Q2 | Cheng Sh et al. | 0 | 1 | 0.5 | 0 | 0.4 | 1 | 0 | 0 |
| Q2 | Wang et al. | 0 | 1 | 0 | 0 | 0.8 | 1 | 0 | 0 |
| Q2 | Flammia et al. | 0 | 1 | 0 | 0 | 0.4 | 0 | 1 | 0 |
| Q2 | Lee et al. | 0 | 1 | 0 | 0 | 0.4 | 0 | 0 | 0 |
| Q2 | Lou et al. | 0 | 1 | 0 | 0 | 0.4 | 0 | 0 | 0 |
| Q2 | Cheng Si et al. | 1 | 1 | 0.5 | 0 | 1 | 0 | 1 | 1 |

Reviewer 2

| Question | Author | Open science and data | Total | % |
|---|---|---|---|---|
| Q1 | Wei et al. | 0 | 13 | 0.361 |
| Q1 | Yang J et al. | 0 | 10 | 0.278 |
| Q1 | Xie H et al. | 0 | 11 | 0.306 |
| Q1 | Chen HY et al. | 0 | 15 | 0.417 |
| Q1 | Chen Sh et al. | 0 | 16 | 0.444 |
| Q1 | Gao et al. | 0 | 15 | 0.417 |
| Q1 | Xie T et al. | 0 | 12 | 0.333 |
| Q1 | Awe et al. | 0.25 | 10 | 0.278 |
| Q1 | Dong et al. | 0 | 12 | 0.333 |
| Q1 | Yang R et al. | 0 | 12 | 0.333 |
| Q1 | Liang et al. | 0 | 14 | 0.389 |
| Q1 | Chu et al. | 0 | 13 | 0.361 |
| Q1 | Fang et al. | 0 | 13 | 0.361 |
| Q2 | Ansari et al. | 0 | 9 | 0.250 |
| Q2 | Torra-Ferrer et al. | 0 | 15 | 0.417 |
| Q2 | Hanania et al. | 0 | 15 | 0.417 |
| Q2 | Permuth et al. | 0 | 13 | 0.361 |
| Q2 | Chakraborty et al. | 0 | 12 | 0.333 |
| Q2 | Polk et al. | 0 | 13 | 0.361 |
| Q2 | Tobaly et al. | 0 | 15 | 0.417 |
| Q2 | Harrington et al. | 0 | 11 | 0.306 |
| Q2 | Li et al | 0 | 12 | 0.3330 |
| Q2 | Cui et al. | 0 | 20 | 0.556 |
| Q2 | Cheng Sh et al. | 0 | 12 | 0.333 |
| Q2 | Wang et al. | 0 | 14 | 0.389 |
| Q2 | Flammia et al. | 0 | 12 | 0.333 |
| Q2 | Lee et al. | 0 | 9 | 0.250 |
| Q2 | Lou et al. | 0 | 11 | 0.306 |
| Q2 | Cheng Si et al. | 0 | 18 | 0.500 |

**Table S10. TRIPOD-AI assessment**. Results for Q1 and Q2, reported separately for both independent reviewers.

Reviewer 1

| Question | Author | Title (1) | Abstract (1) | Introduction (4) | Methods (26) | Open Science (6) |
|---|---|---|---|---|---|---|
| Q1 | Wei et al. | 1 | 1 | 0.625 | 0.635 | 0.583 |
| Q1 | Yang J et al. | 1 | 1 | 0.750 | 0.673 | 0.333 |
| Q1 | Xie H et al. | 1 | 1 | 0.875 | 0.808 | 0.417 |
| Q1 | Chen HY et al. | 1 | 1 | 1.000 | 0.769 | 0.333 |
| Q1 | Chen Sh et al. | 1 | 1 | 0.750 | 0.692 | 0.833 |
| Q1 | Gao et al. | 1 | 1 | 1.000 | 1.000 | 0.750 |
| Q1 | Xie T et al. | 1 | 1 | 0.750 | 0.712 | 1.000 |
| Q1 | Awe et al. | 1 | 1 | 0.750 | 0.788 | 0.667 |
| Q1 | Dong et al. | 1 | 1 | 0.750 | 0.692 | 0.417 |
| Q1 | Yang R et al. | 1 | 1 | 0.750 | 0.615 | 0.583 |
| Q1 | Liang et al. | 1 | 1 | 0.750 | 0.788 | 0.417 |
| Q1 | Chu et al. | 1 | 1 | 0.750 | 0.673 | 0.333 |
| Q1 | Fang et al. | 1 | 1 | 0.750 | 0.731 | 0.417 |
| Q1 | Ansari et al. | 1 | 1 | 0.750 | 0.673 | 0.333 |
| Q1 | Torra-Ferrer et al. | 1 | 1 | 0.750 | 0.885 | 0.750 |
| Q2 | Hanania et al. | 1 | 1 | 0.750 | 0.558 | 0.333 |
| Q2 | Permuth et al. | 1 | 1 | 0.750 | 0.673 | 0.500 |
| Q2 | Chakraborty et al. | 1 | 1 | 0.750 | 0.769 | 0.500 |
| Q2 | Polk et al. | 1 | 1 | 0.750 | 0.500 | 0.333 |
| Q2 | Tobaly et al. | 1 | 1 | 0.750 | 0.865 | 0.500 |
| Q2 | Harrington et al. | 1 | 1 | 0.750 | 0.615 | 0.500 |
| Q2 | Li et al | 1 | 1 | 0.875 | 0.538 | 0.333 |
| Q2 | Cui et al. | 1 | 1 | 1.000 | 1.000 | 1.000 |
| Q2 | Cheng Sh et al. | 1 | 1 | 0.750 | 0.750 | 0.500 |
| Q2 | Wang et al. | 1 | 1 | 0.750 | 0.846 | 0.333 |
| Q2 | Flammia et al. | 1 | 1 | 0.750 | 0.635 | 0.333 |
| Q2 | Lee et al. | 1 | 1 | 0.750 | 0.673 | 0.500 |
| Q2 | Lou et al. | 1 | 1 | 0.750 | 0.673 | 0.500 |
| Q2 | Cheng Si et al. | 1 | 1 | 0.750 | 0.750 | 0.500 |

Reviewer 1

| Question | Author | Patient & Public Involvement (1) | Results (8) | Discussion (5) | Overall points | Overall % |
|---|---|---|---|---|---|---|
| Q1 | Wei et al. | 0.5 | 0.500 | 0.600 | 32.000 | 0.615 |
| Q1 | Yang J et al. | 0.5 | 0.438 | 0.900 | 33.000 | 0.635 |
| Q1 | Xie H et al. | 0.5 | 0.750 | 0.900 | 40.000 | 0.769 |
| Q1 | Chen HY et al. | 1.0 | 0.563 | 0.600 | 36.500 | 0.702 |
| Q1 | Chen Sh et al. | 0.0 | 0.688 | 0.800 | 37.500 | 0.721 |
| Q1 | Gao et al. | 0.5 | 0.500 | 0.400 | 43.000 | 0.827 |
| Q1 | Xie T et al. | 0.5 | 0.438 | 0.700 | 37.000 | 0.712 |
| Q1 | Awe et al. | 1.0 | 0.438 | 0.800 | 38.000 | 0.731 |
| Q1 | Dong et al. | 1.0 | 0.375 | 0.800 | 33.500 | 0.644 |
| Q1 | Yang R et al. | 0.0 | 0.438 | 0.600 | 31.000 | 0.596 |
| Q1 | Liang et al. | 0.0 | 0.813 | 0.700 | 38.000 | 0.731 |
| Q1 | Chu et al. | 0.5 | 0.375 | 0.700 | 31.500 | 0.606 |
| Q1 | Fang et al. | 0.0 | 0.750 | 0.600 | 35.500 | 0.683 |
| Q1 | Ansari et al. | 1.0 | 0.375 | 0.700 | 32.000 | 0.615 |
| Q1 | Torra-Ferrer et al. | 0.0 | 1.000 | 0.900 | 45.000 | 0.865 |
| Q2 | Hanania et al. | 0.0 | 0.563 | 0.600 | 29.000 | 0.558 |
| Q2 | Permuth et al. | 0.0 | 0.813 | 0.600 | 35.000 | 0.673 |
| Q2 | Chakraborty et al. | 0.0 | 0.688 | 0.500 | 36.000 | 0.692 |
| Q2 | Polk et al. | 0.5 | 0.750 | 0.600 | 29.500 | 0.567 |
| Q2 | Tobaly et al. | 1.0 | 0.750 | 0.800 | 41.500 | 0.798 |
| Q2 | Harrington et al. | 0.5 | 0.313 | 0.500 | 29.500 | 0.567 |
| Q2 | Li et al | 0.0 | 0.563 | 0.500 | 28.500 | 0.548 |
| Q2 | Cui et al. | 1.0 | 0.563 | 0.100 | 44.000 | 0.846 |
| Q2 | Cheng Sh et al. | 0.0 | 0.625 | 0.600 | 35.500 | 0.683 |
| Q2 | Wang et al. | 0.0 | 0.875 | 0.800 | 40.000 | 0.769 |
| Q2 | Flammia et al. | 0.0 | 0.563 | 0.600 | 31.000 | 0.596 |
| Q2 | Lee et al. | 0.0 | 0.875 | 0.500 | 35.000 | 0.673 |
| Q2 | Lou et al. | 0.0 | 0.438 | 0.700 | 32.500 | 0.625 |
| Q2 | Cheng Si et al. | 1.0 | 0.688 | 0.600 | 37.000 | 0.712 |

Reviewer 2

| Question | Author | Title (1) | Abstract (1) | Introduction (4) | Methods (26) | Open Science (6) |
|---|---|---|---|---|---|---|
| Q1 | Wei et al. | 1 | 1 | 0.750 | 0.635 | 0.333 |
| Q1 | Yang J et al. | 1 | 1 | 0.750 | 0.673 | 0.333 |
| Q1 | Xie H et al. | 1 | 1 | 0.750 | 0.769 | 0.417 |
| Q1 | Chen HY et al. | 1 | 1 | 0.750 | 0.712 | 0.750 |
| Q1 | Chen Sh et al. | 1 | 1 | 0.750 | 0.788 | 0.500 |
| Q1 | Gao et al. | 1 | 1 | 0.750 | 0.865 | 0.833 |
| Q1 | Xie T et al. | 1 | 1 | 0.750 | 0.731 | 0.500 |
| Q1 | Awe et al. | 1 | 1 | 0.750 | 0.827 | 0.667 |
| Q1 | Dong et al. | 1 | 1 | 0.750 | 0.692 | 0.333 |
| Q1 | Yang R et al. | 1 | 1 | 0.750 | 0.615 | 0.417 |
| Q1 | Liang et al. | 1 | 1 | 0.750 | 0.885 | 0.500 |
| Q1 | Chu et al. | 1 | 1 | 0.750 | 0.615 | 0.333 |
| Q1 | Fang et al. | 1 | 1 | 0.750 | 0.769 | 0.167 |
| Q1 | Ansari et al. | 1 | 1 | 0.750 | 0.654 | 0.333 |
| Q1 | Torra-Ferrer et al. | 1 | 1 | 0.750 | 0.904 | 0.500 |
| Q2 | Hanania et al. | 1 | 1 | 0.750 | 0.481 | 0.333 |
| Q2 | Permuth et al. | 1 | 1 | 0.750 | 0.538 | 0.333 |
| Q2 | Chakraborty et al. | 1 | 1 | 0.750 | 0.635 | 0.167 |
| Q2 | Polk et al. | 1 | 1 | 0.750 | 0.500 | 0.417 |
| Q2 | Tobaly et al. | 1 | 1 | 0.750 | 0.865 | 0.417 |
| Q2 | Harrington et al. | 1 | 1 | 0.750 | 0.558 | 0.333 |
| Q2 | Li et al | 1 | 1 | 0.750 | 0.538 | 0.333 |
| Q2 | Cui et al. | 1 | 1 | 0.750 | 0.904 | 0.667 |
| Q2 | Cheng Sh et al. | 1 | 1 | 0.750 | 0.673 | 0.500 |
| Q2 | Wang et al. | 1 | 1 | 0.750 | 0.942 | 0.333 |
| Q2 | Flammia et al. | 1 | 1 | 0.750 | 0.462 | 0.333 |
| Q2 | Lee et al. | 1 | 1 | 0.750 | 0.750 | 0.333 |
| Q2 | Lou et al. | 1 | 1 | 0.750 | 0.654 | 0.333 |
| Q2 | Cheng Si et al. | 1 | 1 | 0.750 | 0.788 | 0.500 |

Reviewer 2

| Question | Author | Patient & Public Involvement (1) | Results (8) | Discussion (5) | Overall points | Overall % |
|---|---|---|---|---|---|---|
| Q1 | Wei et al. | 1 | 0.375 | 0.600 | 30.500 | 0.587 |
| Q1 | Yang J et al. | 1 | 0.438 | 0.600 | 32.000 | 0.615 |
| Q1 | Xie H et al. | 1 | 0.688 | 0.800 | 38.000 | 0.731 |
| Q1 | Chen HY et al. | 1 | 0.438 | 0.700 | 36.000 | 0.692 |
| Q1 | Chen Sh et al. | 1 | 0.813 | 0.600 | 39.000 | 0.750 |
| Q1 | Gao et al. | 1 | 0.875 | 0.800 | 44.500 | 0.856 |
| Q1 | Xie T et al. | 1 | 0.500 | 0.700 | 35.500 | 0.683 |
| Q1 | Awe et al. | 1 | 0.438 | 0.900 | 39.500 | 0.760 |
| Q1 | Dong et al. | 1 | 0.250 | 0.600 | 31.000 | 0.596 |
| Q1 | Yang R et al. | 1 | 0.250 | 0.600 | 29.500 | 0.567 |
| Q1 | Liang et al. | 1 | 0.625 | 0.600 | 40.000 | 0.769 |
| Q1 | Chu et al. | 1 | 0.375 | 0.600 | 30.000 | 0.577 |
| Q1 | Fang et al. | 1 | 0.625 | 0.600 | 35.000 | 0.673 |
| Q1 | Ansari et al. | 1 | 0.188 | 0.600 | 29.500 | 0.567 |
| Q1 | Torra-Ferrer et al. | 1 | 0.750 | 0.900 | 43.000 | 0.827 |
| Q2 | Hanania et al. | 1 | 0.563 | 0.600 | 28.000 | 0.538 |
| Q2 | Permuth et al. | 1 | 0.563 | 0.600 | 29.500 | 0.567 |
| Q2 | Chakraborty et al. | 1 | 0.500 | 0.600 | 30.500 | 0.587 |
| Q2 | Polk et al. | 1 | 0.563 | 0.600 | 29.000 | 0.558 |
| Q2 | Tobaly et al. | 1 | 0.875 | 0.800 | 42.000 | 0.808 |
| Q2 | Harrington et al. | 1 | 0.313 | 0.600 | 28.000 | 0.538 |
| Q2 | Li et al | 1 | 0.188 | 0.600 | 26.500 | 0.510 |
| Q2 | Cui et al. | 1 | 0.625 | 0.700 | 42.000 | 0.808 |
| Q2 | Cheng Sh et al. | 1 | 0.438 | 0.700 | 33.500 | 0.644 |
| Q2 | Wang et al. | 1 | 0.563 | 0.800 | 41.000 | 0.788 |
| Q2 | Flammia et al. | 1 | 0.313 | 0.600 | 25.500 | 0.490 |
| Q2 | Lee et al. | 1 | 0.563 | 0.600 | 35.000 | 0.673 |
| Q2 | Lou et al. | 1 | 0.375 | 0.700 | 31.500 | 0.606 |
| Q2 | Cheng Si et al. | 1 | 0.688 | 0.800 | 39.000 | 0.750 |

**Table S11. PROBAST-AI assessment**. Results for Q1 and Q2, reported separately for both independent reviewers.

| Reviewer 1 | | Model development- Quality Concern | | | | |
|---|---|---|---|---|---|---|
| Question | Author | Participants | Predictors | Outcome | Analysis | Overall Quality Concern |
| Q1 | Wei et al. | low | unclear | unclear | low | unclear |
| Q1 | Yang J et al. | low | low / unclear | low | unclear | unclear |
| Q1 | Xie H et al. | low | unclear | low | unclear | unclear |
| Q1 | Chen HY et al. | low | low | low | low | low |
| Q1 | Chen Sh et al. | low | low | low | unclear | unclear |
| Q1 | Gao et al. | low | unclear | low | low | low |
| Q1 | Xie T et al. | low | low | low | low | low |
| Q1 | Awe et al. | low | unclear | low | low | unclear |
| Q1 | Dong et al. | low | unclear | low | unclear | unclear |
| Q1 | Yang R et al. | low | unclear | low | unclear | unclear |
| Q1 | Liang et al. | low | low | low | unclear | low |
| Q1 | Chu et al. | low | low | unclear | unclear | unclear |
| Q1 | Fang et al. | low | unclear | low | unclear | unclear |
| Q1 | Ansari et al. | low | high | high | high | high |
| Q1 | Torra-Ferrer et al. | low | low | low | low | low |
| Q2 | Hanania et al. | low | unclear | unclear | unclear | unclear |
| Q2 | Permuth et al. | low | unclear | low | unclear | unclear |
| Q2 | Chakraborty et al. | low | low | low | unclear | unclear |
| Q2 | Polk et al. | low | unclear | low | unclear | unclear |
| Q2 | Tobaly et al. | low | low | low | low | low |
| Q2 | Harrington et al. | unclear | unclear | unclear | unclear | unclear |
| Q2 | Li et al | low | unclear | low | low | unclear |
| Q2 | Cui et al. | low | low | low | unclear | unclear |
| Q2 | Cheng Sh et al. | high | unclear | high | high | high |
| Q2 | Wang et al. | low | low | low | low | low |
| Q2 | Flammia et al. | unclear | low | low | unclear | low |
| Q2 | Lee et al. | low | low | low | unclear | unclear |
| Q2 | Lou et al. | low | unclear | unclear | unclear | unclear |
| Q2 | Cheng Si et al. | low | low | low | low | low |

| Reviewer 1 | | Model development - Concern of Applicability | | | | Model evaluation - Concern of Applicability | | | |
|---|---|---|---|---|---|---|---|---|---|
| Question | Author | Participants | Predictors | Outcome | Overall Concern | Participants | Predictors | Outcome | Overall concern |
| Q1 | Wei et al. | low | low | low | low | low | low | low | low |
| Q1 | Yang J et al. | low | low | low | low | low | low | low | low |
| Q1 | Xie H et al. | low | low | low | low | low | low | low | low |
| Q1 | Chen HY et al. | low | low | low | low | low | low | low | low |
| Q1 | Chen Sh et al. | low | low | low | low | low | low | low | low |
| Q1 | Gao et al. | low | low | low | low | low | low | low | low |
| Q1 | Xie T et al. | low | low | low | low | low | low | low | low |
| Q1 | Awe et al. | low | unclear | unclear | unclear | low | unclear | unclear | unclear |
| Q1 | Dong et al. | low | low | low | low | low | low | low | low |
| Q1 | Yang R et al. | low | low | low | low | low | low | low | low |
| Q1 | Liang et al. | low | low | low | low | low | low | low | low |
| Q1 | Chu et al. | low | low | low | low | low | low | low | low |
| Q1 | Fang et al. | low | low | low | low | low | low | low | low |
| Q1 | Ansari et al. | low | low | low | low | low | low | low | low |
| Q1 | Torra-Ferrer et al. | low | low | low | low | low | low | low | low |
| Q2 | Hanania et al. | low | low | low | low | low | low | low | low |
| Q2 | Permuth et al. | low | low | low | low | low | low | low | low |
| Q2 | Chakraborty et al. | low | low | low | low | low | low | low | low |
| Q2 | Polk et al. | low | unclear | low | unclear | low | unclear | low | low |
| Q2 | Tobaly et al. | low | low | low | low | low | low | low | low |
| Q2 | Harrington et al. | unclear | unclear | low | unclear | unclear | unclear | low | unclear |
| Q2 | Li et al | low | low | low | low | low | low | low | unclear |
| Q2 | Cui et al. | low | low | low | low | low | low | low | low |
| Q2 | Cheng Sh et al. | unclear | low | unclear | unclear | low | low | unclear | unclear |
| Q2 | Wang et al. | low | low | low | low | low | low | low | low |
| Q2 | Flammia et al. | low | low | low | low | low | low | low | low |
| Q2 | Lee et al. | low | low | low | low | low | low | low | low |
| Q2 | Lou et al. | low | low | low | low | low | low | low | low |
| Q2 | Cheng Si et al. | low | low | low | low | low | low | low | low |

Reviewer 1 Model evaluation - Risk of bias

| Question | Author | Participants | Predictors | Outcome | Analysis | Overall Risk of Bias | Total Score points | Total % |
|---|---|---|---|---|---|---|---|---|
| Q1 | Wei et al. | low | unclear | unclear | unclear | unclear | 35.25 | 0.678 |
| Q1 | Yang J et al. | low | unclear | low | unclear | unclear | 34.5 | 0.719 |
| Q1 | Xie H et al. | low | low | low | unclear | unclear | 37.25 | 0.716 |
| Q1 | Chen HY et al. | low | low | low | low | low | 43.5 | 0.906 |
| Q1 | Chen Sh et al. | low | low | low | unclear | unclear | 42.75 | 0.891 |
| Q1 | Gao et al. | low | unclear | low | unclear | unclear | 41 | 0.788 |
| Q1 | Xie T et al. | low | low | low | unclear | unclear | 41.75 | 0.803 |
| Q1 | Awe et al. | low | high | low | unclear | high | 39.5 | 0.823 |
| Q1 | Dong et al. | low | high | low | unclear | high | 41.5 | 0.865 |
| Q1 | Yang R et al. | low | unclear | low | unclear | unclear | 39.75 | 0.828 |
| Q1 | Liang et al. | low | unclear | low | unclear | unclear | 37.75 | 0.786 |
| Q1 | Chu et al. | low | low | high | high | high | 38.25 | 0.797 |
| Q1 | Fang et al. | low | unclear | low | low | unclear | 42.5 | 0.885 |
| Q1 | Ansari et al. | low | unclear | low | high | high | 38 | 0.792 |
| Q1 | Torra-Ferrer et al. | low | low | low | low | low | 43.75 | 0.911 |
| Q2 | Hanania et al. | low | unclear | unclear | high | high | 30 | 0.625 |
| Q2 | Permuth et al. | low | unclear | low | high | high | 35 | 0.673 |
| Q2 | Chakraborty et al. | low | high | low | unclear | high | 38 | 0.731 |
| Q2 | Polk et al. | unclear | high | low | high | high | 29 | 0.558 |
| Q2 | Tobaly et al. | low | low | low | low | low | 43 | 0.896 |
| Q2 | Harrington et al. | unclear | high | unclear | high | high | 30 | 0.625 |
| Q2 | Li et al | low | unclear | unclear | unclear | unclear | 35.25 | 0.678 |
| Q2 | Cui et al. | low | low | low | low | low | 43.5 | 0.906 |
| Q2 | Cheng Sh et al. | low | unclear | unclear | unclear | unclear | 30.75 | 0.641 |
| Q2 | Wang et al. | low | low | low | low | low | 44.25 | 0.922 |
| Q2 | Flammia et al. | unclear | high | low | unclear | high | 35.25 | 0.734 |
| Q2 | Lee et al. | low | low | low | unclear | unclear | 44.25 | 0.922 |
| Q2 | Lou et al. | low | high | unclear | unclear | unclear | 36.5 | 0.760 |
| Q2 | Cheng Si et al. | low | low | low | low | low | 42 | 0.875 |

Reviewer 2 | | Model development- Quality Concern | | | | 
| --- | --- | --- | --- | --- | --- | --- |
| Question | Author | Participants | Predictors | Outcome | Analysis | Overall Quality Concern |
| Q1 | Wei et al. | low | low | unclear | unclear | unclear |
| Q1 | Yang J et al. | low | unclear | low | unclear | unclear |
| Q1 | Xie H et al. | low | low | unlcear | high | high |
| Q1 | Chen HY et al. | low | low | low | low | low |
| Q1 | Chen Sh et al. | low | low | low | low | unclear |
| Q1 | Gao et al. | low | unclear | low | unclear | unclear |
| Q1 | Xie T et al. | low | low | low | high | high |
| Q1 | Awe et al. | low | high | unclear | unclear | high |
| Q1 | Dong et al. | low | high | low | high | unclear |
| Q1 | Yang R et al. | low | unclear | low | unclear | unclear |
| Q1 | Liang et al. | low | unclear | unclear | unclear | unlcear |
| Q1 | Chu et al. | low | low | high | high | high |
| Q1 | Fang et al. | low | unclear | unclear | unclear | high |
| Q1 | Ansari et al. | low | high | unclear | high | high |
| Q1 | Torra-Ferrer et al. | low | low | low | low | low |
| Q2 | Hanania et al. | low | unclear | high | high | unclear |
| Q2 | Permuth et al. | low | unclear | low | unclear | unclear |
| Q2 | Chakraborty et al. | low | high | unclear | unclear | high |
| Q2 | Polk et al. | low | high | unclear | unclear | high |
| Q2 | Tobaly et al. | low | low | low | low | low |
| Q2 | Harrington et al. | unclear | high | unclear | high | high |
| Q2 | Li et al | low | unclear | unclear | high | high |
| Q2 | Cui et al. | low | low | low | low | low |
| Q2 | Cheng Sh et al. | high | unclear | unclear | high | high |
| Q2 | Wang et al. | low | low | low | low | low |
| Q2 | Flammia et al. | unclear | high | low | high | high |
| Q2 | Lee et al. | low | low | low | high | high |
| Q2 | Lou et al. | low | high | unclear | high | high |
| Q2 | Cheng Si et al. | low | unclear | low | low | unclear |

| Reviewer 2 | | Model development - Concern of Applicability | | | | Model evaluation - Concern of Applicability | | | |
|---|---|---|---|---|---|---|---|---|---|
| Question | Author | Participants | Predictors | Outcome | Overall Concern | Participants | Predictors | Outcome | Overall concern |
| Q1 | Wei et al. | low | low | low | low | low | low | low | low |
| Q1 | Yang J et al. | low | low | low | low | low | low | low | low |
| Q1 | Xie H et al. | low | low | low | low | low | low | low | low |
| Q1 | Chen HY et al. | low | low | low | low | low | low | low | low |
| Q1 | Chen Sh et al. | low | low | low | low | low | low | low | low |
| Q1 | Gao et al. | low | low | low | low | low | low | low | low |
| Q1 | Xie T et al. | low | low | low | low | low | low | low | low |
| Q1 | Awe et al. | low | low | low | low | low | low | low | low |
| Q1 | Dong et al. | low | low | low | low | low | low | low | low |
| Q1 | Yang R et al. | low | low | low | low | low | low | low | low |
| Q1 | Liang et al. | low | low | low | low | low | low | low | low |
| Q1 | Chu et al. | low | low | low | low | low | low | low | low |
| Q1 | Fang et al. | low | low | low | low | low | low | low | low |
| Q1 | Ansari et al. | low | low | low | low | low | low | low | low |
| Q1 | Torra-Ferrer et al. | low | low | low | low | low | low | low | low |
| Q2 | Hanania et al. | low | low | low | low | low | low | low | low |
| Q2 | Permuth et al. | low | low | low | low | low | low | low | low |
| Q2 | Chakraborty et al. | low | low | low | low | low | low | low | low |
| Q2 | Polk et al. | low | low | low | low | low | low | low | low |
| Q2 | Tobaly et al. | low | low | low | low | low | low | low | low |
| Q2 | Harrington et al. | low | unclear | low | unclear | unclear | unclear | low | low |
| Q2 | Li et al | low | low | low | low | low | low | low | low |
| Q2 | Cui et al. | low | low | low | low | low | low | low | low |
| Q2 | Cheng Sh et al. | unclear | low | unclear | unclear | low | low | low | low |
| Q2 | Wang et al. | low | low | low | low | low | low | low | low |
| Q2 | Flammia et al. | low | low | low | low | low | low | low | low |
| Q2 | Lee et al. | low | low | low | low | low | low | low | low |
| Q2 | Lou et al. | low | low | low | low | low | low | low | low |
| Q2 | Cheng Si et al. | low | low | low | low | low | low | low | low |

| Reviewer 2 | | Model evaluation - Risk of bias | | | | | | |
|---|---|---|---|---|---|---|---|---|
| Question | Author | Participants | Predictors | Outcome | Analysis | Overall Risk of Bias category | Total Score points | Total % |
| Q1 | Wei et al. | low | high | unclear | high | high | 28 | 0.583 |
| Q1 | Yang J et al. | low | unclear | unclear | unclear | unclear | 31.25 | 0.651 |
| Q1 | Xie H et al. | low | low | unclear | high | high | 37.75 | 0.786 |
| Q1 | Chen HY et al. | low | low | low | low | low | 40.75 | 0.849 |
| Q1 | Chen Sh et al. | low | low | low | unclear | unclear | 41.25 | 0.859 |
| Q1 | Gao et al. | low | unclear | low | unclear | unclear | 39.25 | 0.818 |
| Q1 | Xie T et al. | low | low | low | high | high | 35.5 | 0.740 |
| Q1 | Awe et al. | low | high | unclear | high | high | 36.75 | 0.766 |
| Q1 | Dong et al. | low | high | low | high | high | 38.25 | 0.797 |
| Q1 | Yang R et al. | low | unclear | low | unclear | unclear | 38.75 | 0.807 |
| Q1 | Liang et al. | low | unclear | unclear | unclear | unclear | 31 | 0.646 |
| Q1 | Chu et al. | low | low | high | high | high | 37.25 | 0.776 |
| Q1 | Fang et al. | low | unclear | low | unclear | unclear | 41 | 0.854 |
| Q1 | Ansari et al. | low | unclear | low | high | high | 37 | 0.771 |
| Q1 | Torra-Ferrer et al. | low | low | low | low | low | 44 | 0.917 |
| Q2 | Hanania et al. | low | unclear | high | high | high | 34.5 | 0.719 |
| Q2 | Permuth et al. | low | unclear | low | high | high | 36.5 | 0.760 |
| Q2 | Chakraborty et al. | low | high | unclear | high | high | 31.75 | 0.661 |
| Q2 | Polk et al. | low | high | unclear | high | high | 29.75 | 0.620 |
| Q2 | Tobaly et al. | low | low | low | low | low | 43 | 0.896 |
| Q2 | Harrington et al. | unclear | high | unclear | high | high | 27.75 | 0.578 |
| Q2 | Li et al | low | unclear | unclear | unclear | unclear | 34 | 0.708 |
| Q2 | Cui et al. | low | low | low | low | low | 45 | 0.938 |
| Q2 | Cheng Sh et al. | low | unclear | unclear | unclear | unclear | 33 | 0.688 |
| Q2 | Wang et al. | low | low | low | low | low | 44.5 | 0.927 |
| Q2 | Flammia et al. | unclear | high | low | high | high | 33.75 | 0.703 |
| Q2 | Lee et al. | low | low | low | high | high | 42.75 | 0.891 |
| Q2 | Lou et al. | low | high | unclear | high | high | 34.5 | 0.719 |
| Q2 | Cheng Si et al. | low | low | low | low | low | 41 | 0.854 |
